# Cost-effectiveness of COVID-19 vaccination in low- and middle-income countries

**DOI:** 10.1101/2021.04.28.21256237

**Authors:** Mark J. Siedner, Christopher Alba, Kieran P. Fitzmaurice, Rebecca F. Gilbert, Justine A. Scott, Fatma M. Shebl, Andrea Ciaranello, Krishna P. Reddy, Kenneth A. Freedberg

## Abstract

Despite the advent of safe and highly effective COVID-19 vaccines^1–4^, pervasive inequities in global distribution persist^5^. In response, multinational partners have proposed programs to allocate vaccines to low- and middle-income countries (LMICs)^6^. Yet, there remains a substantial funding gap for such programs^7^. Further, the optimal vaccine supply is unknown and the cost-effectiveness of investments into global vaccination programs has not been described. We used a validated COVID-19 simulation model^8^ to project the health benefits and costs of reaching 20%-70% vaccine coverage in 91 LMICs. We show that funding 20% vaccine coverage over one year among 91 LMICs would prevent 294 million infections and 2 million deaths, with 26 million years of life saved at a cost of US$6.4 billion, for an incremental cost effectiveness ratio (ICER) of US$250/year of life saved (YLS). Increasing vaccine coverage up to 50% would prevent millions more infections and save hundreds of thousands of additional lives, with ICERs below US$8,000/YLS. Results were robust to variations in vaccine efficacy and hesitancy, but were more sensitive to assumptions about epidemic pace and vaccination costs. These results support efforts to fund vaccination programs in LMICs and complement arguments about health equity^9^, economic benefits^10^, and pandemic control^11^.

## Introduction

By late April 2021, over 147 million cases and over 3 million deaths were attributed to the COVID-19 pandemic globally^12^. Approximately 53% of COVID-19 deaths have been reported in low- and middle-income countries (LMICs)^13^, although such estimates may underestimate the epidemic’s health impacts^14,15^ and do not account for disruptions in economic productivity, healthcare delivery, and social wellbeing^16–20^. Nonetheless, there has been major progress toward containing the pandemic with the advent and licensure of multiple highly effective vaccines for prevention of COVID-19^1–4^.

Although safe and effective vaccines provide a strategic path out of the pandemic, their benefits have thus far largely been confined to high and upper-middle income countries, which have secured contracts for purchase and distribution of nearly 90% of the global vaccine supply^5^. LMICs have had less success in procuring vaccines^5^, but global initiatives to ameliorate this gap are underway. The COVAX Advance Market Commitment (AMC) program aims to ensure access to low-cost SARS-CoV-2 vaccines to 92 LMICs^7^. By February 2021, higher-income governments and donors had committed over US$6.3 billion to this initiative^21^. However, there remains a funding shortfall for the COVAX AMC initiative to achieve initial goals of vaccine delivery for 20% of the population in LMICs^7^. Moreover, the 20% vaccine target set out by COVAX was an initial target is below the projected levels needed to achieve epidemic control^22^. Whereas the US government and other high-income countries have made longstanding commitments to development assistance for health in LMICs, with an estimated US$39 billion contributed worldwide in 2018 alone^23^, the costs, benefits, and cost-effectiveness of donations into the global COVID-19 vaccine supply have not been estimated.

Our objective was to assess the clinical benefits (COVID-19 infections, hospitalizations, deaths, life years saved), program costs, and value (incremental cost-effectiveness ratios [ICERs]) of donor outlays into the global vaccine supply for 91 LMICs (we excluded India, a COVAX-eligible country, because of its plan to produce vaccines domestically^24^).

We used the Clinical and Economic Analysis of COVID-19 interventions (CEACOV) model, a validated, dynamic microsimulation of the natural history of COVID-19^8,25–27^ (Supplementary Methods and Supplementary Fig. 1). We modeled discrete epidemics over 360 days in each country using country-specific data on age distribution, population size, and hospital and ICU bed capacities (Supplementary Table 1). We compared a situation in which participating LMICs have no vaccine access to one in which vaccination supplies reach 20%-70% population coverage, with vaccines prioritized to individuals aged 60 years or older, and increasing in 10% increments. In our base case, we assumed a modest epidemic growth rate (effective reproductive number [R*_e_*] of 1.2), and used data from the Johnson & Johnson (J&J)/Janssen Ad26.COV2.S vaccine trial to inform vaccine efficacy^1^ (Extended Data Table 1). Costs were from the donor perspective, and included fixed costs for the vaccination program (planning, training, social mobilization, cold chain equipment, and pharmacovigilance^28^) and variable costs per person vaccinated (vaccines, logistics and delivery and technical assistance) ^28,29^ (Extended Data Table 1). We conducted sensitivity analyses among a subset of nine countries, chosen based on a cluster analysis, to assess robustness of our estimates to assumptions about epidemic severity, as well as vaccination hesistancy, efficacy, rollout pace and costs.

## Results

### Benefits and costs of global vaccination

In the base case analysis, achieving 20% vaccine coverage (508 million people vaccinated) in 91 LMICs would decrease infections by over 50% in the following year, from 547 million to 253 million, and decrease projected COVID-19 deaths nearly 80%, from 2.4 million to 512,000, saving 26 million years of life compared to no vaccine coverage (Table 1; individual country and regional estimates are in Supplementary Table 3). Total vaccination program costs to achieve 20% coverage would be US$6.4 billion, resulting in an ICER of US$20/infection prevented and US$250/year of life saved (YLS) compared with no vaccination (Table 1 and Fig. 1). Compared to 20% population vaccine coverage, increasing coverage to 30% would prevent an additional 74 million infections and 208,000 deaths, with an ICER of US$40/infection prevented and US$870/YLS. Increasing coverage from 30% to 40% and from 40% to 50% would prevent an additional 70,000 and 31,000 deaths, and result in ICERs of US$80/infection prevented and US$2,820/YLS and US$150/infection prevented and US$7,240/YLS, respectively. Beyond 50% coverage, reductions in infections and deaths continue, although with diminishing efficiency; the ICER for expanding coverage from 60% to 70% is US$760/infection prevented and US$41,900/YLS.

**Table 1.** Clinical and cost outcomes of the investment into the COVAX AMC initiative. Total population vaccinated, total cost of vaccination, COVID-19 infections, and total YLS are rounded to the nearest thousand. Costs, in US$, are undiscounted since they are an upfront investment. YLS are calculated compared to the 0% vaccine supply strategy and discounted at 3% per year. ICERs are presented as dollars per infection prevented and dollars per YLS and are calculated using unrounded values and then rounded to the nearest ten dollars.

| Modeled vaccine supply* | Total population vaccinated* | Total cost of vaccination (US\$) | COVID-19 infections | COVID-19 deaths | Discounted total YLS | ICER (US\$/infection prevented) | ICER (US\$/YLS) |
| --- | --- | --- | --- | --- | --- | --- | --- |
| 0% | - | - | 547,032,000 | 2,448,000 | - | - | - |
| 20% | 507,685,000 | 6,448,241,000 | 252,978,000 | 512,000 | 25,853,000 | 20 | 250 |
| 30% | 761,472,000 | 9,357,565,000 | 179,094,000 | 305,000 | 29,214,000 | 40 | 870 |
| 40% | 1,015,665,000 | 12,266,889,000 | 144,377,000 | 234,000 | 30,247,000 | 80 | 2,820 |
| 50% | 1,268,506,000 | 15,176,214,000 | 124,397,000 | 204,000 | 30,649,000 | 150 | 7,240 |
| 60% | 1,522,348,000 | 18,085,538,000 | 114,667,000 | 199,000 | 30,735,000 | 300 | 33,750 |
| 70% | 1,776,190,000 | 20,994,863,000 | 110,816,000 | 193,000 | 30,805,000 | 760 | 41,900 |

*Abbreviations: AMC, Advance Market Commitment; YLS, years of life saved; ICER, incremental cost-effectiveness ratio.*
\*Modeled vaccine supply is administered to the indicated proportion of the population of each of the 91 countries (India is excluded, see Methods).

**Figure 1.**
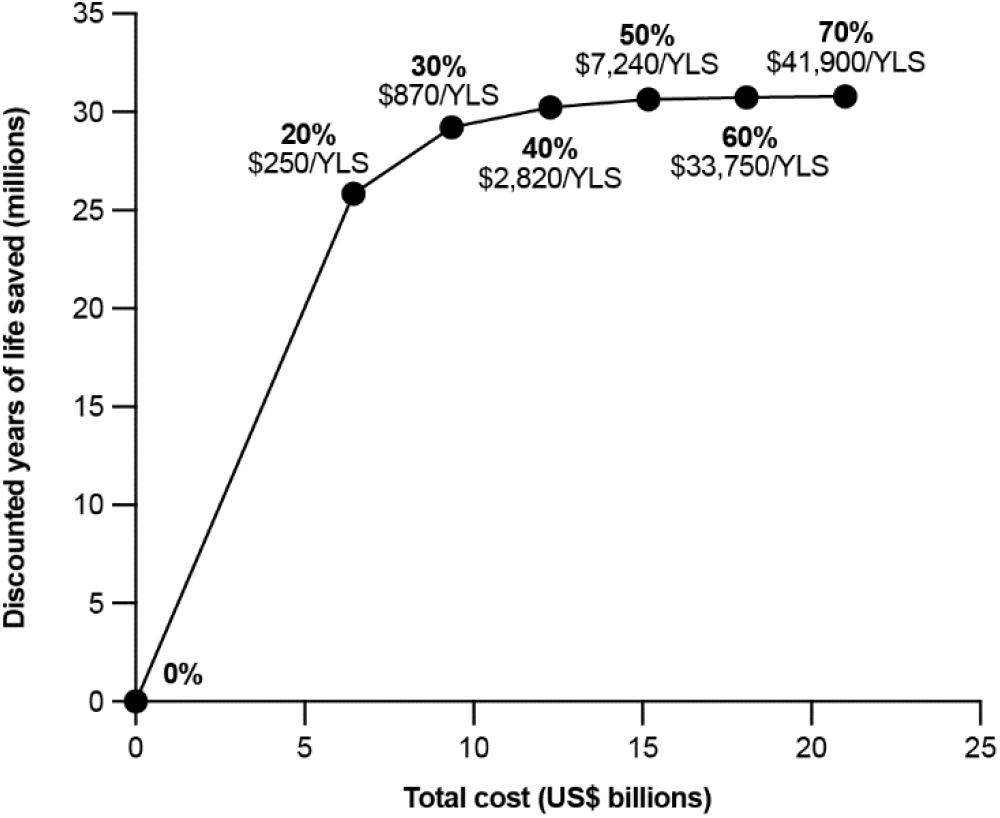
Cost-effectiveness of investment into the COVAX AMC vaccination program for low- and middle-income countries. Modeled outcomes are presented for donor investments into the COVAX Advance Market Commitment (AMC) program. For each vaccine supply strategy, the years of life saved (YLS) compared to the strategy in which no vaccine is plotted against the total cost of the vaccination program. YLS are discounted at 3% per year; costs are undiscounted since they are an upfront investment from the donor perspective. The incremental cost-effectiveness ratio (ICER) of each strategy is represented by the inverse of the slope connecting two points and labeled next to each strategy, rounded to the nearest ten.

### Impacts of vaccine and epidemic traits

In one-way sensitivity analyses, the cost-effectiveness of providing a global vaccination supply to 20% of the population was most affected by the prevalence of prior protective immunity, the infection fatality ratio (IFR) of COVID-19, the epidemic effective reproductive number (R*_e_*), and, to a lesser extent, program costs (Fig. 2). Aside from scenarios in which the population prevalence of prior protective immunity against SARS-CoV-2 infection was 25% or greater, the ICER would remain below US$60/infection prevented and US$1,700/YLS for achieving 20% coverage across a wide range of assumptions (Fig. 2 and Extended Data Tables 2-10). Vaccine efficacy against infection, efficacy against symptomatic disease, and efficacy against severe/critical disease; as well as the pace of vaccination rollout, vaccine uptake, and vaccine cost, had negligible effects on the cost-effectiveness of a 20% vaccine supply to LMICs (Fig. 2). For example, the ICER for a vaccine supply of 20% would be US$500/YLS even with a doubling of program costs (Table 2).

**Figure 2.**
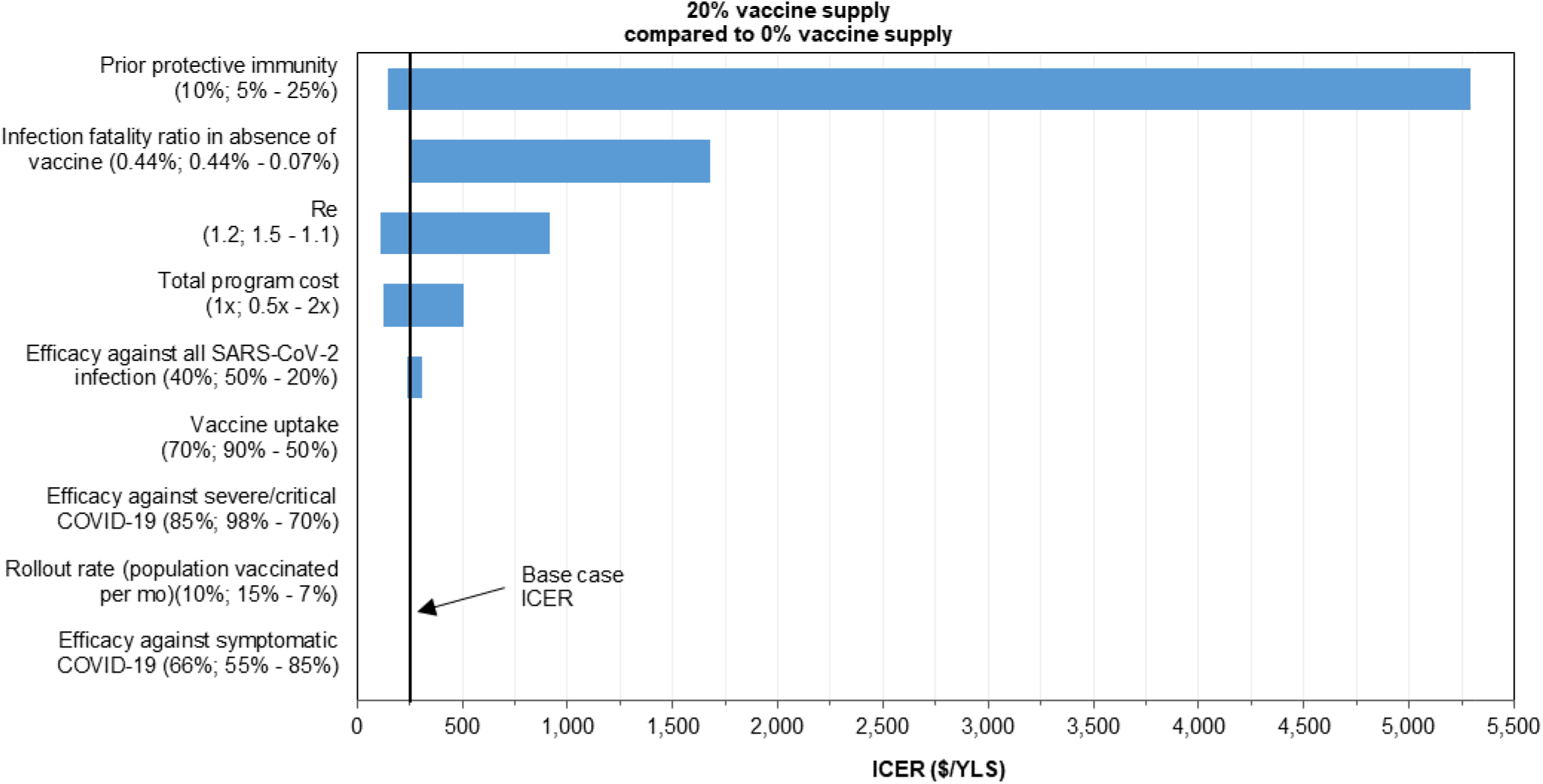
One-way sensitivity analyses: influence of key parameters on the cost-effectiveness of the COVAX AMC program across 9 representative countries. One-way sensitivity analyses were conducted in a subset of 9 representative countries: Afghanistan, Cambodia, Lesotho, Moldova, Mongolia, Morocco, Nicaragua, Sri Lanka, and Zambia. These countries were chosen to reflect variation in global region, age structure, hospital bed capacity, and ICU bed capacity that exists among the 91 COVAX Advance Market Commitment countries (see Methods). The base case incremental cost-effectiveness ratio (ICER, US$/year of life saved) of vaccinating 20% of the population in these 9 countries compared to no vaccination is shown by the black vertical line (US$250/YLS). The range of ICERs when each parameter is varied over a plausible range (base case value; value resulting in lowest ICER – value resulting in highest ICER) is shown in blue horizontal bars. Abbreviations: R*_e_*, effective reproductive number; YLS, year of life saved.

**Table 2.** Cost-effectiveness of the investment into the COVAX AMC program when key parameters are varied in 9 representative countries. One-way sensitivity analyses were conducted in a subset of 9 representative countries: Afghanistan, Cambodia, Lesotho, Moldova, Mongolia, Morocco, Nicaragua, Sri Lanka, and Zambia. These countries were chosen to reflect the variation in global region, age structure, hospital bed capacity, and ICU bed capacity that exists among the 91 COVAX Advance Market Commitment (AMC) countries (Methods). Incremental cost-effectiveness ratios are presented as US$/infection prevented and US$/year of life saved (YLS) and rounded to the nearest ten. Dominated strategies are ones that provide fewer health benefits than a less costly strategy (strong dominance) or have a higher ICER than that of a strategy providing greater health benefits (extended dominance). In incremental scenarios resulting in relatively few additional infections or deaths, strategies with increased vaccine supply may appear to be dominated by those with lower vaccine supply due to stochastic variation. The population-wide infection fatality ratio for the 9 included countries (0.44%) differed from that of all 91 countries (0.45%). Reductions in the infection fatality ratio were modeled by decreasing the risk of developing severe/critical disease to 50% and 25% of the base case risk, resulting in an IFR of 0.18% and 0.07%, respectively. Total cost of the vaccination program included both fixed and variable costs.

| Modeled vaccine supply | $R_e$ | | | Infection fatality ratio in the absence of a vaccine, % | | | Total cost of vaccination program relative to base case | | |
| --- | --- | --- | --- | --- | --- | --- | --- | --- | --- |
|  | 1.1 | 1.2 | 1.5 | 0.07 | 0.18 | 0.44 | 0.5x | 1.0x | 2.0x |
| **Incremental cost-effectiveness ratio (US\$/year of life saved)** | | | | | | | | | |
| 0% | - | - | - | - | - | - | - | - | - |
| 20% | 910 | 250 | 110 | 1,680 | 610 | 250 | 130 | 250 | 500 |
| 30% | 4,570 | 1,040 | 190 | 3,610 | 2,100 | 1,040 | 520 | 1,040 | 2,090 |
| 40% | 16,170 | 3,570 | 440 | 21,730 | 6,760 | 3,570 | 1,790 | 3,570 | 7,140 |
| 50% | dominated | 5,890 | 1,150 | dominated | 18,910 | 5,890 | 2,940 | 5,890 | 11,770 |
| 60% | dominated | 20,080 | 3,840 | 44,360 | 21,490 | 20,080 | 10,040 | 20,080 | 40,170 |
| 70% | 152,780 | dominated | 9,590 | dominated | 130,180 | dominated | dominated | dominated | dominated |
| **Incremental cost-effectiveness ratio (US\$/infection prevented)** | | | | | | | | | |
| 0% | - | - | - | - | - | - | - | - | - |
| 20% | 50 | 20 | dominated | 20 | 20 | 20 | 10 | 20 | 40 |
| 30% | 180 | 40 | 20 | 40 | 40 | 40 | 20 | 40 | 90 |
| 40% | dominated | 80 | 30 | 80 | 80 | 80 | 40 | 80 | 160 |
| 50% | dominated | 130 | 50 | 140 | 130 | 130 | 70 | 130 | 270 |
| 60% | 1,560 | 430 | 100 | 370 | 320 | 430 | 210 | 430 | 860 |
| 70% | 4,850 | 560 | 230 | 530 | 850 | 560 | 280 | 560 | 1,120 |

*Abbreviations: AMC, Advance Market Commitment; $R_e$ , effective reproductive number.*

The projected ranges of ICERs as vaccine supply increased from 20% to 70% while R*_e_*, IFR, and program costs were independently varied are presented in Table 2. For lower levels of R*_e_* and IFR, ICERs would remain below US$5,000/YLS up to 30% coverage (Table 2). Our findings were also robust to assumptions about vaccination costs: if program costs were doubled, ICERs would remain below US$2,500/YLS up to 30% coverage, and below US$8,000/YLS up to 40% coverage (Table 2). The full set of cost and clinical outcomes for all sensitivity analyses conducted (including analyses in which we varied prior protective immunity, IFR, R*_e_*, program costs, vaccine uptake, pace of vaccination rollout, and vaccine efficacy) is shown in Extended Data Tables 2-10.

In two-way sensitivity analyses, we simultaneously varied IFR and program costs and found that the ICER to achieve 20% vaccine coverage would remain at or below US$3,350/YLS, including in a scenario in which the overall IFR was 83% lower than in the base case and program costs doubled (Fig. 3A). In the lowest IFR scenario tested, the ICER for extending the supply from 20% to 30% would be US$7,210/YLS when program costs are doubled, but would otherwise remain at or below US$3,610/YLS at all tested program costs (Fig. 3B). ICERs would range from US$1,790/YLS to US$43,460/YLS when considering expanding vaccine supplies from 30% to 40% (Fig. 3C). We also simultaneously varied R*_e_* and program costs and found that the ICER remained below US$9,140/YLS to achieve up to 30% vaccine supply even with a lower R*_e_* (1.1) and doubling of program costs (Supplementary Figure 2).

**Figure 3.**
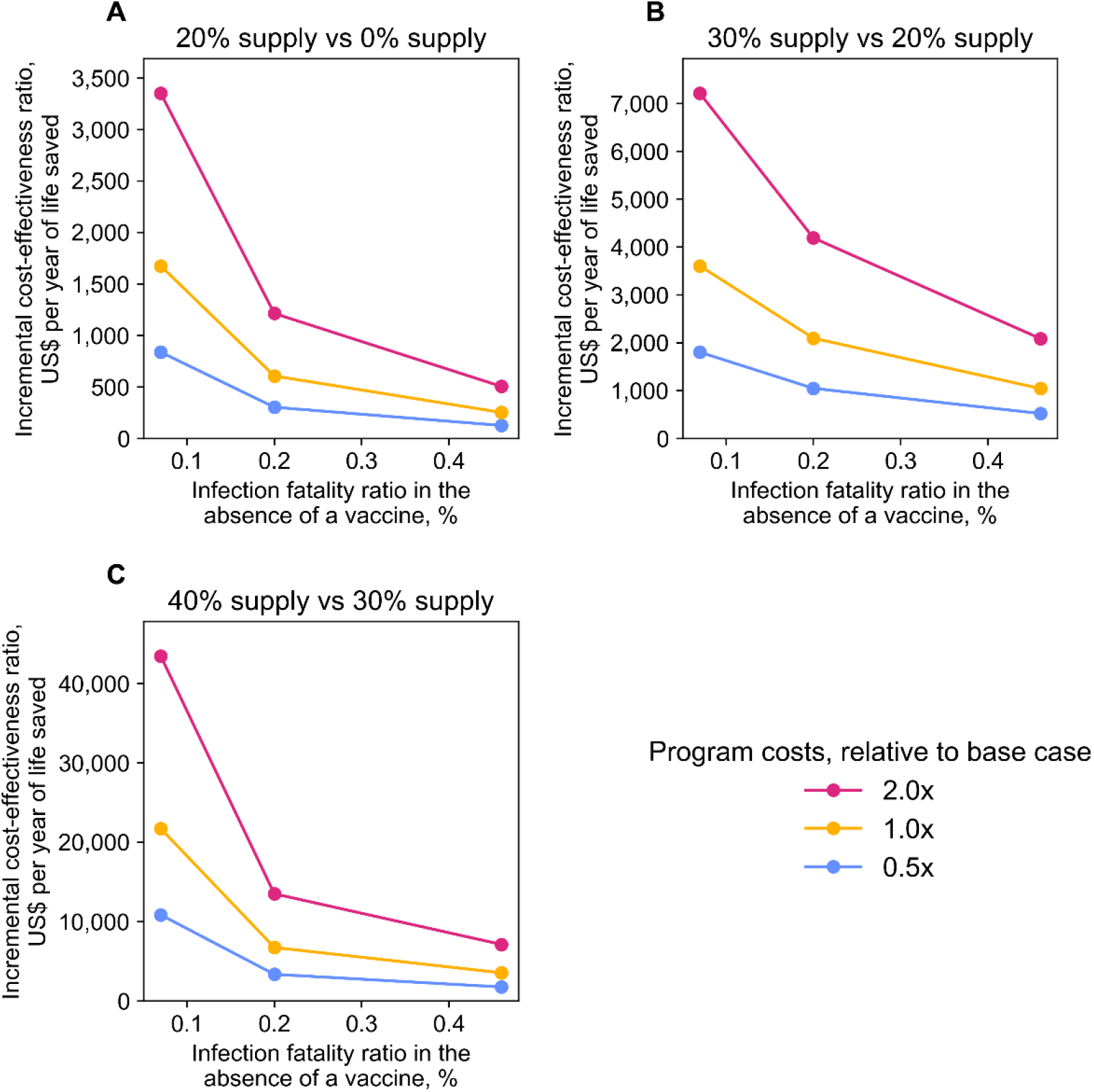
Two-way sensitivity analyses: influence of infection fatality ratio and vaccination cost on cost-effectiveness across 9 representative countries. Two-way sensitivity analyses were conducted in a subset of 9 representative countries: Afghanistan, Cambodia, Lesotho, Moldova, Mongolia, Morocco, Nicaragua, Sri Lanka, and Zambia. These countries reflect variation in global region, age structure, hospital bed capacity, and ICU bed capacity that exists among the 91 COVAX Advance Market Commitment countries (see Methods). In two-way sensitivity analyses, the risk of developing severe/critical disease among those infected with SARS-CoV-2 and the overall cost of the vaccination program were varied concurrently. In the base case scenario, the population-wide infection fatality ratio (IFR) was 0.44% in the absence of a vaccine for the 9 included countries. Decreasing the risk of developing severe/critical disease to 50% and 25% of the base case risk resulted in an IFR of 0.18% and 0.07%, respectively. For each scenario, the IFR in the absence of a vaccine is displayed on the horizontal axis, the incremental cost-effectiveness ratio (ICER) compared to the next-highest supply level is displayed on the vertical axis, and the program cost relative to the base case is denoted by color. The effect of concurrently varying IFR and program cost on the ICER compared to the next-highest supply level is displayed for 20% supply (A), 30% supply (B), and 40% supply (C). The scales of the vertical axes differ in the three plots.

## Discussion

Investments into COVID-19 vaccine supply and distribution to LMICs sufficient to achieve 20% population coverage would prevent 294 million infections and 2 million deaths over a one-year period and be highly cost-effective with an ICER of approximately US$20/infection prevented and US$250/YLS compared to no vaccine coverage. Financing an expanded vaccine supply for up to 50% of these populations would prevent millions of infections and save hundreds of thousands of additional lives, with ICERs below US$200/infection prevented and US$8,000/YLS. These results demonstrate the substantial health benefits and value of global efforts to promptly support LMICs with the infrastructure and supply needed to vaccinate large proportions of their populations, and complement arguments focused on health equity^9^, economic benefits^10^, and pandemic control efforts^11^.

While there is no universally accepted ICER threshold to determine value among donor countries investing on behalf of lower-income countries, the ICERs we estimate for funding of vaccination programs in 91 LMICs are $250/YLS at 20% coverage and below $8,000/YLS at 50% coverage. These ICERs are substantially lower than or comparable to similar and important donor-financed public health measures in LMICs, such as the global delivery of antiretroviral therapy for HIV through the US President’s Emergency Plan for AIDS Relief (PEPFAR)^30,31^. Between 2004-2013, PEPFAR was supported by approximately US$49.8 billion in US government funding and resulted in an estimated 11.6 million years of life saved, for an ICER of approximately US$4,310/YLS (see Supplementary Methods)^30,31^. To put the COVID-19 vaccination program investment into further context, the total estimated cost of funding a 50% vaccine supply to the 91 countries is approximately US$15 billion, which represents approximately 0.3% of the estimated US$5.3 trillion US government domestic investment in the COVID-19 response to date^32^.

Our findings were most sensitive to the prevalence of protective immunity at the time of vaccine rollout. As expected, higher prevalence of prior protective immunity would reduce the vaccination program’s value, due to expenditures on individuals who do not derive benefit in the model. Most LMICs that have conducted population-based estimates have found rates of prior exposure of 4-10%^15,33,34^. Further, our model assumes that prior immunity offers complete protection from re-infection and is durable for the 360-day time horizon, which likely overestimates the protective effects of prior infection and may underestimate the value of vaccination^35–37^.

We also found that effective reproductive numbers lower than our base case assumption (R*_e_*=1.2) would reduce total infections, hospitalizations, and deaths, thereby reducing the value of a vaccination program. In the absence of effective vaccine rollout, however, both the COVID-19 pandemic itself and non-pharmaceutical interventions to limit COVID-19 transmission are expected to continue to have deleterious impacts on both health and the economy in each of these countries, costs that are not reflected in our model^10^. Moreover, the characteristics of COVID-19 surges have been highly variable and unpredictable, and it is likely that, in the absence of vaccines, countries may continue to experience epidemic waves with higher R*_e_*, particularly if and when non-pharmaceutical interventions are relaxed^38^.

Finally, our results were sensitive to assumptions about COVID-19 disease severity. Our base case IFR for the 91 LMICs in the absence of vaccines (0.45%) is derived from published data on the natural history of COVID-19 and calibrated to reflect IFRs in published meta-analyses that include paired population seroprevalence and death reporting data^39^. These estimates are supported by data from South Africa, which includes monitoring of excess natural deaths^40^, and by an autopsy study in Zambia, which found that approximately 16% of deceased individuals were unrecognized as having COVID-19 at death but tested positive for COVID-19 post-mortem^14^. However, IFR estimates in some LMICs based on official death reporting, such as Kenya, have been 20 times lower than our base case value^41^ and there have been sizeable variations in estimated IFR between global regions (e.g. 0.37% in western sub-Sahran Africa versus 1.45% in eastern Europe)^42^. To account for this, we varied the probability of developing severe or critical infection to as low as 25% of the estimates in our base case, resulting in an IFR of 0.07%, or 83% lower than in the base case, and nearly 70% lower than pooled estimates in the sub-Saharan African region^43^. Even in this low IFR scenario, funding 20% vaccination would have an ICER of US$20/infection prevented and US$1,680/YLS. Consequently, although estimates of COVID-19-specific mortality rates remain a matter of debate in LMICs^44,45^, the value of vaccination programs would remain high in most scenarios.

This analysis has several limitations. First, natural history inputs were originally derived and validated as part of an analysis based in South Africa^8^. We used inputs calibrated to data from South Africa for three reasons: 1) accurate IFR data from other countries, particularly many LMICs, is limited; 2) age is well-established as the greatest risk factor for COVID-19 mortality and, after accounting for age, additional co-morbidities appear to have little additional effect on expected and reported mortality in LMICs^46,47^; and 3) use of data from South Africa is likely to more closely reflect SARS-CoV-2 natural history estimates in LMICs than data from high-income countries. Second, our model assumes homogeneous mixing, such that all individuals within a country are equally likely to become infected and transmit to others, and we do not include transmission between countries. The homogenous mixing assumption may underestimate transmissions in high contact and densely populated settings while overestimating transmissions in low contact and rural settings. Not including transmission between countries might also underestimate the value of increased global vaccine distribution. Third, our model includes data on vaccine efficacy, hesitancy, and costs, which are all from published studies but subject to uncertainty. Despite this, our findings were robust to plausible ranges in these parameters. Fourth, we did not account for the potential secondary health benefits of COVID-19 vaccination. The pandemic has been predicted to indirectly increase morbidity and mortality in LMICs through the overwhelming of health systems, worsening of food insecurity, disruption of supply chains, infections of health care workers, and repurposing of healthcare sector budgets^20,48,49^. We also did not account for potential longer term secondary benefits of vaccination programs, such as prevention of the emergence of viral variants and strengthening public health infrastructure in LMICs. Since this analysis is from the donor perspective, we did not account for averted domestic healthcare costs due to fewer COVID-19 hospitalizations that would be borne by recipient countries in the absence of vaccination. Finally, we do not model the potential economic losses from failure to accomplish global vaccination, losses estimated up to US$9 trillion, as much as half of which are expected to be borne by high-income countries^10^.

In summary, we found that donor investments in the COVAX initiative to fully subsidize 20% vaccine coverage in LMICs, would prevent nearly 300 million infections and 2 million deaths over one year, and would be cost-effective compared to current widely-supported public health and medical interventions in the US^30,31^. Attaining coverage levels up to 50% would provide major additional benefits and remain cost-effective at thresholds below those of other donor aid programs for health. These findings, in conjunction with ethical, social, and economic benefits of global vaccine equity, support urgent global efforts to promote vaccine distribution and implementation of vaccination programs in LMICs.

## Data Availability

There are no original data generated as part of this study. All data are derived from published or publicly available data and referenced as such. Code data will be made available to any requesting party by contacting the corresponding author.

## METHODS

### Analytic Overview

The Clinical and Economic Analysis of COVID-19 interventions (CEACOV) model is a validated, dynamic microsimulation of the natural history of COVID-19 and the impact of public health interventions^8,25–27^. We used the CEACOV model to project the clinical impact, cost, and cost-effectiveness of donor outlays to purchase, distribute, and deploy SARS-CoV-2 vaccines to 91 low- and middle-income countries (LMICs). We excluded India from our base case analysis, despite India being named in the original COVAX plan, because of its plan to produce vaccines domestically^24^. We modeled discrete epidemics in each country using country-specific age distribution, population size, and hospital and ICU bed capacity (Supplementary Table 1); we present both country-level and aggregate outcomes.

### Model Structure

#### Disease states and progression

The CEACOV model is based on an SEIR framework, and includes susceptible, exposed, infectious, recovered, and dead states^50^ (Supplementary Methods and Supplementary Fig. 1). Susceptible individuals face a daily probability of infection with SARS-CoV-2, while infected individuals face daily probabilities of disease progression through six COVID-19 states: pre-infectious latency, asymptomatic, mild/moderate, severe, critical, and recuperation (Supplementary Table 2). With mild/moderate disease, individuals have symptoms, such as cough or fever, but do not require inpatient management. With severe disease, symptoms warrant inpatient management, and with critical disease, patients require ICU care to survive. Recovered individuals are assumed immune from repeat infection for the duration of the modeled time horizon^51^. The model considers three broad age bands: 0-19, 20-59, and ≥60 years. Mortality from COVID-19 is dependent on age and availability of hospital and ICU beds (Supplementary Table 2).

#### Transmission

Individuals with SARS-CoV-2 infection transmit to susceptible individuals at health state-stratified rates (Extended Data Table 1). All susceptible people face equal probabilities of contacting infected individuals and becoming infected (i.e., homogenous mixing). The number of projected infections depends on daily prevalence of active disease and daily proportion of the population susceptible to infection; as well as time-invariant transmission rates, calibrated to achieve the base case effective reproductive number (R*_e_*)–the average number of transmissions per infection at simulation onset. Nonpharmaceutical interventions, such as mask mandates and physical distancing, are not explicitly modeled; however, the uptake and effectiveness of these policies are reflected in the value of the effective reproductive number.

#### Years of life lost from COVID-19

We quantified the clinical benefit of a vaccination program in terms of years of life saved from prevented COVID-19 deaths. Life-years occurring in future periods were discounted at a rate of 3%/year—as commonly done in cost-effectiveness analyses of healthcare interventions^52^. For each country included in our analysis, we calculated the average number of years a person would have lived subsequent to his or her current age had he or she not died of COVID-19 using sex-stratified life tables published by the United Nations World Population Prospects for the period 2015-2020^53^. We then calculated discounted years of life lost (dYLL) using the methodology employed by the Global Burden of Disease Project^54^:

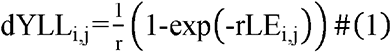

where *r* represents the chosen discount rate and LE_i,j_ represents the average remaining life expectancy among those age i with sex j. Since our model uses large age intervals (0-19, 20-59, ≥60 years) and does not distinguish individuals by sex, we calculated the average years of life lost per COVID-19 death in each of our model’s age brackets 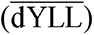 by taking a weighted average of the values calculated using equation (1) over the age-sex distribution of deaths within each large age interval:

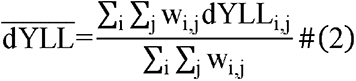

where w_i,j_ represents the relative frequency at which someone age i and sex j is expected to die of COVID-19. We calculated this frequency based on the assumed age-sex distribution of infections (a_i,j_) and the age- and sex-specific infection fatality ratios (IFR_i,j_) reported by O’Driscoll *et al.*^39^.

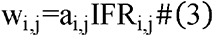

Because we were unable to identify age- and sex-stratified data on the reported number of infections in most of the countries included in our analysis, we assumed that the age-sex distribution of infections would mirror the overall population structure of a given country (i.e., homogeneous mixing).

#### Vaccine supply

We modeled the impact of a COVAX program providing vaccines at various coverage levels–from 20% to 70% of each country’s population. The vaccine would first be given to those aged ≥60 years, regardless of history of COVID-19^55^. If additional doses were available, they were given to those aged 20-59 years, and then to those aged <20 years (trials ongoing)^56–58^.

#### Vaccine efficacy against infection, disease, and severe disease

To reconcile the various endpoints used in vaccine clinical trials, we incorporated three measures of vaccine efficacy into our model: efficacy in preventing SARS-CoV-2 infection (VE_1_) efficacy in preventing symptomatic COVID-19 disease (VE_2_), and efficacy in preventing severe or critical COVID-19 disease (VE_3_). In the context of our model, we defined symptomatic COVID-19 disease as any infected health state other than pre-infectious incubation and asymptomatic states. Severe or critical COVID-19 disease was defined as any illness that would require hospitalization and corresponds to the severe and critical health states in the model.

We modeled four mutually exclusive outcomes of vaccination: protection from SARS-CoV-2 infection (Outcome A, also referred to as full immunity), protection from symptomatic COVID-19 disease but not infection (Outcome B), protection from severe or critical COVID-19 disease but not symptomatic disease (Outcome C), and no protection (Outcome D, also referred to as full susceptibility). The probability of each outcome among those vaccinated was determined based on vaccine efficacy measures as follows:

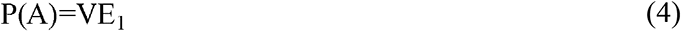

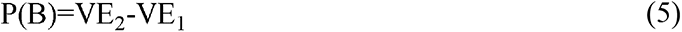

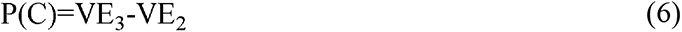

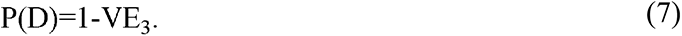

Equations (4)-(7) are subject to the constraint that VE_3_ must be greater than VE_2_, which itself must be greater than VE_1_. Although it is possible to construct situations that would violate this constraint (for example, a vaccine whose reductions in infections are offset by an increase in disease severity among those vaccinated), we believe such scenarios are extremely unlikely in light of data on existing COVID-19 vaccines^1–4^.

Those with full immunity (Outcome A) can neither become infected with SARS-CoV-2 nor transmit the virus to others. Those who are protected from symptomatic disease but not infection (Outcome B) can become infected in the model but will not display symptoms of illness. Similarly, those who are protected from severe or critical disease but not symptomatic disease (Outcome C) are able to develop mild/moderate symptoms of COVID-19 but will not transition to the severe (hospitalized) or critical (ICU) states. Finally, those without an effective response to vaccination (Outcome D) remain fully susceptible to infection and do not experience any reduction in the probability of severe or critical disease. Protection from infection or symptomatic disease as a result of vaccination or prior infection was assumed to last throughout the duration of the 360-day time horizon.

#### Resource use, costs, and cost-effectiveness

The model tallies resource utilization, including hospital and ICU admissions for those with severe or critical disease, accounting for country-specific capacity constraints. Hospitalization is provided for those with severe illness, and ICU care, if available, is provided for those with critical illness. Costs are from a COVAX payer perspective, and thus include only vaccination program costs that would be funded by the COVAX AMC. These include program infrastructure costs as well as cost per vaccine dose delivered^28^. Vaccination program costs occur within the 360-day time horizon and are not discounted. The incremental cost-effectiveness ratios (ICERs) of program strategies corresponding to different levels of population coverage are calculated by dividing the change in costs by the change in benefits (years of life saved or infections prevented) compared to the next least-expensive non-dominated strategy^59^. A strategy is considered “dominated” if it is more expensive and less efficacious (resulting in more years of life lost) than an alternative strategy, or if it is less economically efficient (resulting in a higher ICER) than a more efficacious strategy. In general, dominated strategies are considered unfavorable from a decision-making perspective, and thus are not included when calculating ICERs for the remaining non-dominated strategies.

### Input Parameters

#### Country characteristics

For each of the 91 countries, we grouped age-stratified population data from the United Nations 2019 World Population Prospects^53^ to calculate the proportion of the population in each of the three modeled age strata: 0-19, 20-59, and ≥60 years (Supplementary Table 1). Hospital and ICU bed capacity for each country were derived from data published by the World Health Organization, the World Bank, and country-level health agencies, as well as from peer-reviewed literature (Supplementary Table 1).

#### Disease progression and transmission dynamics

The average duration of each COVID-19 state varies by severity and was derived from studies describing the clinical characteristics of COVID-19 cases in China and the US (Supplementary Methods and Supplementary Table 2). The probability of developing severe or critical disease, as well as mortality, increases with age^60,61^. Transmission rates are highest for individuals in asymptomatic and mild/moderate states, whereas individuals in severe and critical states have fewer infectious contacts due to hospitalization or being homebound (Extended Data Table 1)^61–64^. In the base case, the R*_e_* was 1.2 without vaccination. At the start of the simulation, we assumed 10% prevalence of protective immunity against SARS-CoV-2 (based on prior infection)^33,65,66^, and 0.1% initial prevalence of current infection (the global number of reported cases per capita during the first 10 days of April 2021–the length of time post-symptom onset after which replication-competent virus is unlikely)^12,67^. The distribution of infected individuals across disease states at simulation onset is a function of daily state-transition probabilities, and was determined by conducting initialization runs. We varied R*_e_* and prevalence of protective immunity in sensitivity analyses.

#### Vaccine efficacy against infection, symptomatic disease, and severe/critical disease

In the base case analysis, we modeled vaccine efficacy on the Johnson & Johnson (J&J)/Janssen Ad26.COV2.S vaccine trial data^1^ which reported 66.1% efficacy against symptomatic infection (VE_2_) and 85.4% efficacy against severe or critical disease (VE_3_), both among all participants with onset at least 28 days after immunization. These data included outcomes for people of varied ages (median: 52 years; range: 18-100 years) and geographic location (40.9% from Latin America, 44.1% from the US, and 15.0% from South Africa). Given the 66.1% efficacy against symptomatic infection and the conditions laid out above, we assumed 40% efficacy in preventing SARS-CoV-2 infection (VE_1_). We believe this is a conservative estimate given that data from the J&J/Janssen phase III clinical trial suggest an efficacy of 74.2% against asymptomatic infection after day 29 post-vaccination in a limited sample of study participants^68^.

#### Vaccination costs

We derived the cost of the vaccine program using the COVAX Working Group’s February 2021 updated delivery cost estimates^28^ and a review of negotiated prices for COVID-19 vaccines^29^.

The COVAX Working Group estimated the costs of delivering sufficient vaccine for 9% of India’s population and 20% of the remaining 91 AMC countries’ population: a total of 546.3 million people. The estimated upfront cost of vaccine delivery was US$576.4 million and included costs attributed to planning and coordination, training, social mobilization, cold chain equipment, pharmacovigilance, and hand hygiene. To this, we added the estimated US$98.5 million in global and regional level costs (for innovations, post-introduction evaluations, and additional pharmacovigilance) anticipated over a 3-year period. Given that our analysis focuses on an investment from the donor countries’ perspective, these global and regional level costs were treated as an upfront investment despite these costs potentially being utilized over a 3-year period. Our estimated total fixed costs to vaccinate 546.3 million people was US$674.9 million, or US$1.24 per person vaccinated. Since our analysis excluded India, we re-scaled the fixed costs to a population of 507.7 million (20% of the population in the 91 included AMC countries) under the assumption that fixed costs are evenly distributed on a *per capita* basis. For the 91 included AMC countries, this resulted in a total fixed cost of US$630 million for strategies in which 20%-70% of the population is vaccinated (Extended Data Table 1).

We assumed that each vaccine dose delivered would incur costs associated with cold chain, logistics, storage, waste, and transportation. The estimated variable costs of administering vaccines to 546.3 million people totaled US$1.1 billion for delivery (including cold chain recurrent materials, vaccination certificates, personal protective equipment, hand hygiene, vaccine transport, waste management, per diem for outreach, and transportation for outreach) and US$198.3 million for technical assistance–resulting in a total of US$1.3 billion^28^, or US$2.46 per person vaccinated (Extended Data Table 1).

Finally, in our base case, we included a per-dose vaccine cost of US$9, based on the negotiated prices of the J&J/Janssen vaccine (US$8.50 by the European Union, US$10 by the United States, and US$10 by the African Union)^29^. UNICEF and Gavi also announced a negotiated price for the Novavax NVX-CoV2373 and AstraZeneca ChAdOx1 nCoV-19 vaccines of US$6 per vaccine course (US$3 per dose) for COVAX-eligible countries^69^. In our base analysis, we chose the US$9 per vaccine course estimate because it is more conservative. However, we adjusted the total program costs to be 50% and 200% of the base case value in sensitivity analyses to account for the uncertainty in delivery costs and variability in per-unit vaccine costs.

### Sensitivity Analyses

#### Selection of representative countries for sensitivity analyses

To evaluate the influence of key parameters on measures of cost-effectiveness, we performed sensitivity analyses on a subset of representative countries. First, we partitioned our dataset of 91 COVAX AMC-eligible economies (excluding India) into five groups based on age structure, hospital beds *per capita*, and ICU beds *per capita* using *k*-means clustering. Next, we selected 1-2 countries from each group to create a subset of 9 countries representing the range of “country domains” observed in our dataset: Afghanistan, Cambodia, Lesotho, Moldova, Mongolia, Morocco, Nicaragua, Sri Lanka, and Zambia (combined population: 148 million). Under the base case scenario, we were able to reproduce the ICERs of a global vaccination program targeting 91 AMC countries with reasonable accuracy while only considering the costs and benefits (years of life saved) accrued within the chosen subset of 9 countries. The costs of program startup, vaccine purchase, and delivery were scaled to reflect the combined population of these 9 countries. For the 20-40% coverage levels, the mean absolute percentage error (MAPE) between the ICERs calculated using a 9-country approximation and the ICERs calculated using the full dataset of 91 countries was 16% and rose to 30% for the 50-60% coverage levels. The 70% coverage level was found to be dominated when a 9-country approximation was used to calculate ICERs; in contrast, this coverage level was not found to be dominated when the full dataset of 91 countries was used. This suggests that program cost-effectiveness may be reasonably approximated from a subset of countries for coverage levels less than 70%.

#### Parameters varied in one- and two-way sensitivity analyses

In one-way sensitivity analyses, we varied multiple parameters: speed of vaccine rollout; vaccine uptake; vaccine efficacy in preventing SARS-CoV-2 infection, any symptomatic COVID-19, and severe or critical COVID-19 disease requiring hospitalization; baseline probability of developing severe/critical disease in the absence of vaccines among those infected (and resulting IFR); prevalence of prior protective immunity; mean epidemic R*_e_*; and total vaccination program costs.

Variations in vaccine efficacy and costs were ranged to include estimates that correspond to the J&J/Janssen Ad26.COV2.S, Pfizer-BioNTech BNT162b2, and AstraZeneca ChAdOx1 nCoV-19 vaccines^1,3,70^, vaccines that are currently being distributed as part of the COVAX-AMC program or are included in future distribution forecasts^71,72^. In a two-way sensitivity analysis, we simultaneously varied the effective IFR in the absence of vaccine and vaccination program costs.

## Data and code availability statement

There are no original data generated as part of this study. All data are derived from published or publically available data and referenced as such. Code data will be made available to any requesting party by contacting the corresponding author.

## ACKNOWLEDGEMENTS

This study was supported by the National Institutes of Health (R37 AI058736-16S1). The findings and conclusions in this report are those of the authors and do not necessarily represent the official views of the US National Institutes of Health. We thank Pooyan Kazemian and Christopher Panella for assistance in developing and coding the model.

## AUTHOR CONTRIBUTIONS

M.J.S. and K.A.F. conceived of the project. C.A., K.P.F., R.G., J.A.S., and F.S. contributed to data acquisition and analysis. M.J.S., K.A.F., A.C., K.P.R., C.A., K.P.F., and J.A.S. contributed to data interpretation. M.J.S. wrote the first draft of the manuscript. All authors contributed to significant revisions of the manuscript and approved the final version.

## ADDITIONAL INFORMATION

Supplementary Information is available for this paper. Correspondence and reqeuests for materials should be addressed to Mark Siedner at. Reprints and permissions information is available at www.nature.com/reprints.

## EXTENDED DATA FIGURE/TABLE LEGENDS

**Extended Data Table 1.** Input parameters for an analysis of COVID-19 vaccination strategies in COVAX AMC-eligible economies.

| Parameter | Base case value (Range) | Source(s) |
| --- | --- | --- |
| **Initial state distributions** |  |  |
| Susceptible | 89.9 (74.9-94.9) |  |
| Infected | 0.1 | 12,67 |
| Recovered | 10.0 (5.0-25.0) | 33,65,66 |
| **Transmission dynamics** |  |  |
| $R_e$ | 1.2 (1.1-1.5) | Assumption |
| Rate of onward transmission by health state<sup>a</sup>, transmissions per person-day* |  | 61–64,73 |
| Asymptomatic | 0.1239 |  |
| Mild or moderate disease | 0.1008 |  |
| Severe disease | 0.0070 |  |
| Critical disease | 0.0056 |  |
| Recuperation after critical disease | 0.0070 |  |
| **Vaccine specifications** |  |  |
| Proportion of population vaccinated per month, % | 10 (7-15) | Assumption |
| Uptake, % of population accepting vaccine | 70 (55-90) | 74 |
| Efficacy in preventing SARS-CoV-2 infection, % | 40 (20-50) | Assumption |
| Efficacy in preventing symptomatic COVID-19, % | 66 (55-85) | 1,3 |
| Efficacy in preventing severe/critical COVID-19, % | 85 (70-98) | 1,70 |
| **Costs of vaccine purchase and delivery** |  |  |
| Fixed costs, US\$ | 629,592,000 | 28 |
| Variable costs, US\$ per person vaccinated | | |
| Variable delivery costs | 2.46 | 28 |
| Vaccine purchase | 9.00 | 29 |

**Abbreviations:** *AMC*, Advance Market Commitment; $R_e$ , effective reproductive number.
\* Corresponds to $R_e$ of 1.2

**Extended Data Table 2.**
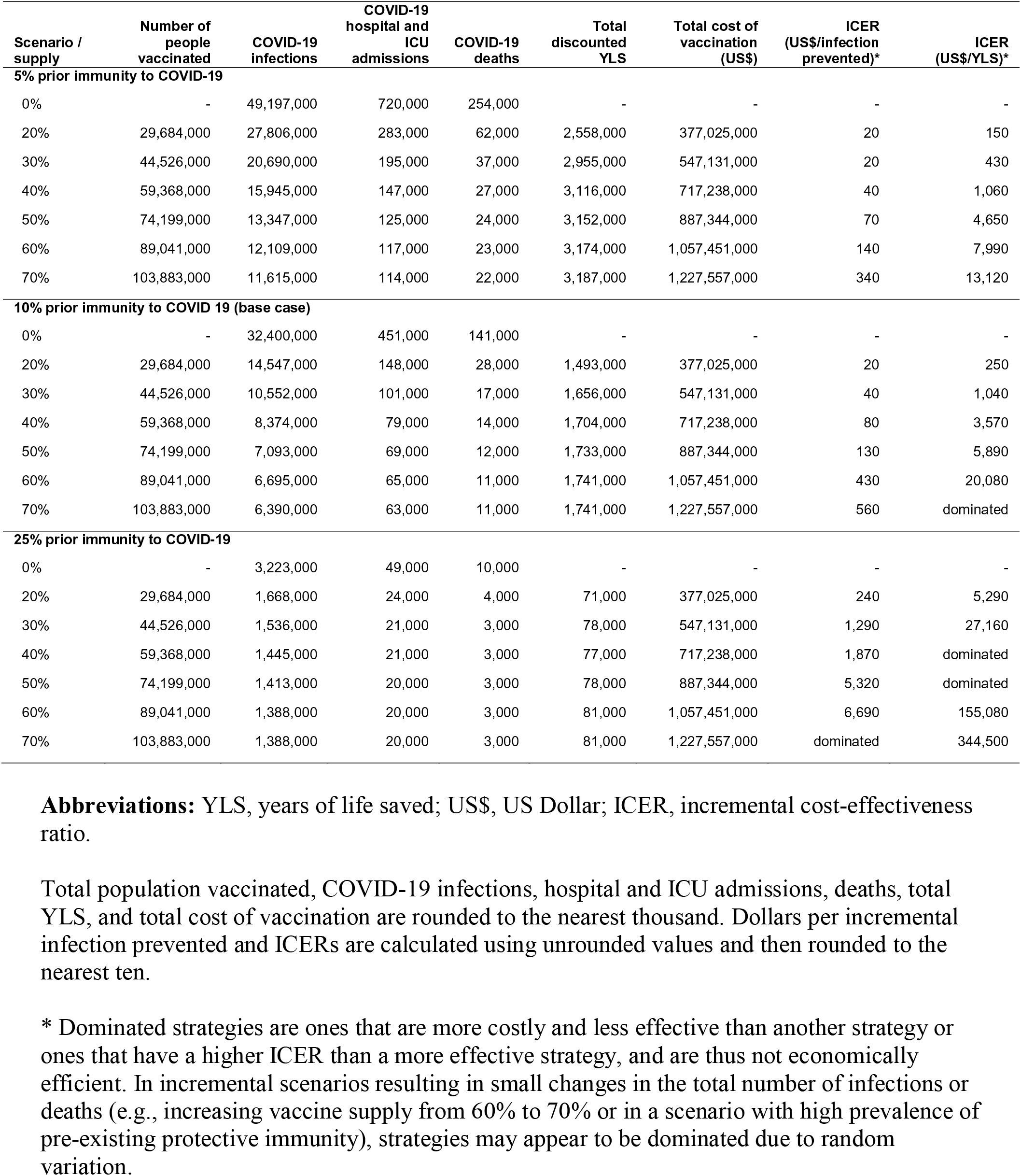
One-way sensitivity analyses: influence of prior immunity to COVID-19 on clinical and economic outcomes across 9 representative countries.

| Scenario / supply | Number of people vaccinated | COVID-19 infections | COVID-19 hospital and ICU admissions | COVID-19 deaths | Total discounted YLS | Total cost of vaccination (US\$) | ICER (US\$/infection prevented)* | ICER (US\$/YLS)* |
| --- | --- | --- | --- | --- | --- | --- | --- | --- |
| **5% prior immunity to COVID-19** |  |  |  |  |  |  |  |  |
| 0% | - | 49,197,000 | 720,000 | 254,000 | - | - | - | - |
| 20% | 29,684,000 | 27,806,000 | 283,000 | 62,000 | 2,558,000 | 377,025,000 | 20 | 150 |
| 30% | 44,526,000 | 20,690,000 | 195,000 | 37,000 | 2,955,000 | 547,131,000 | 20 | 430 |
| 40% | 59,368,000 | 15,945,000 | 147,000 | 27,000 | 3,116,000 | 717,238,000 | 40 | 1,060 |
| 50% | 74,199,000 | 13,347,000 | 125,000 | 24,000 | 3,152,000 | 887,344,000 | 70 | 4,650 |
| 60% | 89,041,000 | 12,109,000 | 117,000 | 23,000 | 3,174,000 | 1,057,451,000 | 140 | 7,990 |
| 70% | 103,883,000 | 11,615,000 | 114,000 | 22,000 | 3,187,000 | 1,227,557,000 | 340 | 13,120 |
| **10% prior immunity to COVID 19 (base case)** |  |  |  |  |  |  |  |  |
| 0% | - | 32,400,000 | 451,000 | 141,000 | - | - | - | - |
| 20% | 29,684,000 | 14,547,000 | 148,000 | 28,000 | 1,493,000 | 377,025,000 | 20 | 250 |
| 30% | 44,526,000 | 10,552,000 | 101,000 | 17,000 | 1,656,000 | 547,131,000 | 40 | 1,040 |
| 40% | 59,368,000 | 8,374,000 | 79,000 | 14,000 | 1,704,000 | 717,238,000 | 80 | 3,570 |
| 50% | 74,199,000 | 7,093,000 | 69,000 | 12,000 | 1,733,000 | 887,344,000 | 130 | 5,890 |
| 60% | 89,041,000 | 6,695,000 | 65,000 | 11,000 | 1,741,000 | 1,057,451,000 | 430 | 20,080 |
| 70% | 103,883,000 | 6,390,000 | 63,000 | 11,000 | 1,741,000 | 1,227,557,000 | 560 | dominated |
| **25% prior immunity to COVID-19** |  |  |  |  |  |  |  |  |
| 0% | - | 3,223,000 | 49,000 | 10,000 | - | - | - | - |
| 20% | 29,684,000 | 1,668,000 | 24,000 | 4,000 | 71,000 | 377,025,000 | 240 | 5,290 |
| 30% | 44,526,000 | 1,536,000 | 21,000 | 3,000 | 78,000 | 547,131,000 | 1,290 | 27,160 |
| 40% | 59,368,000 | 1,445,000 | 21,000 | 3,000 | 77,000 | 717,238,000 | 1,870 | dominated |
| 50% | 74,199,000 | 1,413,000 | 20,000 | 3,000 | 78,000 | 887,344,000 | 5,320 | dominated |
| 60% | 89,041,000 | 1,388,000 | 20,000 | 3,000 | 81,000 | 1,057,451,000 | 6,690 | 155,080 |
| 70% | 103,883,000 | 1,388,000 | 20,000 | 3,000 | 81,000 | 1,227,557,000 | dominated | 344,500 |

**Abbreviations:** YLS, years of life saved; US\$, US Dollar; ICER, incremental cost-effectiveness ratio.
Total population vaccinated, COVID-19 infections, hospital and ICU admissions, deaths, total YLS, and total cost of vaccination are rounded to the nearest thousand. Dollars per incremental infection prevented and ICERs are calculated using unrounded values and then rounded to the nearest ten.
\* Dominated strategies are ones that are more costly and less effective than another strategy or ones that have a higher ICER than a more effective strategy, and are thus not economically efficient. In incremental scenarios resulting in small changes in the total number of infections or deaths (e.g., increasing vaccine supply from 60% to 70% or in a scenario with high prevalence of pre-existing protective immunity), strategies may appear to be dominated due to random variation.

**Extended Data Table 3.** One-way sensitivity analyses: influence of risk of infection fatality ratio on clinical and economic outcomes across 9 representative countries.

| Scenario / supply | Number of people vaccinated | COVID-19 infections | COVID-19 hospital and ICU admissions | COVID-19 deaths | Total YLS | Total cost of vaccination (US\$) | ICER (US\$/infection prevented)* | ICER (US\$/YLS)* |
| --- | --- | --- | --- | --- | --- | --- | --- | --- |
| **Infection fatality ratio 0.07% in the absence of a vaccine** |  |  |  |  |  |  |  |  |
| 0% | - | 33,316,000 | 116,000 | 25,000 | - | - | - | - |
| 20% | 29,684,000 | 15,018,000 | 39,000 | 6,000 | 225,000 | 377,025,000 | 20 | 1,680 |
| 30% | 44,526,000 | 10,585,000 | 25,000 | 3,000 | 272,000 | 547,131,000 | 40 | 3,610 |
| 40% | 59,368,000 | 8,423,000 | 21,000 | 3,000 | 280,000 | 717,238,000 | 80 | 21,730 |
| 50% | 74,199,000 | 7,164,000 | 18,000 | 3,000 | 279,000 | 887,344,000 | 140 | dominated |
| 60% | 89,041,000 | 6,705,000 | 17,000 | 2,000 | 288,000 | 1,057,451,000 | 370 | 44,360 |
| 70% | 103,883,000 | 6,386,000 | 17,000 | 2,000 | 287,000 | 1,227,557,000 | 530 | dominated |
| **Infection fatality ratio 0.18% in the absence of a vaccine** |  |  |  |  |  |  |  |  |
| 0% | - | 33,327,000 | 239,000 | 61,000 | - | - | - | - |
| 20% | 29,684,000 | 15,502,000 | 79,000 | 13,000 | 621,000 | 377,025,000 | 20 | 610 |
| 30% | 44,526,000 | 10,710,000 | 53,000 | 8,000 | 702,000 | 547,131,000 | 40 | 2,100 |
| 40% | 59,368,000 | 8,607,000 | 41,000 | 6,000 | 728,000 | 717,238,000 | 80 | 6,760 |
| 50% | 74,199,000 | 7,317,000 | 36,000 | 5,000 | 737,000 | 887,344,000 | 130 | 18,910 |
| 60% | 89,041,000 | 6,780,000 | 33,000 | 5,000 | 745,000 | 1,057,451,000 | 320 | 21,490 |
| 70% | 103,883,000 | 6,579,000 | 33,000 | 5,000 | 746,000 | 1,227,557,000 | 850 | 130,180 |
| **Infection fatality ratio 0.44% in the absence of a vaccine (base case)** |  |  |  |  |  |  |  |  |
| 0% | - | 32,400,000 | 451,000 | 141,000 | - | - | - | - |
| 20% | 29,684,000 | 14,547,000 | 148,000 | 28,000 | 1,493,000 | 377,025,000 | 20 | 250 |
| 30% | 44,526,000 | 10,552,000 | 101,000 | 17,000 | 1,656,000 | 547,131,000 | 40 | 1,040 |
| 40% | 59,368,000 | 8,374,000 | 79,000 | 14,000 | 1,704,000 | 717,238,000 | 80 | 3,570 |
| 50% | 74,199,000 | 7,093,000 | 69,000 | 12,000 | 1,733,000 | 887,344,000 | 130 | 5,890 |
| 60% | 89,041,000 | 6,695,000 | 65,000 | 11,000 | 1,741,000 | 1,057,451,000 | 430 | 20,080 |
| 70% | 103,883,000 | 6,390,000 | 63,000 | 11,000 | 1,741,000 | 1,227,557,000 | 560 | dominated |

**Abbreviations:** YLS, years of life saved; US\$, US Dollar; ICER, incremental cost-effectiveness ratio.

**Extended Data Table 4.**
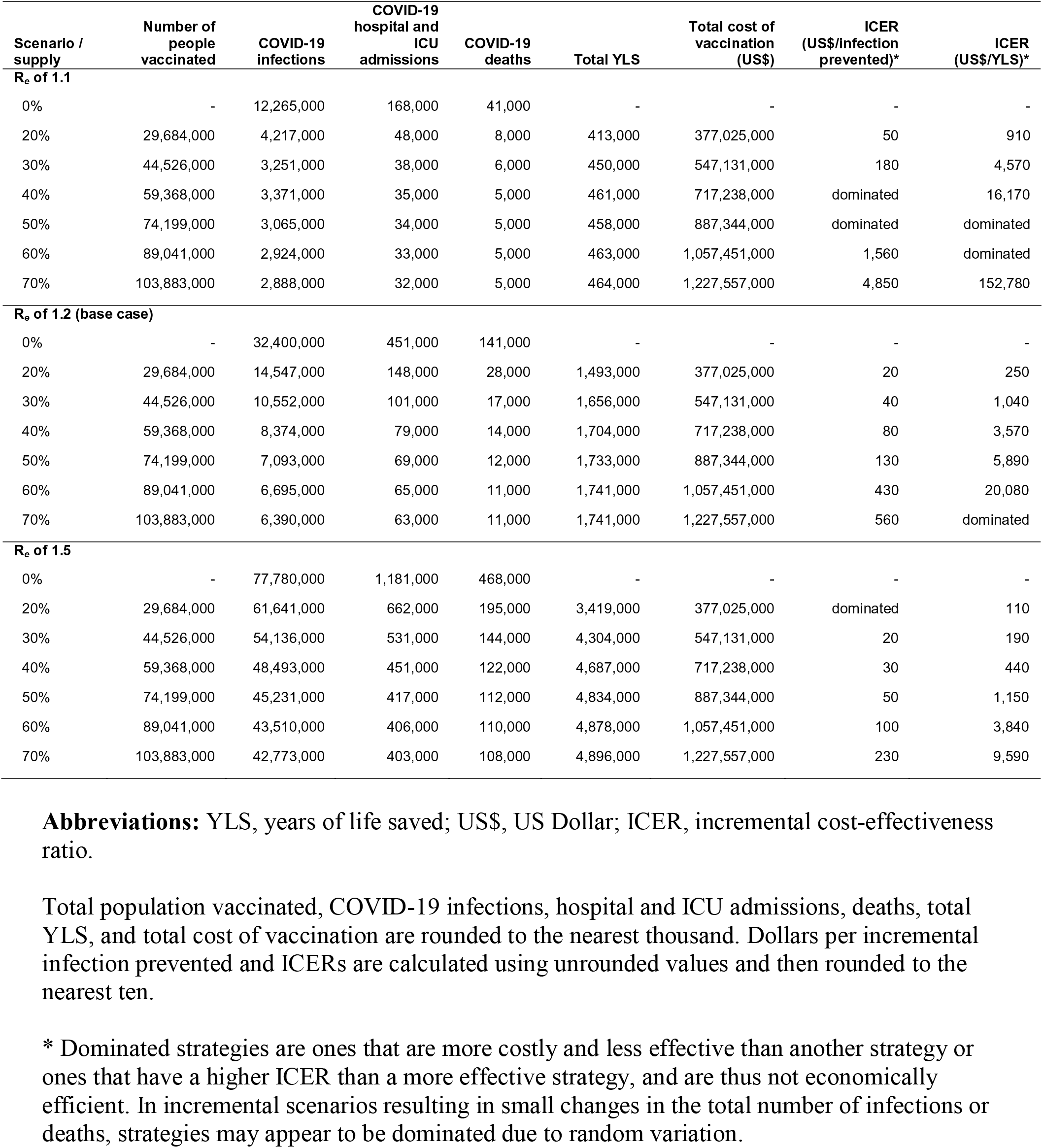
One-way sensitivity analyses: influence of R*_e_* on clinical and economic outcomes across 9 representative countries.

**Extended Data Table 5.** One-way sensitivity analyses: influence of total vaccination program cost on clinical and economic outcomes across 9 representative countries.

| Scenario / supply | Number of people vaccinated | COVID-19 infections | COVID-19 hospital and ICU admissions | COVID-19 deaths | Total discounted YLS | Total cost of vaccination (US\$) | ICER (US\$/infection prevented)* | ICER (US\$/YLS)* |
| --- | --- | --- | --- | --- | --- | --- | --- | --- |
| **0.5x total cost of vaccination program** |  |  |  |  |  |  |  |  |
| 0% | - | 32,400,000 | 451,000 | 141,000 | - | - | - | - |
| 20% | 29,684,000 | 14,547,000 | 148,000 | 28,000 | 1,493,000 | 188,512,000 | 10 | 130 |
| 30% | 44,526,000 | 10,552,000 | 101,000 | 17,000 | 1,656,000 | 273,566,000 | 20 | 520 |
| 40% | 59,368,000 | 8,374,000 | 79,000 | 14,000 | 1,704,000 | 358,619,000 | 40 | 1,790 |
| 50% | 74,199,000 | 7,093,000 | 69,000 | 12,000 | 1,733,000 | 443,672,000 | 70 | 2,940 |
| 60% | 89,041,000 | 6,695,000 | 65,000 | 11,000 | 1,741,000 | 528,725,000 | 210 | 10,040 |
| 70% | 103,883,000 | 6,390,000 | 63,000 | 11,000 | 1,741,000 | 613,779,000 | 280 | dominated |
| **1.0x total cost of vaccination program (base case)** |  |  |  |  |  |  |  |  |
| 0% | - | 32,400,000 | 451,000 | 141,000 | - | - | - | - |
| 20% | 29,684,000 | 14,547,000 | 148,000 | 28,000 | 1,493,000 | 377,025,000 | 20 | 250 |
| 30% | 44,526,000 | 10,552,000 | 101,000 | 17,000 | 1,656,000 | 547,131,000 | 40 | 1,040 |
| 40% | 59,368,000 | 8,374,000 | 79,000 | 14,000 | 1,704,000 | 717,238,000 | 80 | 3,570 |
| 50% | 74,199,000 | 7,093,000 | 69,000 | 12,000 | 1,733,000 | 887,344,000 | 130 | 5,890 |
| 60% | 89,041,000 | 6,695,000 | 65,000 | 11,000 | 1,741,000 | 1,057,451,000 | 430 | 20,080 |
| 70% | 103,883,000 | 6,390,000 | 63,000 | 11,000 | 1,741,000 | 1,227,557,000 | 560 | dominated |
| **2.0x total cost of vaccination program** |  |  |  |  |  |  |  |  |
| 0% | - | 32,400,000 | 451,000 | 141,000 | - | - | - | - |
| 20% | 29,684,000 | 14,547,000 | 148,000 | 28,000 | 1,493,000 | 754,050,000 | 40 | 500 |
| 30% | 44,526,000 | 10,552,000 | 101,000 | 17,000 | 1,656,000 | 1,094,263,000 | 90 | 2,090 |
| 40% | 59,368,000 | 8,374,000 | 79,000 | 14,000 | 1,704,000 | 1,434,476,000 | 160 | 7,140 |
| 50% | 74,199,000 | 7,093,000 | 69,000 | 12,000 | 1,733,000 | 1,774,689,000 | 270 | 11,770 |
| 60% | 89,041,000 | 6,695,000 | 65,000 | 11,000 | 1,741,000 | 2,114,902,000 | 860 | 40,170 |
| 70% | 103,883,000 | 6,390,000 | 63,000 | 11,000 | 1,741,000 | 2,455,115,000 | 1,120 | dominated |

**Abbreviations:** YLS, years of life saved; US\$, US Dollar; ICER, incremental cost-effectiveness ratio.
Total population vaccinated, COVID-19 infections, hospital and ICU admissions, deaths, total YLS, and total cost of vaccination are rounded to the nearest thousand. Dollars per incremental infection prevented and ICERs are calculated using unrounded values and then rounded to the nearest ten.
\* Dominated strategies are ones that are more costly and less effective than another strategy or ones that have a higher ICER than a more effective strategy, and are thus not economically efficient. In incremental scenarios resulting in small changes in the total number of infections or deaths, strategies may appear to be dominated due to random variation.

**Extended Data Table 6.**
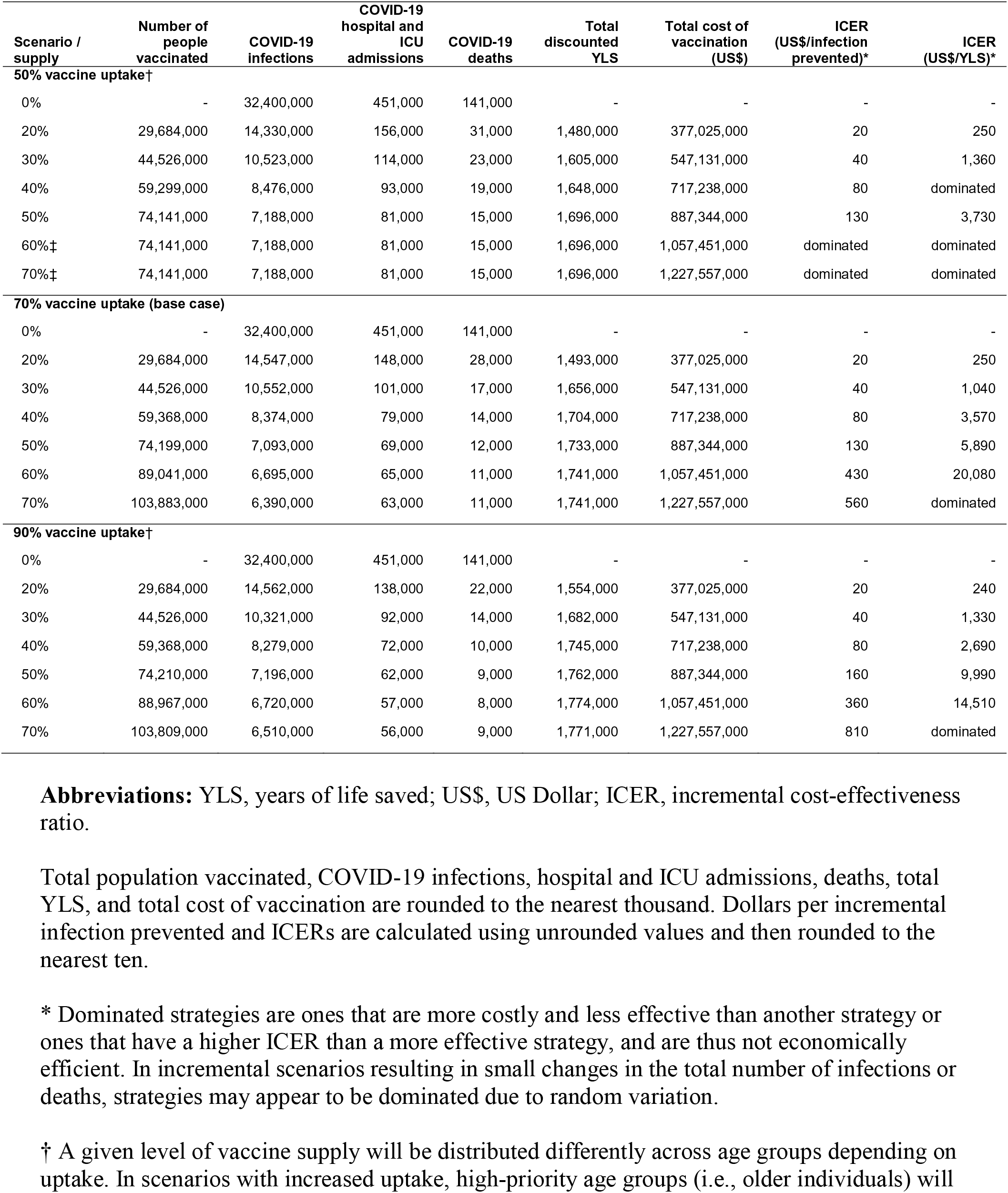
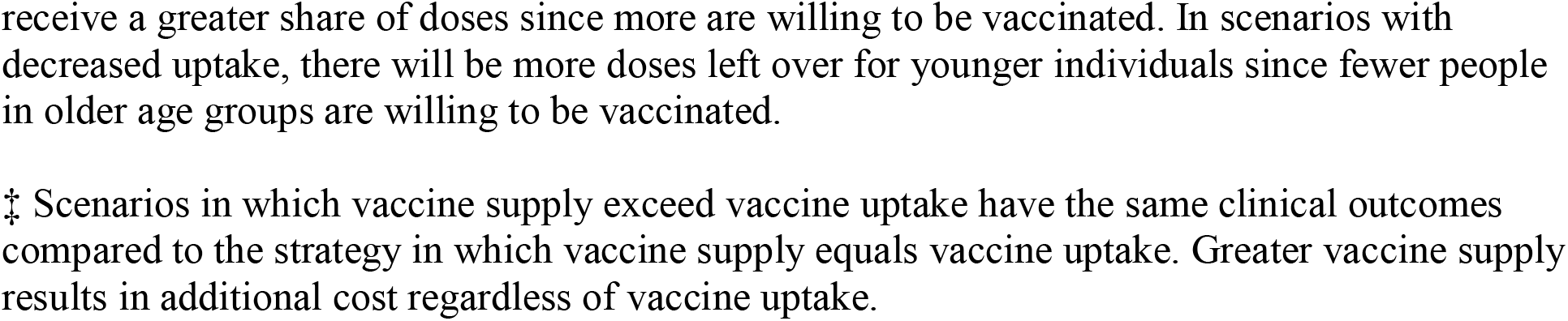
One-way sensitivity analyses: influence of vaccine uptake on clinical and economic outcomes across 9 representative countries.

| Scenario / supply | Number of people vaccinated | COVID-19 infections | COVID-19 hospital and ICU admissions | COVID-19 deaths | Total discounted YLS | Total cost of vaccination (US\$) | ICER (US\$/infection prevented)* | ICER (US\$/YLS)* |
| --- | --- | --- | --- | --- | --- | --- | --- | --- |
| **50% vaccine uptake†** |  |  |  |  |  |  |  |  |
| 0% | - | 32,400,000 | 451,000 | 141,000 | - | - | - | - |
| 20% | 29,684,000 | 14,330,000 | 156,000 | 31,000 | 1,480,000 | 377,025,000 | 20 | 250 |
| 30% | 44,526,000 | 10,523,000 | 114,000 | 23,000 | 1,605,000 | 547,131,000 | 40 | 1,360 |
| 40% | 59,299,000 | 8,476,000 | 93,000 | 19,000 | 1,648,000 | 717,238,000 | 80 | dominated |
| 50% | 74,141,000 | 7,188,000 | 81,000 | 15,000 | 1,696,000 | 887,344,000 | 130 | 3,730 |
| 60%‡ | 74,141,000 | 7,188,000 | 81,000 | 15,000 | 1,696,000 | 1,057,451,000 | dominated | dominated |
| 70%‡ | 74,141,000 | 7,188,000 | 81,000 | 15,000 | 1,696,000 | 1,227,557,000 | dominated | dominated |
| **70% vaccine uptake (base case)** |  |  |  |  |  |  |  |  |
| 0% | - | 32,400,000 | 451,000 | 141,000 | - | - | - | - |
| 20% | 29,684,000 | 14,547,000 | 148,000 | 28,000 | 1,493,000 | 377,025,000 | 20 | 250 |
| 30% | 44,526,000 | 10,552,000 | 101,000 | 17,000 | 1,656,000 | 547,131,000 | 40 | 1,040 |
| 40% | 59,368,000 | 8,374,000 | 79,000 | 14,000 | 1,704,000 | 717,238,000 | 80 | 3,570 |
| 50% | 74,199,000 | 7,093,000 | 69,000 | 12,000 | 1,733,000 | 887,344,000 | 130 | 5,890 |
| 60% | 89,041,000 | 6,695,000 | 65,000 | 11,000 | 1,741,000 | 1,057,451,000 | 430 | 20,080 |
| 70% | 103,883,000 | 6,390,000 | 63,000 | 11,000 | 1,741,000 | 1,227,557,000 | 560 | dominated |
| **90% vaccine uptake†** |  |  |  |  |  |  |  |  |
| 0% | - | 32,400,000 | 451,000 | 141,000 | - | - | - | - |
| 20% | 29,684,000 | 14,562,000 | 138,000 | 22,000 | 1,554,000 | 377,025,000 | 20 | 240 |
| 30% | 44,526,000 | 10,321,000 | 92,000 | 14,000 | 1,682,000 | 547,131,000 | 40 | 1,330 |
| 40% | 59,368,000 | 8,279,000 | 72,000 | 10,000 | 1,745,000 | 717,238,000 | 80 | 2,690 |
| 50% | 74,210,000 | 7,196,000 | 62,000 | 9,000 | 1,762,000 | 887,344,000 | 160 | 9,990 |
| 60% | 88,967,000 | 6,720,000 | 57,000 | 8,000 | 1,774,000 | 1,057,451,000 | 360 | 14,510 |
| 70% | 103,809,000 | 6,510,000 | 56,000 | 9,000 | 1,771,000 | 1,227,557,000 | 810 | dominated |

**Abbreviations:** YLS, years of life saved; US\$, US Dollar; ICER, incremental cost-effectiveness ratio.
Total population vaccinated, COVID-19 infections, hospital and ICU admissions, deaths, total YLS, and total cost of vaccination are rounded to the nearest thousand. Dollars per incremental infection prevented and ICERs are calculated using unrounded values and then rounded to the nearest ten.
\* Dominated strategies are ones that are more costly and less effective than another strategy or ones that have a higher ICER than a more effective strategy, and are thus not economically efficient. In incremental scenarios resulting in small changes in the total number of infections or deaths, strategies may appear to be dominated due to random variation.
† A given level of vaccine supply will be distributed differently across age groups depending on uptake. In scenarios with increased uptake, high-priority age groups (i.e., older individuals) will
receive a greater share of doses since more are willing to be vaccinated. In scenarios with decreased uptake, there will be more doses left over for younger individuals since fewer people in older age groups are willing to be vaccinated.
‡ Scenarios in which vaccine supply exceed vaccine uptake have the same clinical outcomes compared to the strategy in which vaccine supply equals vaccine uptake. Greater vaccine supply results in additional cost regardless of vaccine uptake.

**Extended Data Table 7.** One-way sensitivity analyses: influence of administration rate on clinical and economic outcomes across 9 representative countries.

| Scenario / supply | Number of people vaccinated | COVID-19 infections | COVID-19 hospital and ICU admissions | COVID-19 deaths | Total discounted YLS | Total cost of vaccination (US\$) | ICER (US\$/infection prevented)* | ICER (US\$/YLS)* |
| --- | --- | --- | --- | --- | --- | --- | --- | --- |
| **7% of population vaccinated per month** |  |  |  |  |  |  |  |  |
| 0% | - | 32,400,000 | 451,000 | 141,000 | - | - | - | - |
| 20% | 29,689,000 | 16,057,000 | 165,000 | 29,000 | 1,474,000 | 377,025,000 | 20 | 260 |
| 30% | 44,580,000 | 12,068,000 | 119,000 | 20,000 | 1,624,000 | 547,131,000 | 40 | 1,130 |
| 40% | 59,447,000 | 10,370,000 | 103,000 | 17,000 | 1,659,000 | 717,238,000 | 100 | 4,950 |
| 50% | 74,211,000 | 9,695,000 | 94,000 | 16,000 | 1,672,000 | 887,344,000 | 250 | 12,220 |
| 60% | 89,094,000 | 9,471,000 | 94,000 | 16,000 | 1,673,000 | 1,057,451,000 | 760 | dominated |
| 70% | 103,849,000 | 9,395,000 | 94,000 | 16,000 | 1,675,000 | 1,227,557,000 | 2,230 | 130,550 |
| **10% of population vaccinated per month (base case)** |  |  |  |  |  |  |  |  |
| 0% | - | 32,400,000 | 451,000 | 141,000 | - | - | - | - |
| 20% | 29,684,000 | 14,547,000 | 148,000 | 28,000 | 1,493,000 | 377,025,000 | 20 | 250 |
| 30% | 44,526,000 | 10,552,000 | 101,000 | 17,000 | 1,656,000 | 547,131,000 | 40 | 1,040 |
| 40% | 59,368,000 | 8,374,000 | 79,000 | 14,000 | 1,704,000 | 717,238,000 | 80 | 3,570 |
| 50% | 74,199,000 | 7,093,000 | 69,000 | 12,000 | 1,733,000 | 887,344,000 | 130 | 5,890 |
| 60% | 89,041,000 | 6,695,000 | 65,000 | 11,000 | 1,741,000 | 1,057,451,000 | 430 | 20,080 |
| 70% | 103,883,000 | 6,390,000 | 63,000 | 11,000 | 1,741,000 | 1,227,557,000 | 560 | dominated |
| **15% of population vaccinated per month** |  |  |  |  |  |  |  |  |
| 0% | - | 32,400,000 | 451,000 | 141,000 | - | - | - | - |
| 20% | 29,687,000 | 14,469,000 | 142,000 | 27,000 | 1,508,000 | 377,025,000 | 20 | 250 |
| 30% | 44,531,000 | 9,771,000 | 92,000 | 15,000 | 1,684,000 | 547,131,000 | 40 | 970 |
| 40% | 59,374,000 | 7,561,000 | 71,000 | 13,000 | 1,719,000 | 717,238,000 | 80 | dominated |
| 50% | 74,486,000 | 5,858,000 | 57,000 | 10,000 | 1,758,000 | 887,344,000 | 100 | 4,550 |
| 60% | 89,309,000 | 5,085,000 | 51,000 | 9,000 | 1,772,000 | 1,057,451,000 | 220 | 12,370 |
| 70% | 104,152,000 | 4,726,000 | 48,000 | 8,000 | 1,779,000 | 1,227,557,000 | 470 | 26,740 |

**Abbreviations:** YLS, years of life saved; US\$, US Dollar; ICER, incremental cost-effectiveness ratio.
Total population vaccinated, COVID-19 infections, hospital and ICU admissions, deaths, total YLS, and total cost of vaccination are rounded to the nearest thousand. Dollars per incremental infection prevented and ICERs are calculated using unrounded values and then rounded to the nearest ten.
\* Dominated strategies are ones that are more costly and less effective than another strategy or ones that have a higher ICER than a more effective strategy, and are thus not economically efficient. In incremental scenarios resulting in small changes in the total number of infections or deaths, strategies may appear to be dominated due to random variation.

**Extended Data Table 8.**
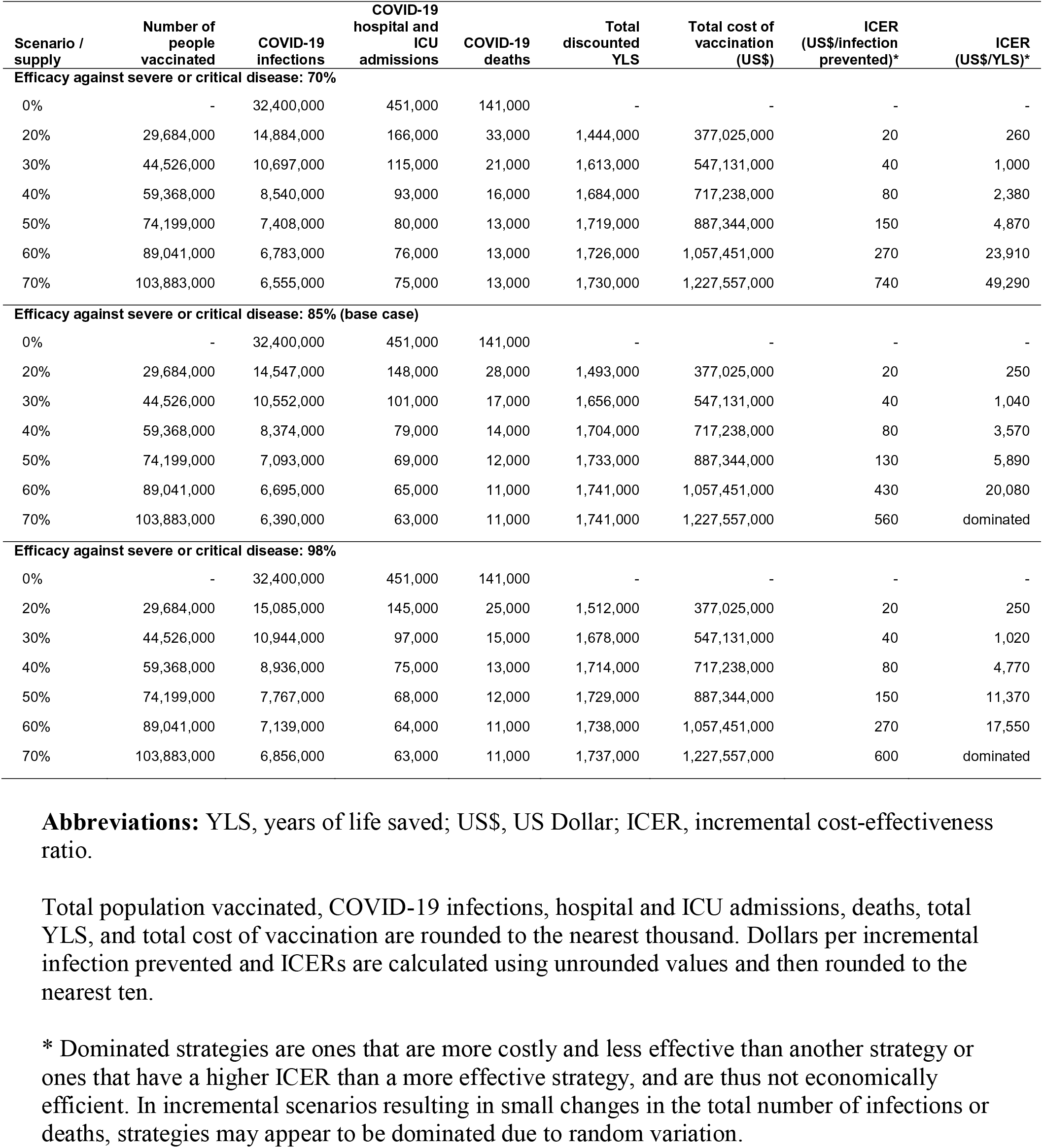
One-way sensitivity analyses: influence of vaccine efficacy against severe or critical disease on clinical and economic outcomes across 9 representative countries.

| Scenario / supply | Number of people vaccinated | COVID-19 infections | COVID-19 hospital and ICU admissions | COVID-19 deaths | Total discounted YLS | Total cost of vaccination (US\$) | ICER (US\$/infection prevented)* | ICER (US\$/YLS)* |
| --- | --- | --- | --- | --- | --- | --- | --- | --- |
| **Efficacy against severe or critical disease: 70%** |  |  |  |  |  |  |  |  |
| 0% | - | 32,400,000 | 451,000 | 141,000 | - | - | - | - |
| 20% | 29,684,000 | 14,884,000 | 166,000 | 33,000 | 1,444,000 | 377,025,000 | 20 | 260 |
| 30% | 44,526,000 | 10,697,000 | 115,000 | 21,000 | 1,613,000 | 547,131,000 | 40 | 1,000 |
| 40% | 59,368,000 | 8,540,000 | 93,000 | 16,000 | 1,684,000 | 717,238,000 | 80 | 2,380 |
| 50% | 74,199,000 | 7,408,000 | 80,000 | 13,000 | 1,719,000 | 887,344,000 | 150 | 4,870 |
| 60% | 89,041,000 | 6,783,000 | 76,000 | 13,000 | 1,726,000 | 1,057,451,000 | 270 | 23,910 |
| 70% | 103,883,000 | 6,555,000 | 75,000 | 13,000 | 1,730,000 | 1,227,557,000 | 740 | 49,290 |
| **Efficacy against severe or critical disease: 85% (base case)** |  |  |  |  |  |  |  |  |
| 0% | - | 32,400,000 | 451,000 | 141,000 | - | - | - | - |
| 20% | 29,684,000 | 14,547,000 | 148,000 | 28,000 | 1,493,000 | 377,025,000 | 20 | 250 |
| 30% | 44,526,000 | 10,552,000 | 101,000 | 17,000 | 1,656,000 | 547,131,000 | 40 | 1,040 |
| 40% | 59,368,000 | 8,374,000 | 79,000 | 14,000 | 1,704,000 | 717,238,000 | 80 | 3,570 |
| 50% | 74,199,000 | 7,093,000 | 69,000 | 12,000 | 1,733,000 | 887,344,000 | 130 | 5,890 |
| 60% | 89,041,000 | 6,695,000 | 65,000 | 11,000 | 1,741,000 | 1,057,451,000 | 430 | 20,080 |
| 70% | 103,883,000 | 6,390,000 | 63,000 | 11,000 | 1,741,000 | 1,227,557,000 | 560 | dominated |
| **Efficacy against severe or critical disease: 98%** |  |  |  |  |  |  |  |  |
| 0% | - | 32,400,000 | 451,000 | 141,000 | - | - | - | - |
| 20% | 29,684,000 | 15,085,000 | 145,000 | 25,000 | 1,512,000 | 377,025,000 | 20 | 250 |
| 30% | 44,526,000 | 10,944,000 | 97,000 | 15,000 | 1,678,000 | 547,131,000 | 40 | 1,020 |
| 40% | 59,368,000 | 8,936,000 | 75,000 | 13,000 | 1,714,000 | 717,238,000 | 80 | 4,770 |
| 50% | 74,199,000 | 7,767,000 | 68,000 | 12,000 | 1,729,000 | 887,344,000 | 150 | 11,370 |
| 60% | 89,041,000 | 7,139,000 | 64,000 | 11,000 | 1,738,000 | 1,057,451,000 | 270 | 17,550 |
| 70% | 103,883,000 | 6,856,000 | 63,000 | 11,000 | 1,737,000 | 1,227,557,000 | 600 | dominated |

**Abbreviations:** YLS, years of life saved; US\$, US Dollar; ICER, incremental cost-effectiveness ratio.
Total population vaccinated, COVID-19 infections, hospital and ICU admissions, deaths, total YLS, and total cost of vaccination are rounded to the nearest thousand. Dollars per incremental infection prevented and ICERs are calculated using unrounded values and then rounded to the nearest ten.
\* Dominated strategies are ones that are more costly and less effective than another strategy or ones that have a higher ICER than a more effective strategy, and are thus not economically efficient. In incremental scenarios resulting in small changes in the total number of infections or deaths, strategies may appear to be dominated due to random variation.

**Extended Data Table 9.** One-way sensitivity analyses: influence of vaccine efficacy against symptomatic disease on clinical and economic outcomes across 9 representative countries.

| Scenario / supply | Number of people vaccinated | COVID-19 infections | COVID-19 hospital and ICU admissions | COVID-19 deaths | Total discounted YLS | Total cost of vaccination (US\$) | ICER (US\$/infection prevented)* | ICER (US\$/YLS)* |
| --- | --- | --- | --- | --- | --- | --- | --- | --- |
| **Efficacy against symptomatic disease: 55%** |  |  |  |  |  |  |  |  |
| 0% | - | 32,400,000 | 451,000 | 141,000 | - | - | - | - |
| 20% | 29,684,000 | 15,256,000 | 154,000 | 28,000 | 1,486,000 | 377,025,000 | 20 | 250 |
| 30% | 44,526,000 | 10,576,000 | 105,000 | 18,000 | 1,641,000 | 547,131,000 | 40 | 1,100 |
| 40% | 59,368,000 | 8,453,000 | 82,000 | 14,000 | 1,699,000 | 717,238,000 | 80 | 2,960 |
| 50% | 74,199,000 | 7,301,000 | 73,000 | 12,000 | 1,732,000 | 887,344,000 | 150 | 5,090 |
| 60% | 89,041,000 | 6,881,000 | 70,000 | 12,000 | 1,733,000 | 1,057,451,000 | 400 | dominated |
| 70% | 103,883,000 | 6,530,000 | 66,000 | 11,000 | 1,741,000 | 1,227,557,000 | 480 | 40,280 |
| **Efficacy against symptomatic disease: 66% (base case)** |  |  |  |  |  |  |  |  |
| 0% | - | 32,400,000 | 451,000 | 141,000 | - | - | - | - |
| 20% | 29,684,000 | 14,547,000 | 148,000 | 28,000 | 1,493,000 | 377,025,000 | 20 | 250 |
| 30% | 44,526,000 | 10,552,000 | 101,000 | 17,000 | 1,656,000 | 547,131,000 | 40 | 1,040 |
| 40% | 59,368,000 | 8,374,000 | 79,000 | 14,000 | 1,704,000 | 717,238,000 | 80 | 3,570 |
| 50% | 74,199,000 | 7,093,000 | 69,000 | 12,000 | 1,733,000 | 887,344,000 | 130 | 5,890 |
| 60% | 89,041,000 | 6,695,000 | 65,000 | 11,000 | 1,741,000 | 1,057,451,000 | 430 | 20,080 |
| 70% | 103,883,000 | 6,390,000 | 63,000 | 11,000 | 1,741,000 | 1,227,557,000 | 560 | dominated |
| **Efficacy against symptomatic disease: 85%** |  |  |  |  |  |  |  |  |
| 0% | - | 32,400,000 | 451,000 | 141,000 | - | - | - | - |
| 20% | 29,684,000 | 14,736,000 | 148,000 | 29,000 | 1,484,000 | 377,025,000 | 20 | 250 |
| 30% | 44,526,000 | 10,467,000 | 103,000 | 17,000 | 1,654,000 | 547,131,000 | 40 | 1,000 |
| 40% | 59,368,000 | 8,400,000 | 80,000 | 13,000 | 1,706,000 | 717,238,000 | 80 | 3,280 |
| 50% | 74,199,000 | 7,163,000 | 71,000 | 13,000 | 1,722,000 | 887,344,000 | 140 | dominated |
| 60% | 89,041,000 | 6,710,000 | 68,000 | 12,000 | 1,738,000 | 1,057,451,000 | 380 | 10,580 |
| 70% | 103,883,000 | 6,461,000 | 65,000 | 12,000 | 1,737,000 | 1,227,557,000 | 680 | dominated |

**Abbreviations:** YLS, years of life saved; US\$, US Dollar; ICER, incremental cost-effectiveness ratio.
Total population vaccinated, COVID-19 infections, hospital and ICU admissions, deaths, total YLS, and total cost of vaccination are rounded to the nearest thousand. Dollars per incremental infection prevented and ICERs are calculated using unrounded values and then rounded to the nearest ten.
\* Dominated strategies are ones that are more costly and less effective than another strategy or ones that have a higher ICER than a more effective strategy, and are thus not economically efficient. In incremental scenarios resulting in small changes in the total number of infections or deaths, strategies may appear to be dominated due to random variation.

**Extended Data Table 10.** One-way sensitivity analyses: influence of vaccine efficacy against infection on clinical and economic outcomes across 9 representative countries.

| Scenario / supply | Number of people vaccinated | COVID-19 infections | COVID-19 hospital and ICU admissions | COVID-19 deaths | Total discounted YLS | Total cost of vaccination (US\$) | ICER (US\$/infection prevented)* | ICER (US\$/YLS)* |
| --- | --- | --- | --- | --- | --- | --- | --- | --- |
| **Efficacy against infection: 20%** |  |  |  |  |  |  |  |  |
| 0% | - | 32,400,000 | 451,000 | 141,000 | - | - | - | - |
| 20% | 29,684,000 | 22,846,000 | 218,000 | 47,000 | 1,214,000 | 377,025,000 | 40 | 310 |
| 30% | 44,526,000 | 19,305,000 | 167,000 | 32,000 | 1,459,000 | 547,131,000 | 50 | 690 |
| 40% | 59,368,000 | 16,833,000 | 135,000 | 24,000 | 1,566,000 | 717,238,000 | 70 | 1,590 |
| 50% | 74,199,000 | 14,779,000 | 119,000 | 22,000 | 1,613,000 | 887,344,000 | 80 | 3,620 |
| 60% | 89,041,000 | 13,871,000 | 112,000 | 21,000 | 1,621,000 | 1,057,451,000 | 190 | 19,910 |
| 70% | 103,883,000 | 13,332,000 | 108,000 | 20,000 | 1,629,000 | 1,227,557,000 | 320 | 23,690 |
| **Efficacy against infection: 40% (base case)** |  |  |  |  |  |  |  |  |
| 0% | - | 32,400,000 | 451,000 | 141,000 | - | - | - | - |
| 20% | 29,684,000 | 14,547,000 | 148,000 | 28,000 | 1,493,000 | 377,025,000 | 20 | 250 |
| 30% | 44,526,000 | 10,552,000 | 101,000 | 17,000 | 1,656,000 | 547,131,000 | 40 | 1,040 |
| 40% | 59,368,000 | 8,374,000 | 79,000 | 14,000 | 1,704,000 | 717,238,000 | 80 | 3,570 |
| 50% | 74,199,000 | 7,093,000 | 69,000 | 12,000 | 1,733,000 | 887,344,000 | 130 | 5,890 |
| 60% | 89,041,000 | 6,695,000 | 65,000 | 11,000 | 1,741,000 | 1,057,451,000 | 430 | 20,080 |
| 70% | 103,883,000 | 6,390,000 | 63,000 | 11,000 | 1,741,000 | 1,227,557,000 | 560 | dominated |
| **Efficacy against infection: 50%** |  |  |  |  |  |  |  |  |
| 0% | - | 32,400,000 | 451,000 | 141,000 | - | - | - | - |
| 20% | 29,684,000 | 11,888,000 | 125,000 | 22,000 | 1,589,000 | 377,025,000 | 20 | 240 |
| 30% | 44,526,000 | 8,297,000 | 86,000 | 14,000 | 1,704,000 | 547,131,000 | 50 | 1,490 |
| 40% | 59,368,000 | 6,409,000 | 65,000 | 11,000 | 1,747,000 | 717,238,000 | 90 | 3,900 |
| 50% | 74,199,000 | 5,620,000 | 62,000 | 11,000 | 1,752,000 | 887,344,000 | 220 | 36,090 |
| 60% | 89,041,000 | 5,352,000 | 59,000 | 10,000 | 1,755,000 | 1,057,451,000 | 630 | dominated |
| 70% | 103,883,000 | 5,229,000 | 59,000 | 10,000 | 1,759,000 | 1,227,557,000 | 1,380 | 49,970 |

**Abbreviations:** YLS, years of life saved; US\$, US Dollar; ICER, incremental cost-effectiveness ratio.
Total population vaccinated, COVID-19 infections, hospital and ICU admissions, deaths, total YLS, and total cost of vaccination are rounded to the nearest thousand. Dollars per incremental infection prevented and ICERs are calculated using unrounded values and then rounded to the nearest ten.
\* Dominated strategies are ones that are more costly and less effective than another strategy or ones that have a higher ICER than a more effective strategy, and are thus not economically efficient. In incremental scenarios resulting in small changes in the total number of infections or deaths, strategies may appear to be dominated due to random variation.

## REFERENCES

1. Sadoff, J. et al. Safety and efficacy of single-dose Ad26.COV2.S vaccine against Covid-19. N Engl J Med (2021). doi:10.1056/NEJMoa2101544.

2. Polack, F. P. et al. Safety and efficacy of the BNT162b2 mRNA Covid-19 vaccine. N Engl J Med 383, 2603–2615 (2020).

3. Voysey, M. et al. Safety and efficacy of the ChAdOx1 nCoV-19 vaccine (AZD1222) against SARS-CoV-2: an interim analysis of four randomised controlled trials in Brazil, South Africa, and the UK. Lancet 397, 99–111 (2021).

4. Baden, L. R. et al. Efficacy and safety of the mRNA-1273 SARS-CoV-2 vaccine. N Engl J Med 384, 403–416 (2021).

5. World Health Organization. Director-General’s opening remarks at the media briefing on COVID-19 – 9 April 2021. World Health Organization https://www.who.int/director-general/speeches/detail/director-general-s-opening-remarks-at-the-media-briefing-on-covid-19-9-april-2021 (2021).

6. Nkengasong, J. N., Ndembi, N., Tshangela, A. & Raji, T. COVID-19 vaccines: how to ensure Africa has access. Nature 586, 197–199 (2020).

7. Gavi. One World Protected: The Gavi COVAX AMC Investment Opportunity. https://www.gavi.org/gavi-covax-amc-launch-event-april-2021 (2021).

8. Reddy, K. P. et al. Cost-effectiveness of public health strategies for COVID-19 epidemic control in South Africa: a microsimulation modelling study. Lancet Glob Health 9, e120–e129 (2021).

9. Emanuel, E. J. et al. An ethical framework for global vaccine allocation. Science 369, 1309–1312 (2020).

10. International Chamber of Commerce. Study shows vaccine nationalism could cost rich countries US$4.5 trillion. https://iccwbo.org/media-wall/news-speeches/study-shows-vaccine-nationalism-could-cost-rich-countries-us4-5-trillion/ (2021).

11. Chinazzi, M. et al. Estimating the effect of cooperative versus uncooperative strategies of COVID-19 vaccine allocation: a modeling study. Northeastern University Network Science Institute https://www.networkscienceinstitute.org/publications/estimating-the-effect-of-cooperative-versus-uncooperative-strategies-of-covid-19-vaccine-allocation-a-modeling-study (2020).

12. Dong, E., Du, H. & Gardner, L. An interactive web-based dashboard to track COVID-19 in real time. Lancet Infect Dis 20, 533–534 (2020).

13. World Health Organization. WHO coronavirus (COVID-19) dashboard. World Health Organization https://covid19.who.int/table (2021).

14. Mwananyanda, L. et al. Covid-19 deaths in Africa: prospective systematic postmortem surveillance study. BMJ, n334 (2021) doi:10.1136/bmj.n334.

15. Mulenga, L. B. et al. Prevalence of SARS-CoV-2 in six districts in Zambia in July, 2020: a cross-sectional cluster sample survey. Lancet Glob Health (2021). doi:10.1016/S2214-109X(21)00053-X.

16. Organisation for Economic Co-operation and Development. COVID-19 and Africa: Socio-economic implications and policy responses. OECD https://www.oecd.org/coronavirus/policy-responses/covid-19-and-africa-socio-economic-implications-and-policy-responses-96e1b282/ (2020).

17. Dorward, J. et al. The impact of the COVID-19 lockdown on HIV care in 65 South African primary care clinics: an interrupted time series analysis. Lancet HIV 8, e158–e165 (2021).

18. Kim, A. W., Nyengerai, T. & Mendenhall, E. Evaluating the mental health impacts of the COVID-19 pandemic: perceived risk of COVID-19 infection and childhood trauma predict adult depressive symptoms in urban South Africa. Psychol Med (2020). doi:10.1017/S0033291720003414.

19. Weiss, D. J. et al. Indirect effects of the COVID-19 pandemic on malaria intervention coverage, morbidity, and mortality in Africa: a geospatial modelling analysis. Lancet Infect Dis 21, 59–69 (2021).

20. Roberton, T. et al. Early estimates of the indirect effects of the COVID-19 pandemic on maternal and child mortality in low-income and middle-income countries: a modelling study. Lancet Glob Health 8, e901–e908 (2020).

21. Gavi. G7 backs Gavi’s COVAX Advance Market Commitment to boost COVID-19 vaccines in world’s poorest countries. Gavi https://www.gavi.org/news/media-room/g7-backs-gavis-covax-amc-boost-covid-19-vaccines-worlds-poorest-countries (2021).

22. Fontanet, A. & Cauchemez, S. COVID-19 herd immunity: where are we? Nat Rev Immunol 20, 583–584 (2020).

23. Chang, A. Y. et al. Past, present, and future of global health financing: a review of development assistance, government, out-of-pocket, and other private spending on health for 195 countries, 1995–2050. Lancet 393, 2233–2260 (2019).

24. Menon, S. India coronavirus: Can its vaccine producers meet demand? BBC News (2021).

25. Losina, E. et al. College campuses and COVID-19 mitigation: clinical and economic value. Ann Intern Med 174, 472–483 (2021).

26. Neilan, A. M. et al. Clinical impact, costs, and cost-effectiveness of expanded SARS-CoV-2 testing in Massachusetts. Clin Infect Dis (2020). doi:10.1093/cid/ciaa1418.

27. Baggett, T. P. et al. Clinical outcomes, costs, and cost-effectiveness of strategies for adults experiencing sheltered homelessness during the COVID-19 pandemic. JAMA Netw Open 3, e2028195 (2020).

28. COVAX Working Group. Costs of delivering COVID-19 vaccine in 92 AMC countries. https://www.who.int/publications/m/item/costs-of-delivering-covid-19-vaccine-in-92-amc-countries (2021).

29. Wouters, O. J. et al. Challenges in ensuring global access to COVID-19 vaccines: production, affordability, allocation, and deployment. Lancet 397, 1023–1034 (2021).

30. Heaton, L. M. et al. Estimating the impact of the US President’s Emergency Plan for AIDS Relief on HIV treatment and prevention programmes in Africa. Sex Transm Infect 91, 615–620 (2015).

31. Kaiser Family Foundation. U.S. Global Health Budget: HIV/PEPFAR. https://files.kff.org/attachment/Fact-Sheet-US-Global-Health-Budget-HIV-PEPFAR.pdf (2020).

32. Peter G. Peterson Foundation. Here’s Everything the Federal Government Has Done to Respond to the Coronavirus So Far. Peter G. Peterson Foundation https://www.pgpf.org/blog/2021/03/heres-everything-congress-has-done-to-respond-to-the-coronavirus-so-far (2021).

33. Uyoga, S. et al. Seroprevalence of anti–SARS-CoV-2 IgG antibodies in Kenyan blood donors. Science 371, 79–82 (2021).

34. Murhekar, M. V. et al. SARS-CoV-2 antibody seroprevalence in India, August–September, 2020: findings from the second nationwide household serosurvey. Lancet Glob Health 9, e257–e266 (2021).

35. Madhi, S. A. et al. Efficacy of the ChAdOx1 nCoV-19 Covid-19 vaccine against the B.1.351 variant. N Engl J Med (2021). doi:10.1056/NEJMoa2102214.

36. Callaway, E. Fast-spreading COVID variant can elude immune responses. Nature 589, 500–501 (2021).

37. Sabino, E. C. et al. Resurgence of COVID-19 in Manaus, Brazil, despite high seroprevalence. Lancet 397, 452–455 (2021).

38. Salyer, S. J. et al. The first and second waves of the COVID-19 pandemic in Africa: a cross-sectional study. Lancet 397, 1265–1275 (2021).

39. O’Driscoll, M. et al. Age-specific mortality and immunity patterns of SARS-CoV-2. Nature 590, 140–145 (2021).

40. South African Medical Research Council. Report on Weekly Deaths in South Africa. South African Medical Research Council https://www.samrc.ac.za/reports/report-weekly-deaths-south-africa (2021).

41. Ojal, J. et al. Revealing the extent of the COVID-19 pandemic in Kenya based on serological and PCR-test data. Preprint at https://www.medrxiv.org/content/10.1101/2020.09.02.20186817v1 (2020).

42. Ghisolfi, S. et al. Predicted COVID-19 fatality rates based on age, sex, comorbidities and health system capacity. BMJ Glob Health 5, e003094 (2020).

43. Onovo, A., et al. Estimates of the COVID-19 infection fatality rate for 48 African countries: a model-based analysis. Preprint at https://papers.ssrn.com/sol3/papers.cfm?abstract_id=3657607 (2020).

44. Lawal, Y. Africa’s low COVID-19 mortality rate: A paradox? Int J Infect Dis 102, 118–122 (2021).

45. Njenga, M. K. et al. Why is there low morbidity and mortality of COVID-19 in Africa? Am J Trop Med Hyg 103, 564–569 (2020).

46. Novosad, P., Jain, R., Campion, A. & Asher, S. COVID-19 mortality effects of underlying health conditions in India: a modelling study. BMJ Open 10, e043165 (2020).

47. Docherty, A. B. et al. Features of 20 133 UK patients in hospital with covid-19 using the ISARIC WHO Clinical Characterisation Protocol: prospective observational cohort study. BMJ 369, m1985 (2020).

48. Headey, D. et al. Impacts of COVID-19 on childhood malnutrition and nutrition-related mortality. Lancet 396, 519–521 (2020).

49. Akseer, N., Kandru, G., Keats, E. C. & Bhutta, Z. A. COVID-19 pandemic and mitigation strategies: implications for maternal and child health and nutrition. Am J Clin Nutr 112, 251–256 (2020).

## METHODS REFERENCES

50. Martcheva, M. An Introduction to Mathematical Epidemiology. Springer, New York, NY (2015).

51. Chandrashekar, A. et al. SARS-CoV-2 infection protects against rechallenge in rhesus macaques. Science 369, 812–817 (2020).

52. Attema, A. E., Brouwer, W. B. F. & Claxton, K. Discounting in Economic Evaluations. Pharmacoeconomics 36, 745–758 (2018).

53. United Nations, Department of Economic and Social Affairs, Population Division. World Population Prospects 2019, Online Edition, Rev. 1. https://population.un.org/wpp/Download/Standard/Population/ (2019).

54. Lopez, A.D., Mathers, C.D., Ezzati, M., Jamison, D.T., & Murray, C.J.L. Global burden of disease and risk factors. The International Bank for Reconstruction and Development, New York, NY. Oxford University Press, Washington, DC (2006).

55. Bubar, K. M. et al. Model-informed COVID-19 vaccine prioritization strategies by age and serostatus. Science 371, 916–921 (2021).

56. ModernaTX, Inc. A Study to Evaluate Safety and Effectiveness of mRNA-1273 Vaccine in Healthy Children Between 6 Months of Age and Less Than 12 Years of Age. https://clinicaltrials.gov/ct2/show/NCT04796896 (2021).

57. BioNTech SE & Pfizer. Study to Evaluate the Safety, Tolerability, and Immunogenicity of an RNA Vaccine Candidate Against COVID-19 in Healthy Children <12 Years of Age. https://clinicaltrials.gov/ct2/show/NCT04816643 (2021).

58. Pfizer. Pfizer-BioNTech announce positive topline results of pivotal COVID-19 vaccine study in adolescents. Pfizer https://www.pfizer.com/news/press-release/press-release-detail/pfizer-biontech-announce-positive-topline-results-pivotal (2021).

59. Paulden, M. Calculating and interpreting ICERs and net benefit. Pharmacoeconomics 38, 785–807 (2020).

60. Wang, D. et al. Clinical characteristics of 138 hospitalized patients with 2019 novel coronavirus–infected pneumonia in Wuhan, China. JAMA 323, 1061 (2020).

61. Zhou, F. et al. Clinical course and risk factors for mortality of adult inpatients with COVID-19 in Wuhan, China: a retrospective cohort study. Lancet 395, 1054–1062 (2020).

62. World Health Organization. Report of the WHO-China Joint Mission on Coronavirus Disease 2019 (COVID-19). https://www.who.int/docs/default-source/coronaviruse/who-china-joint-mission-on-covid-19-final-report.pdf (2020).

63. Hu, Z. et al. Clinical characteristics of 24 asymptomatic infections with COVID-19 screened among close contacts in Nanjing, China. Sci China Life Sci 63, 706–711 (2020).

64. Liu, Y., Gayle, A. A., Wilder-Smith, A. & Rocklöv, J. The reproductive number of COVID-19 is higher compared to SARS coronavirus. J Travel Med 27, taaa021 (2020).

65. Arora, R. K. et al. SeroTracker: a global SARS-CoV-2 seroprevalence dashboard. Lancet Infect Dis 21, e75–e76 (2021).

66. Chibwana, M. G. et al. High SARS-CoV-2 seroprevalence in health care workers but relatively low numbers of deaths in urban Malawi. Preprint at https://www.medrxiv.org/content/10.1101/2020.07.30.20164970v3 (2020).

67. Centers for Disease Control and Prevention. Interim Guidance on Duration of Isolation and Precautions for Adults with COVID-19. Centers for Disease Control and Prevention https://www.cdc.gov/coronavirus/2019-ncov/hcp/duration-isolation.html (2021).

68. U.S. Food and Drug Administration. FDA Briefing Document Janssen Ad26.COV2.S Vaccine for the Prevention of COVID-19. https://www.fda.gov/media/146217/download (2021).

69. Gavi. New collaboration makes further 100 million doses of COVID-19 vaccine available to low- and middle-income countries. https://www.gavi.org/news/media-room/new-collaboration-makes-further-100-million-doses-covid-19-vaccine-available-low (2020).

70. U.S. Food and Drug Administration. FDA Briefing Document Pfizer-BioNTech COVID-19 Vaccine. https://www.fda.gov/media/144245/download (2020).

71. World Health Organization. Third round of allocation Pfizer-BioNTech vaccine. https://www.who.int/publications/m/item/third-round-of-allocation-pfizer-biontech-vaccine (2021).

72. World Health Organization. COVAX announces additional deals to access promising COVID-19 vaccine candidates; plans global rollout starting Q1 2021. https://www.who.int/news/item/18-12-2020-covax-announces-additional-deals-to-access-promising-covid-19-vaccine-candidates-plans-global-rollout-starting-q1-2021 (2020).

73. He, X. et al. Temporal dynamics in viral shedding and transmissibility of COVID-19. Nat Med 26, 672–675 (2020).

74. Lazarus, J. V. et al. A global survey of potential acceptance of a COVID-19 vaccine. Nat Med 27, 1–4 (2020).

