## Supplemental Methods, Tables, and Figures for "Cost-effectiveness of COVID-19 vaccination in low- and middle-income countries"

Mark J. Siedner, MD, MPH

Christopher Alba, BS

Kieran P. Fitzmaurice, BS

Rebecca Gilbert, BA

Justine A. Scott, MPH

Fatma M. Shebl, MD, MPH

Andrea Ciaranello, MD, MPH

Krishna P. Reddy, MD, MS

Kenneth A. Freedberg, MD, MSc

**Table of Contents:**

**Supplementary Methods** ... **3**

**Model Structure and Analytic Overview** ... 3
Overview ... 3
Health states and natural history of COVID-19 ... 3
Transmission ... 4
Model validation ... 5
**Input Parameters** ... 6
Population estimates ... 6
Hospital and ICU capacity ... 6
Disease natural history ... 6
Cost-effectiveness comparisons to PEPFAR ... 7

**Supplementary References** ... **8**

**Supplementary Tables** ... **11**

**Supplementary Table 1.** Country-specific inputs ... 11
**Supplementary Table 2.** Additional inputs associated with the natural history of COVID-19 disease. ... 13
**Supplementary Table 3.** Clinical and cost outcomes of the investment into the COVAX AMC initiative, regional and country breakdown. ... 14

**Supplementary Figures** ... **15**

**Supplementary Figure 1.** Illustration of health states and disease paths in the Clinical and Economic Analysis of COVID-19 Interventions (CEACOV) model. ... 15
**Supplementary Figure 2.** Two-way sensitivity analysis: influence of $R_e$ and vaccination cost on cost-effectiveness across 9 representative countries. ... 16

### Supplementary Methods

#### Model Structure and Analytic Overview

***Overview***

The Clinical and Economic Analysis of COVID-19 Interventions (CEACOV) model incorporates aspects of SARS-CoV-2 transmission, COVID-19 disease natural history, and public health interventions to project epidemic growth, clinical outcomes, healthcare resource utilization and costs.

Country-specific projections for each of the COVAX Advance Market Commitment (AMC) eligible economies are generated by simulating a cohort of 1 million individuals (agents) over a 360-day time horizon. Simulation outputs are scaled to reflect the total population of each country, and subsequently aggregated to produce regional and global projections of clinical and economic outcomes under different levels of vaccination program funding and coverage.

We measured the clinical benefit of expanded vaccination coverage in terms of the years of life saved (YLS) from prevented COVID-19 deaths, with life-years occurring in future periods discounted at a rate of 3% per year. We considered capital costs associated with program startup, as well as variable costs associated with the purchase and distribution of additional vaccines. The total program cost was treated as an upfront investment in the COVAX AMC program from the donor perspective, and thus undiscounted. Currency was expressed in 2020 United States dollars (US$).

We compared outcomes under several strategies of investment into the COVAX AMC program: zero program funding, corresponding to 0% vaccination coverage in AMC-eligible economies; funding targets as of April 2021[7], corresponding to 20% vaccination coverage; and outlays to expand vaccination coverage in AMC-eligible economies to between 30% and 70% in increments of 10%.

***Health states and natural history of COVID-19***

CEACOV is a dynamic microsimulation model of COVID-19 based on a stochastic susceptible-exposed-infectious-recovered (SEIR) framework. Individuals face daily state-transition probabilities that govern movement between the following health states (Supplementary Figure 1):

- *Susceptible*: individuals who are not currently infected with SARS-CoV-2 but may become infected during the model simulation horizon
- *Pre-infectious incubation*: individuals who have been exposed to SARS-CoV-2 but are not yet infectious to others
- *Asymptomatic*: individuals infected with SARS-CoV-2 who do not display symptoms but can still transmit the virus to others
- *Mild/Moderate*: individuals infected with SARS-CoV-2 who display mild or moderate symptoms of COVID-19 but do not require hospitalization
- *Severe*: individuals with severe symptoms of COVID-19 ideally managed in a hospital but not requiring admission to an intensive care unit (ICU)
- *Critical*: individuals with life-threatening signs and symptoms of COVID-19 who require ICU care to survive
- *Recuperation*: individuals recuperating from critical illness while remaining in the hospital
- *Recovered*: individuals fully recovered from SARS-CoV-2 infection, who can no longer transmit the virus to others and are immune from reinfection during the model simulation horizon
- *Dead*: individuals who die because of critical COVID-19 disease

The state transition from susceptible to pre-infectious incubation is dependent on the prevalence of active disease at a given point in time. Once infected, individuals have an age-dependent probability of progressing along one of four “paths” culminating in either asymptomatic infection, mild/moderate disease, severe disease, or critical disease (Supplementary Figure 1). Before reaching a more advanced disease state, individuals must first transition through intermediate states (e.g., individuals destined for severe disease must first pass through the asymptomatic and mild/moderate health states).

Hospital and ICU care improves the chances of survival for those at risk of developing critical COVID-19 disease in two ways: by providing a chance at early recovery to those in the severe state who have access to a hospital bed (thus bypassing the critical state) and by increasing the chances of survival for those in the critical state who have access to ICU care. The CEACOV model incorporates constraints on health care resources, and in the event hospital or ICU capacity is reached, individuals will default to the next highest level of treatment.

***Transmission***

In CEACOV, infected individuals in asymptomatic, mild/moderate, severe, critical, and recuperation states can transmit SARS-CoV-2 to susceptible individuals; individuals in the pre-infectious incubation or recovered states cannot transmit the virus.

The basic reproduction number ($R_0$) is defined as the daily rate in a fully susceptible cohort at which an infected individual contacts and infects susceptible individuals, multiplied by the duration of infectivity:

$$R_0 = K \times b \times D, \text{ where}$$

- *K*: number of contacts per day an infected individual has with susceptible people in a fully susceptible cohort
- *b*: the probability of transmission per contact between an infectious individual and a susceptible person
- *D*: the mean duration of infectivity.

The effective transmission rate of infected individuals in one of the infected states (asymptomatic, mild/moderate, severe, critical, and recuperation) is calculated as follows:

$$\text{Effective transmission rate } (R_{eff}) = \text{Nominal transmission rate } (R_{nom}) * \text{transmission multiplier,}$$
$$\text{where } R_{nom} = \frac{R_0}{D}.$$

The Nominal Transmission Rate is defined as $R_0$ in a fully susceptible cohort divided by the average duration of infectivity (D). Equivalently, it can be defined as a function of the average number of contacts per day an infected individual has with susceptible people in a fully susceptible cohort (K) multiplied by the probability of transmission per contact between an infectious individual and a susceptible person (b), therefore the contact rates and probability of transmission per contact are implicitly included in the nominal transmission rate. $R_0$ estimates the expected number of secondary cases produced by an infected individual in a fully susceptible cohort. Once the epidemic is underway and a subset of the population is infected, the effective reproductive number ($R_e$) measures the number of secondary cases produced by an infected individual in a progressed epidemic. This Nominal Transmission Rate captures the ratio (rather than the magnitude) of daily infectivity across different infection states (e.g., asymptomatic to mild/moderate, or mild/moderate to severe). $R_{eff}$ determines the effective transmission rate of infected individuals on each day of simulation and is equivalent to the effective reproductive number ($R_e$) divided by the duration of infectivity. Therefore, the effective transmission rate ($R_{eff}$) in CEACOV and the effective reproductive number ($R_e$) are directly related.

In the base case, we calibrated transmission multipliers to achieve an $R_e$ of 1.2 when 0% of the population is vaccinated, representing a modest epidemic growth rate. Estimates of country-specific $R_e$ values are tracked by Abbott *et al.* and estimates are available for 78 of the 91 modeled countries (86%)[75]. On April 12, 2021, the median $R_e$ among the 78 included countries was 1.00 (IQR: 0.24). However, these estimates are based on reported case data and are impacted by changes in testing volume over time according to Abbott *et al*. Moreover, the epidemic in the past year has taken the pattern of undulating waves characterized by $R_e$ levels that often reach levels of 2-3 at their height with valleys closer to 1.0 in the interim. Because of the uncertainty and changing nature of this parameter over time, we conducted sensitivity analyses in which we modeled epidemic growth with $R_e$ ranging from 1.1 to 1.5.

We assumed that all susceptible persons have an equal probability of contacting infected individuals and acquiring the virus (i.e., homogenous mixing). As the epidemic grows, the number of susceptible persons declines. Thus, not all daily contacts of infected individuals will be with susceptible persons. The daily infection rate for a susceptible person is equal to the sum of transmission rates from all infected persons across all infection states divided by the cohort size. This leads to an expected daily number of infections equal to the number of susceptible people multiplied by the infection rate on that day.

#### *Model validation*

We compared our modeled outcomes to those produced by the COVID-19 Scenario Analysis Tool published by the MRC Centre for Global Infectious Disease Analysis (Imperial College London)[76]. The COVID-19 Scenario Analysis Tool is an SEIR simulation model which estimates country-specific infections, deaths, and hospitalizations based on user-specified inputs, including $R_e$, hospital bed capacity, ICU bed capacity, vaccine efficacy, vaccine supply, vaccine rollout speed, and vaccine uptake. The epidemic in each country is fixed up until the day the model is run based on reported data obtained from the Center for Systems Science and Engineering at Johns Hopkins University and supplemented by data from Worldometers[12,77].

We ran the COVID-19 Scenario Analysis Tool model using our base case assumptions (Table 1) and country-specific inputs (Supplementary Table 1) to estimate the outcomes of our subset of 9 representative countries under a scenario in which vaccine supply is sufficient to vaccinate 20% of the population of each country. Base case assumptions included an $R_e$ of 1.2, vaccine efficacy against symptomatic infection of 66.1%, vaccine efficacy against critical/severe disease of 85.4%, vaccine uptake of 70%, and a rollout rate equal to vaccinating 10% of a country's population each month.

Under these assumptions, the COVID-19 Scenario Analysis Tool projected 16.4 million infections, 14,000 deaths, and 756,000 hospital and ICU admissions in the 9 representative countries over 360 days. Over the same time horizon, the CEACOV model projected 14.5 million infections, 28,000 deaths, and 148,000 hospital and ICU admissions. We project 11% fewer infections than the COVID-19 Scenario Analysis Tool. This may be attributed to the fact that the COVID-19 Scenario Analysis Tool assumes a two-dose vaccine course with protection only beginning 28 days after the second dose. Given that we modeled a single-dose Johnson & Johnson/Janssen Ad26.COV2.S vaccine, protection in our model occurs sooner and likely reduces the number of projected infections. Our estimated number of COVID-19 deaths in scenarios with 20% vaccine supply are approximately 2.0 times higher than those projected by the COVID-19 Scenario Analysis Tool in the same 20% vaccine supply scenario (0.19% versus 0.08%). An Imperial College London report estimated an overall IFR of 0.23% (95% CI: 0.14-0.42%) in low-income countries[78]. Other published IFR estimates that are adjusted for demographics, comorbidities, and health system capacity range from 0.37% in Western and Southern Sub-Saharan Africa to 1.45% for Eastern Europe[42]. Our estimated IFR of 0.19% is on the lower end of published estimates, which is expected given that we modeled a scenario in which 20% of the population would be vaccinated. In this scenario, most of those aged 60+ will have received protection from severe/critical disease and not be as likely to die from COVID-19.

Our model projects a similar number of infections as the COVID-19 Scenario Analysis Tool. While our model estimates a two-fold higher number of deaths than the COVID-19 Scenario Analysis Tool, we believe our IFR estimates are more in line with other published IFR estimates specific to low-income countries and corrected for demographics, comorbidities, and health system capacity.

### Input Parameters

#### *Population estimates*

Country-specific age structures and population estimates were based on data published by the United Nations World Population Prospects (reference year 2020) and The World Bank (reference year 2019)[53,79]. We were unable to find data on the age structures of Dominica, Kosovo, Marshall Islands, and Tuvalu; we modeled these four countries using their true total populations and age distributions from similar countries. We assumed Dominica to have the same age structure and mortality rates as the average for the United Nations Small Island Developing States Caribbean subregion. For Marshall Islands and Tuvalu, we used the age structure and mortality rates of the United Nations Pacific Island Developing Economies subregion. For Kosovo, we used the age distribution and mortality rates specific to Albania. Country-specific age structures and population estimates are presented in Supplementary Table 1.

#### *Hospital and ICU capacity*

The World Health Organization's (WHO) most recently reported estimates of country-specific hospital beds (per 10,000 population) were used to inform hospital capacities for each country [80]. Hospital beds were defined as inpatient beds available in public, private, general, and specialized hospitals and rehabilitation centers. Data for Micronesia, Dominica, Marshall Islands, Tonga, Kosovo, West Bank and Gaza, South Sudan, Vanuatu, and Tuvalu were supplemented by additional sources, such as the U.S. Central Intelligence Agency, The World Bank, WHO reports, and country-level health agency reports[81–85]. We were unable to find an estimate of overall hospital bed capacity for Papua New Guinea and instead used the median per capita hospital beds for all lower-middle income COVAX countries in this case. A full list of hospital capacity estimates, scaled to a population of 100,000, and their sources are found in Supplementary Table 1.

Country-specific ICU capacities were mostly informed by published literature on critical care capacities[86,87] and Reuters reporting[88]. Data for West Bank and Gaza, Papua New Guinea, Vanuatu, South Sudan, and Nicaragua were supplemented by additional sources, such as country-level health agency reports and news reporting [89–93]. We were unable to find estimates for Benin, Burkina Faso, Cambodia, Cameroon, Democratic People's Republic of Korea, Federated States of Micronesia, Kiribati, Kyrgyzstan, Madagascar, Marshall Islands, Mozambique, Tonga, and Uzbekistan. In these cases, we classified each country by its COVAX-defined eligibility criteria [94]–low income, lower-middle income, or additional International Development Association countries–and calculated the median number of ICU beds for each of the two income groups. The median number of ICU beds for a country's income group was used as its ICU capacity estimate if no other sources were available. ICU capacity estimates for each country, scaled to a population of 100,000, and their sources are found in Supplementary Table 1.

#### *Disease natural history*

We calculated age-stratified disease path probabilities based on data displaying the proportion of people in each age group who developed asymptomatic infections, disease requiring hospitalization, and disease requiring ICU care[62,95–98]. We derived the average duration that infected individuals spend in each health state based on the following sources: the pre-infectious times reported by the WHO-China Joint Mission on COVID-19[62] and He *et al.*[73]; time to development of pneumonia (Wang *et al.*[60]); time to ICU admission and time spent in the ICU (Zhou *et al.*[61]); and median time to death (Zhou *et al.*[61]). The time until recovery was derived from the durations of viral shedding based on polymerase chain reaction (PCR) detectability reported by the WHO-China Joint Mission on COVID-19[62], Hu *et al.*[63], and Zhou *et al.*[61]. After determining the average duration of each health state (measured in days), we calculated state transition rates as follows:

$$\text{Transition rate} = r = (\text{average duration of health state})^{-1} \tag{1}$$

Based on the transition rates estimated using equation (1), we then calculated daily state-transition probabilities as:

$$\text{Daily transition probability} = 1 - \exp(-r\Delta t) \quad (2)$$

where $\Delta t$ represents the chosen time step duration–in this case one day.

***Cost-effectiveness comparisons to PEPFAR***

We recognize that there is no established threshold by which to evaluate the cost-effectiveness of increased funding for COVID-19 vaccines in low- and middle-income countries. With this in mind, we calculated the incremental cost-effectiveness ratio of the President's Emergency Plan for AIDS Relief (PEPFAR) to provide readers with a comparison against which to ground their interpretations of cost-effectiveness. Heaton *et al.* estimated that PEPFAR-supported antiretroviral therapy, voluntary medical male circumcision, and prevention of mother-to-child transmission programs were responsible for 11,560,114 life-years saved from 2004-2013 compared to a counterfactual scenario that subtracts the direct contributions of PEPFAR to these services [30]. To determine the cost of United States government investment in these services through PEPFAR, we examined PEPFAR's annual budget for bilateral programs from 2004-2013 while adjusting for inflation[31,99]. We estimate that during this period, the cumulative cost of these programs to the United States government was US$49.8 billion (expressed in 2020 US$). This would imply that the incremental cost-effectiveness ratio of funding PEPFAR versus not funding PEPFAR was US$4,310 per year of life saved.

### Supplementary References

75. Abbott, S. *et al.* Estimating the time-varying reproduction number of SARS-CoV-2 using national and subnational case counts. *Wellcome Open Res* **5**, 112 (2020).
76. Covid-19 Scenario Analysis Tool. https://www.covidsim.org/v4.20210322/?place=us (2020).
77. Woldometer, COVID-19 Coronavirus Pandemic. https://www.worldometers.info/coronavirus/.
78. Report 34 - COVID-19 Infection Fatality Ratio Estimates from Seroprevalence. *Imperial College London* http://www.imperial.ac.uk/medicine/departments/school-public-health/infectious-disease-epidemiology/mrc-global-infectious-disease-analysis/covid-19/report-34-ifr/.
79. The World Bank. Population, total. *The World Bank* https://data.worldbank.org/indicator/SP.POP.TOTL.
80. World Health Organization. Hospital beds (per 10,000 population). *World Health Organization* https://www.who.int/data/gho/data/indicators/indicator-details/GHO/hospital-beds-(per-10-000-population) (2020).
81. Central Intelligence Agency. Hospital bed density. *The World Factbook* https://www.cia.gov/the-world-factbook/field/hospital-bed-density.
82. World Health Organization. *Kosovo Health Sector Situation Report*. (2000).
83. World Health Organization Regional Office for the Eastern Mediterranean. *Country cooperation strategy for WHO and the Occupied Palestinian Territory 2017–2020*. https://www.un.org/unispal/wp-content/uploads/2017/12/WHOREP_111217.pdf (2017).
84. United Nations Office for the Coordination of Humanitarian Affairs. United Nations South Sudan 2020 - COVID-19 Socio-Economic Response Plan. *Humanitarian Response* https://www.humanitarianresponse.info/ru/operations/south-sudan/document/covid-19-socio-economic-response-plan (2020).
85. The World Bank. Hospital beds (per 1,000 people). *The World Bank Data* https://data.worldbank.org/indicator/SH.MED.BEDS.ZS.
86. Craig, J., Kalanxhi, E. & Hauck, S. National estimates of critical care capacity in 54 African countries. Preprint at https://www.medrxiv.org/content/10.1101/2020.05.13.20100727v2 (2020).

87. Ma, X. & Vervoort, D. Critical care capacity during the COVID-19 pandemic: Global availability of intensive care beds. *J Crit Care* **58**, 96–97 (2020).

88. Houreld, K., Lewis, D., McNeill, R. & Granados, S. Virus exposes gaping holes in Africa's health systems. *Reuters Graphics* https://graphics.reuters.com/HEALTH-CORONAVIRUS/AFRICA/yzdpxoqbdvx/ (2020).

89. World Health Organization. Coronavirus disease (COVID-19) Situation Report 33, occupied Palestinian territory. https://who.createsend1.com/t/ViewEmail/j/0BE7B888005CB5172540EF23F30FEDED (2020).

90. Papua New Guinea National Department of Health & World Health Organization. *Papua New Guinea Coronavirus Disease 2019 (COVID-19) Health Situation Report #51*. https://www.health.gov.pg/covid19/PNGSR51PCOVID-19(2020-12-14).pdf (2020).

91. Vanuatu National Ministry of Health. *Coronavirus disease 2019 (COVID-19) Vanuatu Situation Report #34*. https://covid19.gov.vu/images/Situation-reports/Vanuatu_COVID19_NHEOC_SitRep_34_12032021_3_.pdf (2021).

92. Republic of South Sudan Ministry of Health. *COVID-19 Weekly Situation Report*. https://moh.gov.ss/weekly_updates.php (2020).

93. Lopez, I. Packed hospital wards cast doubt on Nicaragua's low coronavirus count, doctors say. *Reuters* (2020).

94. COVAX. *Commitment agreements*. https://www.gavi.org/sites/default/files/covid/pr/COVAX_CA_COIP_List_COVAX_PR_24-11.pdf (2020).

95. Mizumoto, K., Kagaya, K., Zarebski, A. & Chowell, G. Estimating the asymptomatic proportion of coronavirus disease 2019 (COVID-19) cases on board the Diamond Princess cruise ship, Yokohama, Japan, 2020. *Eurosurveill* **25**, 2000180 (2020).

96. Tao, Y. *et al.* High incidence of asymptomatic SARS-CoV-2 infection, Chongqing, China. Preprint at https://www.medrxiv.org/content/10.1101/2020.03.16.20037259v1 (2020).

97. Massachusetts Department of Public Health. COVID-19 Dashboard. https://www.mass.gov/doc/covid-19-dashboard-april-20-2020/download (2020).

98. Rothwell, J. Estimating COVID-19 Prevalence in Symptomatic Americans. *Gallup Blog* (2020).

99. Organization for Economic Co-operation and Development. Consumer Price Index: Total All Items for the United States. FRED, Federal Reserve Bank of St. Louis https://fred.stlouisfed.org/series/CPALTT01USA661S (2020).

100. Sen-Crowe, B., Sutherland, M., McKenney, M. & Elkbuli, A. A Closer Look Into Global Hospital Beds Capacity and Resource Shortages During the COVID-19 Pandemic. *J Surg Res* **260**, 56–63 (2021).

### Supplementary Tables

#### **Supplementary Table 1.** Country-specific inputs

| Country | Region | Population (thousands) | Age distribution (% of population) | | | Hospital bed capacity/100,000 people | | ICU bed capacity/100,000 people | |
|---|---|---|---|---|---|---|---|---|---|
| | | | 0-19y | 20-59y | ≥60y | *Estimate* | *Source* | *Estimate* | *Source* |
| Afghanistan | EMRO | 38,927 | 53.7 | 42.1 | 4.2 | 39.0 | 80 | 0.2 | 87 |
| Algeria | AFRO | 43,852 | 37.4 | 52.7 | 9.9 | 190.0 | 80 | 1.0 | 87 |
| Angola | AFRO | 32,866 | 57.1 | 39.2 | 3.7 | 80.0 | 80 | 0.4 | 86 |
| Bangladesh | SEARO | 164,688 | 36.2 | 55.8 | 8.0 | 79.5 | 80 | 0.7 | 87 |
| Benin | AFRO | 12,123 | 52.6 | 42.3 | 5.1 | 50.0 | 80 | 0.3 | 87* |
| Bhutan | SEARO | 771 | 34.0 | 57.1 | 8.9 | 174.0 | 80 | 2.2 | 87 |
| Bolivia | PAHO | 11,673 | 39.9 | 49.7 | 10.4 | 129.3 | 80 | 1.3 | 87 |
| Burkina Faso | AFRO | 20,904 | 55.4 | 40.7 | 3.9 | 40.0 | 80 | 0.3 | 87* |
| Burundi | AFRO | 11,890 | 55.5 | 40.4 | 4.1 | 79.0 | 80 | 0.1 | 86 |
| Cabo Verde | AFRO | 557 | 36.8 | 55.7 | 7.5 | 210.0 | 80 | 4.8 | 86 |
| Cambodia | WPRO | 16,719 | 39.7 | 52.8 | 7.6 | 90.0 | 80 | 1.2 | 100* |
| Cameroon | AFRO | 26,545 | 52.8 | 42.9 | 4.3 | 130.0 | 80 | 1.2 | 87* |
| Central African Republic | AFRO | 4,829 | 55.9 | 39.6 | 4.5 | 100.0 | 80 | 0.2 | 86 |
| Chad | AFRO | 16,426 | 57.8 | 38.3 | 3.9 | 40.0 | 80 | 0.4 | 86 |
| Comoros | AFRO | 872 | 49.3 | 45.5 | 5.2 | 216.0 | 80 | 1.2 | 87 |
| Congo | AFRO | 5,517 | 51.7 | 43.8 | 4.5 | 160.0 | 80 | 0.4 | 86,87 |
| Côte d'Ivoire | AFRO | 26,378 | 52.5 | 42.8 | 4.7 | 40.0 | 80 | 0.6 | 86 |
| Democratic People’s Republic of Korea | SEARO | 25,779 | 27.1 | 57.8 | 15.1 | 1,430.0 | 80 | 0.3 | Assum. |
| Democratic Republic of the Congo | AFRO | 89,562 | 56.4 | 38.9 | 4.7 | 80.0 | 80 | 0.1 | 86 |
| Djibouti | EMRO | 987 | 38.2 | 54.5 | 7.3 | 140.0 | 80 | 0.0 | 86,88 |
| Dominica | PAHO | 72 | 31.9 | 53.2 | 14.9 | 380.0 | 81 | 5.4 | 87 |
| Egypt | EMRO | 102,334 | 42.4 | 49.4 | 8.2 | 143.0 | 80 | 10.6 | 86,87 |
| El Salvador | PAHO | 6,488 | 35.7 | 52.3 | 12.1 | 119.9 | 80 | 0.7 | 87 |
| Eritrea | AFRO | 3,546 | 51.7 | 41.9 | 6.4 | 70.0 | 80 | 0.2 | 86,87 |
| Eswatini | AFRO | 1,162 | 48.5 | 45.7 | 5.8 | 210.0 | 80 | 0.5 | 86 |
| Ethiopia | AFRO | 114,962 | 51.2 | 43.5 | 5.3 | 33.0 | 80 | 0.5 | 86 |
| Federated States of Micronesia | WPRO | 115 | 40.9 | 51.3 | 7.8 | 320.0 | 81 | 1.2 | Assum. |
| Fiji | WPRO | 894 | 37.5 | 53.0 | 9.5 | 199.8 | 80 | 5.0 | 87 |
| Gambia | AFRO | 2,417 | 54.6 | 41.5 | 3.9 | 110.0 | 80 | 0.4 | 86,87 |
| Ghana | AFRO | 31,070 | 47.2 | 47.5 | 5.3 | 90.0 | 80 | 1.8 | 86 |
| Grenada | PAHO | 114 | 30.7 | 54.4 | 14.9 | 357.1 | 80 | 1.9 | 87 |
| Guinea | AFRO | 13,132 | 54.6 | 40.7 | 4.7 | 30.0 | 80 | 0.3 | 86 |
| Guinea-Bissau | AFRO | 1,968 | 52.4 | 43.0 | 4.6 | 100.0 | 80 | 2.7 | 86 |
| Guyana | PAHO | 786 | 37.3 | 51.9 | 10.8 | 171.5 | 80 | 1.5 | 87 |
| Haiti | PAHO | 11,402 | 42.5 | 49.7 | 7.7 | 70.7 | 80 | 1.1 | 87 |
| Honduras | PAHO | 9,906 | 41.1 | 51.5 | 7.4 | 64.3 | 80 | 1.5 | 87 |
| Indonesia | SEARO | 273,523 | 34.5 | 55.5 | 10.1 | 104.0 | 80 | 2.7 | 87 |
| Kenya | AFRO | 53,772 | 49.8 | 46.1 | 4.2 | 140.0 | 80 | 1.0 | 86,87 |
| Kiribati | WPRO | 118 | 44.1 | 50.0 | 5.9 | 186.0 | 80 | 1.2 | 87* |
| Kosovo | EURO | 1,794 | 24.2 | 54.6 | 21.2 | 264.0 | 82 | 5.6 | 87 |
| Kyrgyzstan | EURO | 6,525 | 40.3 | 51.6 | 8.1 | 441.3 | 80 | 1.2 | Assum. |
| Lao People's Democratic Republic | WPRO | 7,278 | 41.7 | 51.5 | 6.8 | 150.0 | 80 | 2.1 | 87 |
| Lesotho | AFRO | 2,144 | 42.4 | 50.1 | 7.6 | 130.0 | 80 | 0.5 | 86,87 |
| Liberia | AFRO | 5,057 | 51.3 | 43.5 | 5.2 | 80.0 | 80 | 0.7 | 87 |
| Madagascar | AFRO | 27,690 | 50.9 | 44.0 | 5.0 | 20.0 | 80 | 0.3 | 87* |
| Malawi | AFRO | 19,131 | 54.4 | 41.5 | 4.1 | 130.0 | 80 | 0.1 | 86,87 |
| Maldives | SEARO | 541 | 24.4 | 69.9 | 5.7 | 430.0 | 80 | 3.2 | 87 |
| Mali | AFRO | 20,251 | 58.1 | 38.0 | 3.9 | 10.0 | 80 | 0.3 | 87 |
| Marshall Islands | WPRO | 59 | 44.1 | 48.9 | 6.9 | 270.0 | 81 | 4.1 | 87* |
| Mauritania | AFRO | 4,650 | 49.8 | 45.1 | 5.1 | 40.0 | 80 | 0.2 | 86 |
| Moldova | EURO | 4,034 | 20.9 | 60.2 | 18.9 | 566.1 | 80 | 14.1 | 87 |
| Mongolia | WPRO | 3,279 | 37.8 | 54.9 | 7.3 | 800.0 | 80 | 8.8 | 87 |
| Morocco | EMRO | 36,911 | 34.8 | 53.3 | 11.9 | 100.0 | 80 | 4.6 | 86,87 |
| Mozambique | AFRO | 31,258 | 55.4 | 40.2 | 4.4 | 70.0 | 80 | 0.3 | 87* |
| Myanmar | SEARO | 54,410 | 34.8 | 55.2 | 10.0 | 104.4 | 80 | 1.1 | 87 |

**Supplementary Table 1.** Country-specific inputs

| Country | Region | Population (thousands) | Age distribution (% of population) | | | Hospital bed capacity/100,000 people | | ICU bed capacity/100,000 people | |
|---|---|---|---|---|---|---|---|---|---|
| | | | 0-19y | 20-59y | ≥60y | *Estimate* | *Source* | *Estimate* | *Source* |
| Nepal | SEARO | 29,137 | 39.8 | 51.6 | 8.7 | 30.0 | 80 | 2.8 | 87 |
| Nicaragua | PAHO | 6,622 | 38.6 | 52.7 | 8.7 | 93.0 | 80 | 8.6 | 93 |
| Niger | AFRO | 24,207 | 60.6 | 35.3 | 4.1 | 39.0 | 80 | 0.3 | 86 |
| Nigeria | AFRO | 206,142 | 54.1 | 41.4 | 4.5 | 50.0 | 80 | 0.2 | 86,87 |
| Pakistan | EMRO | 220,893 | 44.8 | 48.5 | 6.7 | 63.0 | 80 | 1.5 | 87 |
| Papua New Guinea | WPRO | 8,947 | 45.5 | 48.5 | 6.0 | 140.0 | Assum. | 1.0 | 90 |
| Philippines | WPRO | 109,581 | 39.6 | 51.8 | 8.6 | 99.0 | 80 | 2.2 | 87 |
| Rwanda | AFRO | 12,952 | 49.9 | 45.0 | 5.1 | 160.0 | 80 | 0.3 | 86,87 |
| St. Lucia | PAHO | 182 | 25.3 | 60.4 | 14.3 | 129.7 | 80 | 5.6 | 87 |
| St. Vincent and the Grenadines | PAHO | 111 | 29.7 | 55.9 | 14.4 | 432.3 | 80 | 1.8 | 87 |
| Samoa | WPRO | 200 | 46.5 | 45.5 | 8.0 | 100.0 | 80 | 2.0 | 87 |
| São Tomé and Principe | AFRO | 220 | 53.2 | 41.8 | 5.0 | 290.0 | 80 | 1.8 | 86 |
| Senegal | AFRO | 16,744 | 53.1 | 42.1 | 4.8 | 30.0 | 80 | 0.2 | 86,87 |
| Sierra Leone | AFRO | 7,975 | 51.3 | 44.1 | 4.6 | 40.0 | 80 | 0.4 | 86 |
| Solomon Islands | WPRO | 687 | 50.2 | 44.3 | 5.5 | 140.0 | 80 | 0.0 | 87 |
| Somalia | EMRO | 15,895 | 57.6 | 37.9 | 4.6 | 87.0 | 80 | 0.1 | 86,87 |
| South Sudan | AFRO | 11,191 | 52.1 | 42.8 | 5.1 | 65.0 | 84 | 0.9 | 92 |
| Sri Lanka | SEARO | 21,414 | 31.5 | 52.1 | 16.4 | 415.0 | 80 | 2.3 | 87 |
| Sudan | EMRO | 43,849 | 50.7 | 43.6 | 5.7 | 74.0 | 80 | 0.2 | 86,87 |
| Syrian Arab Republic | EMRO | 17,500 | 39.8 | 52.7 | 7.5 | 140.0 | 80 | 1.8 | 87 |
| Tajikistan | EURO | 9,538 | 45.8 | 48.5 | 5.8 | 466.7 | 80 | 6.7 | 87 |
| Tanzania | AFRO | 59,736 | 54.3 | 41.4 | 4.2 | 70.0 | 80 | 0.1 | 86 |
| Timor-Leste | SEARO | 1,318 | 48.5 | 45.0 | 6.5 | 590.0 | 80 | 0.8 | 87 |
| Togo | AFRO | 8,280 | 51.4 | 43.9 | 4.7 | 70.0 | 80 | 0.5 | 86 |
| Tonga | WPRO | 104 | 45.2 | 46.2 | 8.7 | 260.0 | 81 | 4.1 | 87* |
| Tunisia | EMRO | 11,819 | 31.0 | 55.6 | 13.4 | 218.0 | 80 | 4.3 | 86,87 |
| Tuvalu | WPRO | 12 | 44.1 | 48.9 | 6.9 | 560.0 | 85 | 8.9 | 87 |
| Uganda | AFRO | 45,741 | 57.5 | 39.3 | 3.2 | 50.0 | 80 | 0.1 | 86 |
| Ukraine | EURO | 43,733 | 20.5 | 55.8 | 23.6 | 746.3 | 80 | 12.0 | 87 |
| Uzbekistan | EURO | 33,468 | 36.4 | 55.3 | 8.3 | 397.8 | 80 | 1.2 | Assum. |
| Vanuatu | WPRO | 308 | 48.1 | 46.1 | 5.8 | 170.0 | 85 | 1.6 | 91 |
| Vietnam | WPRO | 97,339 | 29.9 | 57.8 | 12.3 | 318.0 | 80 | 0.4 | 87 |
| West Bank and Gaza | EMRO | 5,101 | 48.5 | 46.6 | 4.9 | 126.0 | 83 | 6.9 | 89 |
| Yemen | EMRO | 29,825 | 49.6 | 45.8 | 4.6 | 71.0 | 80 | 3.4 | 87 |
| Zambia | AFRO | 18,385 | 55.6 | 41.0 | 3.4 | 200.0 | 80 | 0.6 | 86,87 |
| Zimbabwe | AFRO | 14,863 | 52.9 | 42.4 | 4.6 | 170.0 | 80 | 0.2 | 86,87 |

**Abbreviations:** ICU, intensive care unit; yrs, years; AFRO, African Region; AMRO, Region of the Americas; EMRO, Eastern Mediterranean Region; EURO, European Region; SEARO, South-East Asia Region; WPRO, Western Pacific Region; Assum., assumption.

* The cited source provides a range of values (e.g., <1 bed per 100,000) as the estimate for the country's ICU capacity. To estimate a single value, we used the median ICU capacity of the country's COVAX Advance Market Commitment eligibility category as described in the Supplementary Methods.

**Supplementary Table 2.** Additional inputs associated with the natural history of COVID-19 disease.

| Parameter | Base case values | | | | Sources |
|---|---|---|---|---|---|
| Natural history of COVID-19 disease | | | | | |
| Disease path probability, stratified by age, % | Asymptomatic | Mild/Moderate | Severe | Critical | 62,95–98 |
| 0-19y | 29.93 | 69.78 | 0.25 | 0.03 | |
| 20-59y | 17.90 | 80.38 | 0.80 | 0.93 | |
| ≥60y | 17.10 | 76.37 | 1.36 | 5.16 | |
| Daily disease progression probability, stratified by path, % | Asymptomatic | Mild/Moderate | Severe | Critical | 60–63,73 |
| From pre-infectious incubation to asymptomatic | 31.93 | 31.93 | 31.93 | 31.93 | |
| From asymptomatic to mild or moderate disease | -- | 39.35 | 39.35 | 39.35 | |
| From mild or moderate disease to severe disease | -- | -- | 14.26 | 28.35 | |
| From severe disease to critical disease | -- | -- | -- | 20.17 | |
| From critical disease to recuperation | -- | -- | -- | 9.61* | |
| Daily recovery probability, stratified by path, % | Asymptomatic | Mild/Moderate | Severe | Critical | 60–63,73 |
| From asymptomatic | 9.99 | -- | -- | -- | |
| From mild or moderate disease | -- | 9.52 | -- | -- | |
| From severe disease | -- | -- | 9.08 | 5.48† | |
| From recuperation after critical disease | -- | -- | -- | 16.09 | |
| Daily mortality probability while critically ill, stratified by age, % | 0-19y | 20-59y | | ≥60y | |
| Without ICU care | 11.75 | 16.62 | | 20.33 | Assumption |
| With ICU care | 0.0006 | 0.38 | | 5.00 | 60–63,73 |

**Abbreviations**: y, years.

*The transition from the critical state to recuperation requires admission to an intensive care unit (ICU).

†Early recovery from the severe state on the critical path requires admission to a hospital.

**Supplementary Table 3.** Clinical and cost outcomes of the investment into the COVAX AMC initiative, regional and country breakdown.

*Provided as a separate, downloadable Excel file.*

**Supplementary Figures**

**Supplementary Figure 1.** Illustration of health states and disease paths in the Clinical and Economic Analysis of COVID-19 Interventions (CEACOV) model.

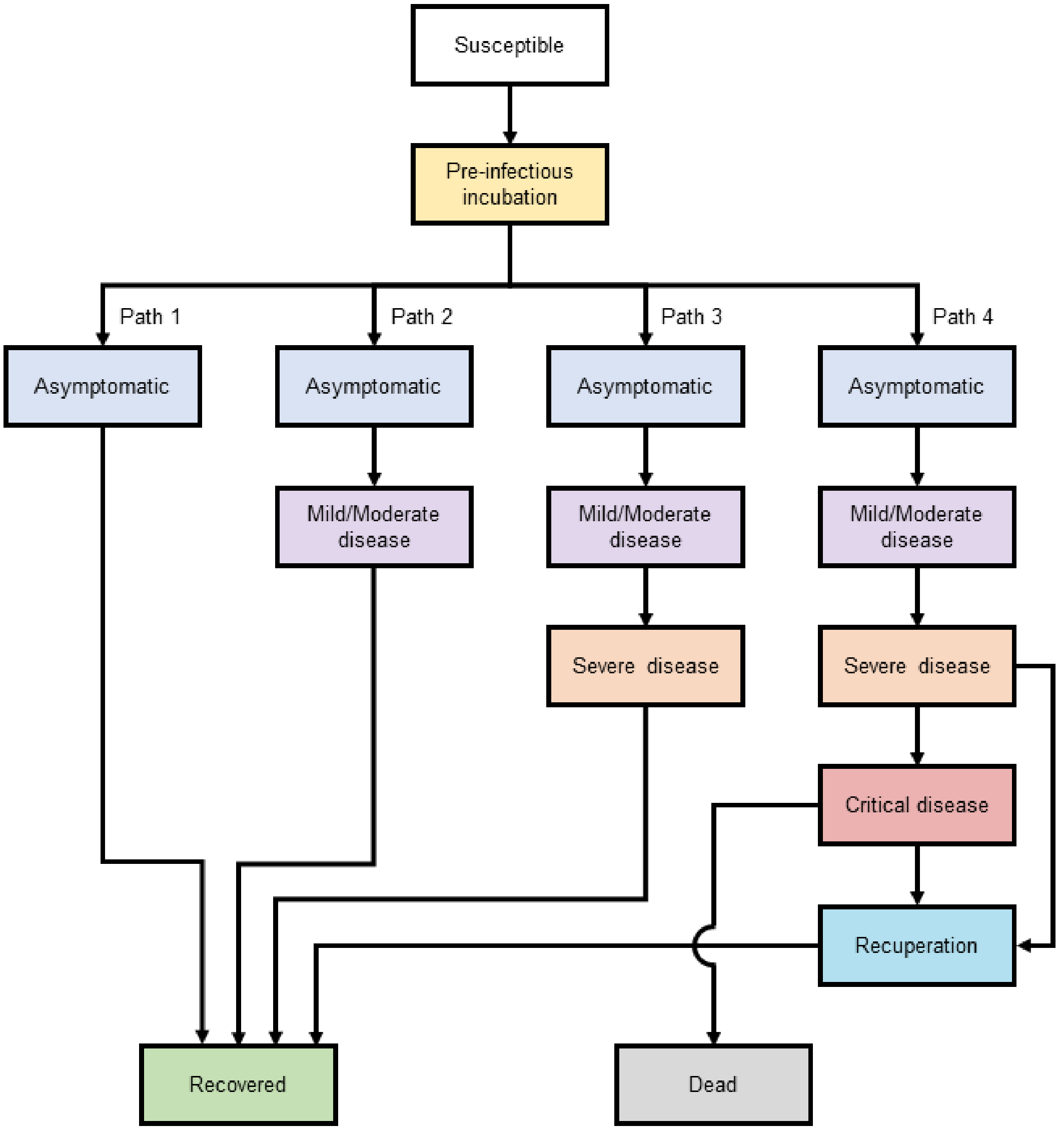

Once infected with SARS-CoV-2, individuals have an age-dependent probability of progressing along four disease paths culminating in either asymptomatic, mild/moderate, severe, or critical disease. Individuals with asymptomatic disease do not display symptoms but may still transmit the virus to others. Those with mild/moderate illness experience moderate symptoms of COVID-19 but do not require medical intervention. Those with severe disease display symptoms that will prompt hospitalization if beds are available. Those with critical disease experience life-threatening symptoms of COVID-19 and require an intensive-care bed to survive. Those recuperating from critical disease will remain hospitalized until fully recovered. Recovered individuals cannot transmit the virus to others and are immune from reinfection.

**Supplementary Figure 2.** Two-way sensitivity analysis: influence of $R_e$ and vaccination cost on cost-effectiveness across 9 representative countries.

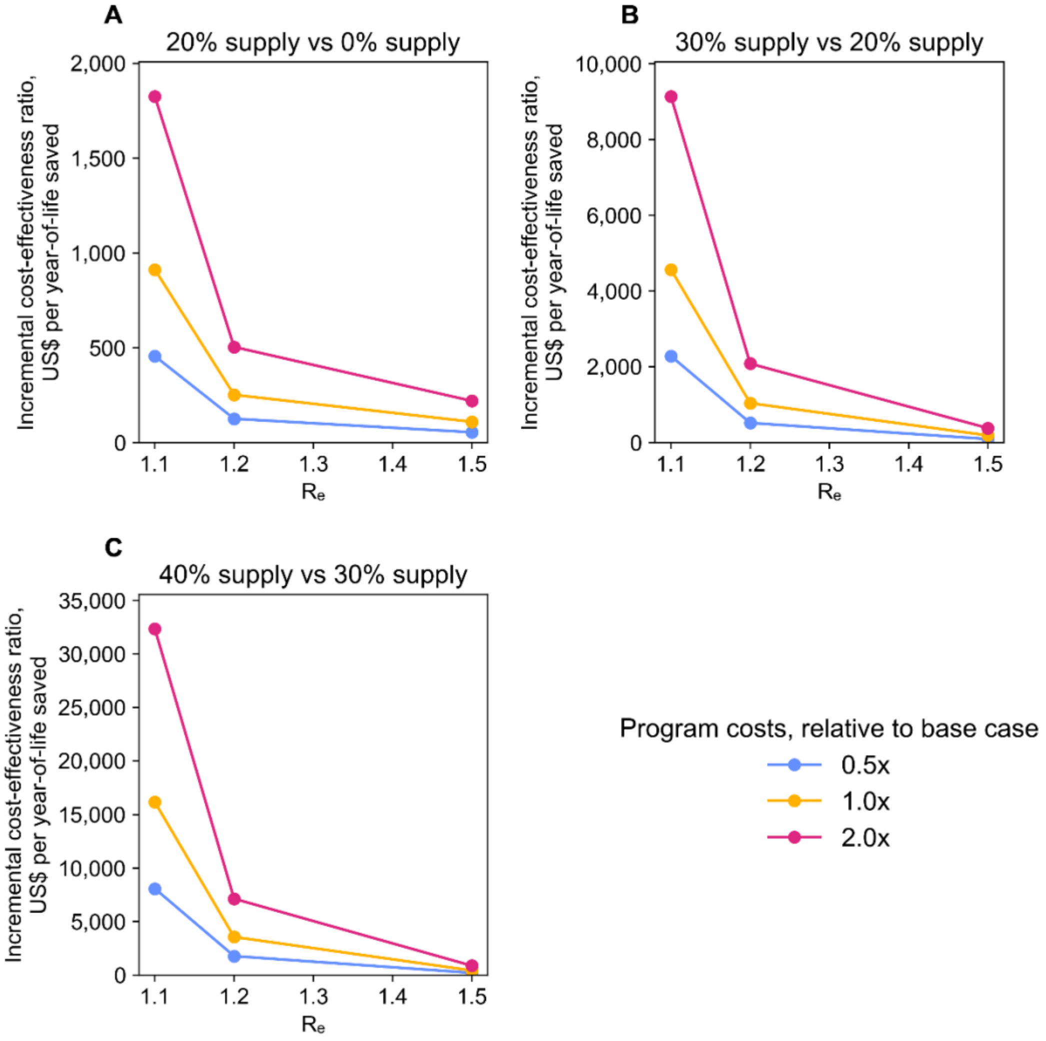

**Abbreviations***:* $R_e$, effective reproductive number.

Two-way sensitivity analyses were conducted in a subset of 9 representative countries: Afghanistan, Cambodia, Lesotho, Moldova, Mongolia, Morocco, Nicaragua, Sri Lanka, and Zambia. These countries reflect variation in global region, age structure, hospital bed capacity, and ICU bed capacity that exists among the 91 COVAX Advance Market Commitment countries (see Methods). In two-way sensitivity analyses, $R_e$ and the overall cost of the vaccination program were varied concurrently. In the base case scenario, the effective reproduction number at simulation onset was 1.2. For each scenario, $R_e$ is displayed on the horizontal axis, the incremental cost-effectiveness ratio (ICER) compared to the next-highest supply level is displayed on the vertical axis, and the program cost relative to the base case is denoted by color. The effect of concurrently varying $R_e$ and program cost on the ICER compared to the next-highest supply level is displayed for 20% supply (A), 30% supply (B), and 40% supply (C). The scales of the vertical axes differ in the three plots.

**Cost-effectiveness of COVID-19 vaccination in low- and middle-income countries**
**Mark J. Siedner et al.**

**Supplementary Table 3. Clinical and cost outcomes of the investment into the COVAX AMC initiative, regional and country breakdown**

**Abbreviations:**
- ICU = intensive care unit
- YLS = year of life saved
- ICER = incremental cost-effectiveness ratio
- AFRO = African Region
- AMRO = Region of the Americas
- EMRO = Eastern Mediterranean Region
- EURO = European Region
- SEARO = South East Asia Region
- WPRO = Western Pacific Region

**Notes:**
**Dollars per incremental infection prevented and ICERs are calculated using unrounded values and then rounded to the nearest ten.**

*** Dominated strategies are ones that are more costly and less effective than another strategy or ones that have a higher ICER than a more**

| Region | Country | Modeled vaccine supply | Total number of people vaccinated | COVID-19 infections | COVID-19 hospital and ICU admissions | COVID-19 deaths | Discounted total YLS | Total cost of vaccination (US$) | ICER (US$/infection prevented)* | ICER (US$/YLS)* |
|---|---|---|---|---|---|---|---|---|---|---|
| AFRO | All countries | 0% | - | 223,359,359 | 2,409,060 | 928,854 | - | - | - | - |
| AFRO | All countries | 20% | 210,172,381 | 102,178,961 | 805,822 | 214,051 | 9,999,208 | 2,669,456,180 | 20 | 270 |
| AFRO | All countries | 30% | 315,203,820 | 74,390,612 | 538,747 | 113,082 | 11,644,334 | 3,873,864,362 | 40 | 730 |
| AFRO | All countries | 40% | 420,493,390 | 58,502,233 | 423,358 | 80,537 | 12,128,047 | 5,078,272,544 | 80 | 2,490 |
| AFRO | All countries | 50% | 525,725,738 | 50,483,595 | 374,623 | 71,637 | 12,264,686 | 6,282,680,726 | 150 | 8,810 |
| AFRO | All countries | 60% | 630,811,930 | 46,431,410 | 357,726 | 68,864 | 12,314,293 | 7,487,088,908 | 300 | 24,280 |
| AFRO | All countries | 70% | 735,898,120 | 45,145,804 | 352,529 | 67,855 | 12,318,583 | 8,691,497,090 | 940 | 280,730 |
| AFRO | Algeria | 0% | - | 9,470,015 | 144,317 | 57,928 | - | - | - | - |
| AFRO | Algeria | 20% | 8,769,523 | 4,421,553 | 48,983 | 9,691 | 698,274 | 111,384,080 | 20 | 160 |
| AFRO | Algeria | 30% | 13,154,284 | 3,333,103 | 35,301 | 5,482 | 772,045 | 161,638,472 | 50 | 680 |
| AFRO | Algeria | 40% | 17,539,046 | 2,683,304 | 25,829 | 3,552 | 798,061 | 211,892,864 | 80 | 1,930 |
| AFRO | Algeria | 50% | 22,069,966 | 2,223,866 | 23,154 | 3,026 | 808,069 | 262,147,256 | 110 | 5,020 |
| AFRO | Algeria | 60% | 26,454,728 | 2,029,646 | 21,926 | 3,377 | 801,753 | 312,401,648 | 260 | dominated |
| AFRO | Algeria | 70% | 30,839,489 | 1,974,085 | 21,049 | 3,245 | 801,375 | 362,656,040 | 900 | dominated |
| AFRO | Angola | 0% | - | 6,564,885 | 61,722 | 22,250 | - | - | - | - |
| AFRO | Angola | 20% | 6,572,543 | 2,989,984 | 19,917 | 4,273 | 255,145 | 83,479,640 | 20 | 330 |
| AFRO | Angola | 30% | 9,858,814 | 2,032,499 | 13,081 | 2,333 | 283,603 | 121,144,076 | 40 | 1,320 |
| AFRO | Angola | 40% | 13,145,085 | 1,713,568 | 12,029 | 1,972 | 288,615 | 158,808,512 | dominated | dominated |
| AFRO | Angola | 50% | 16,431,357 | 1,350,957 | 9,597 | 1,413 | 295,069 | 196,472,948 | 110 | 6,570 |
| AFRO | Angola | 60% | 19,717,628 | 1,333,965 | 9,827 | 1,578 | 294,861 | 234,137,384 | dominated | dominated |
| AFRO | Angola | 70% | 23,003,899 | 1,247,199 | 9,597 | 1,709 | 291,227 | 271,801,820 | 730 | dominated |
| AFRO | Benin | 0% | - | 2,569,409 | 29,144 | 11,808 | - | - | - | - |
| AFRO | Benin | 20% | 2,424,358 | 1,250,572 | 9,808 | 2,679 | 125,751 | 30,792,420 | 20 | 240 |
| AFRO | Benin | 30% | 3,636,536 | 812,386 | 5,358 | 1,115 | 153,392 | 44,685,378 | 30 | 500 |
| AFRO | Benin | 40% | 4,848,715 | 690,708 | 4,995 | 1,006 | 155,239 | 58,578,336 | dominated | dominated |

| | | | | | | | | | | |
|---|---|---|---|---|---|---|---|---|---|---|
| AFRO | Benin | 50% | 6,060,894 | 563,550 | 3,916 | 740 | 158,686 | 72,471,294 | 110 | 5,250 |
| AFRO | Benin | 60% | 7,273,073 | 538,589 | 3,843 | 776 | 158,274 | 86,364,252 | 560 | dominated |
| AFRO | Benin | 70% | 8,485,251 | 522,235 | 3,673 | 667 | 160,056 | 100,257,210 | 850 | 20,290 |
| AFRO | Burkina Faso | 0% | - | 4,202,770 | 42,372 | 15,657 | - | - | - | - |
| AFRO | Burkina Faso | 20% | 4,180,382 | 1,904,563 | 13,650 | 3,366 | 176,623 | 53,096,160 | 20 | 300 |
| AFRO | Burkina Faso | 30% | 6,270,573 | 1,288,794 | 8,257 | 1,693 | 201,868 | 77,052,144 | 40 | 950 |
| AFRO | Burkina Faso | 40% | 8,291,091 | 1,068,884 | 7,233 | 1,526 | 204,500 | 101,008,128 | 110 | dominated |
| AFRO | Burkina Faso | 50% | 10,381,282 | 920,236 | 6,564 | 1,087 | 211,737 | 124,964,112 | 160 | 4,850 |
| AFRO | Burkina Faso | 60% | 12,471,473 | 838,689 | 6,104 | 1,129 | 210,728 | 148,920,096 | 290 | dominated |
| AFRO | Burkina Faso | 70% | 14,561,664 | 803,926 | 5,540 | 1,066 | 212,556 | 172,876,080 | 690 | 58,540 |
| AFRO | Burundi | 0% | - | 2,630,627 | 26,194 | 11,093 | - | - | - | - |
| AFRO | Burundi | 20% | 2,377,762 | 1,169,810 | 8,799 | 3,127 | 116,858 | 30,200,600 | 20 | 260 |
| AFRO | Burundi | 30% | 3,566,643 | 746,573 | 4,839 | 1,474 | 144,319 | 43,826,540 | 30 | 500 |
| AFRO | Burundi | 40% | 4,795,154 | 612,799 | 4,233 | 1,367 | 145,406 | 57,452,480 | 100 | dominated |
| AFRO | Burundi | 50% | 5,984,035 | 532,862 | 3,745 | 1,201 | 147,810 | 71,078,420 | 170 | dominated |
| AFRO | Burundi | 60% | 7,172,916 | 492,080 | 3,531 | 1,046 | 150,491 | 84,704,360 | 330 | 6,620 |
| AFRO | Burundi | 70% | 8,361,797 | 487,454 | 3,519 | 999 | 151,445 | 98,330,300 | 2,950 | 14,290 |
| AFRO | Cabo Verde | 0% | - | 120,529 | 1,660 | 270 | - | - | - | - |
| AFRO | Cabo Verde | 20% | 111,389 | 54,085 | 564 | 65 | 2,319 | 1,414,780 | 20 | 610 |
| AFRO | Cabo Verde | 30% | 167,083 | 41,053 | 419 | 61 | 2,447 | 2,053,102 | 50 | dominated |
| AFRO | Cabo Verde | 40% | 222,778 | 33,284 | 331 | 43 | 2,664 | 2,691,424 | 80 | 3,700 |
| AFRO | Cabo Verde | 50% | 278,472 | 28,376 | 271 | 31 | 2,827 | 3,329,746 | 130 | 3,920 |
| AFRO | Cabo Verde | 60% | 334,167 | 26,055 | 263 | 37 | 2,707 | 3,968,068 | 280 | dominated |
| AFRO | Cabo Verde | 70% | 389,861 | 25,087 | 263 | 36 | 2,779 | 4,606,390 | 660 | dominated |
| AFRO | Cameroon | 0% | - | 5,744,471 | 60,788 | 17,998 | - | - | - | - |
| AFRO | Cameroon | 20% | 5,308,469 | 2,751,044 | 22,085 | 2,814 | 202,109 | 67,424,300 | 20 | 330 |
| AFRO | Cameroon | 30% | 7,962,704 | 1,874,502 | 14,016 | 1,487 | 219,998 | 97,844,870 | 30 | 1,700 |
| AFRO | Cameroon | 40% | 10,616,938 | 1,519,569 | 10,645 | 1,115 | 226,585 | 128,265,440 | 90 | 4,620 |
| AFRO | Cameroon | 50% | 13,271,173 | 1,305,457 | 9,715 | 1,009 | 226,767 | 158,686,010 | 140 | 167,320 |
| AFRO | Cameroon | 60% | 15,925,407 | 1,214,752 | 9,344 | 982 | 226,404 | 189,106,580 | 340 | dominated |
| AFRO | Cameroon | 70% | 18,579,642 | 1,159,725 | 9,185 | 1,009 | 226,767 | 219,527,150 | 550 | dominated |
| AFRO | Central African Republic | 0% | - | 987,415 | 10,431 | 4,225 | - | - | - | - |
| AFRO | Central African Republic | 20% | 965,703 | 411,378 | 3,037 | 845 | 42,669 | 12,265,660 | 20 | 290 |
| AFRO | Central African Republic | 30% | 1,448,555 | 305,347 | 2,207 | 604 | 46,305 | 17,799,694 | 50 | 1,520 |
| AFRO | Central African Republic | 40% | 1,915,312 | 265,658 | 1,941 | 415 | 48,981 | 23,333,728 | 140 | 2,070 |
| AFRO | Central African Republic | 50% | 2,398,163 | 231,425 | 1,680 | 328 | 50,169 | 28,867,762 | 160 | 4,660 |
| AFRO | Central African Republic | 60% | 2,881,015 | 209,965 | 1,690 | 406 | 49,130 | 34,401,796 | 260 | dominated |
| AFRO | Central African Republic | 70% | 3,363,867 | 201,278 | 1,579 | 406 | 49,226 | 39,935,830 | 640 | dominated |
| AFRO | Chad | 0% | - | 3,404,190 | 34,380 | 13,042 | - | - | - | - |
| AFRO | Chad | 20% | 3,284,871 | 1,628,556 | 11,515 | 2,365 | 145,474 | 41,722,040 | 20 | 290 |
| AFRO | Chad | 30% | 4,872,559 | 1,163,914 | 6,965 | 1,281 | 164,872 | 60,546,236 | 40 | 970 |
| AFRO | Chad | 40% | 6,514,995 | 880,039 | 6,127 | 608 | 173,696 | 79,370,432 | 70 | 2,130 |
| AFRO | Chad | 50% | 8,157,431 | 772,827 | 5,388 | 641 | 173,449 | 98,194,628 | 180 | dominated |
| AFRO | Chad | 60% | 9,799,867 | 696,035 | 4,747 | 526 | 174,489 | 117,018,824 | 250 | 47,470 |
| AFRO | Chad | 70% | 11,442,302 | 686,180 | 4,879 | 542 | 174,529 | 135,843,020 | 1,910 | 476,300 |
| AFRO | Comoros | 0% | - | 191,384 | 2,214 | 691 | - | - | - | - |
| AFRO | Comoros | 20% | 174,383 | 85,968 | 727 | 91 | 8,369 | 2,214,880 | 20 | 260 |
| AFRO | Comoros | 30% | 261,574 | 61,597 | 463 | 68 | 8,821 | 3,214,192 | 40 | 2,210 |
| AFRO | Comoros | 40% | 351,671 | 46,249 | 338 | 40 | 9,173 | 4,213,504 | 70 | 2,840 |
| AFRO | Comoros | 50% | 438,863 | 42,075 | 316 | 38 | 9,188 | 5,212,816 | 240 | dominated |
| AFRO | Comoros | 60% | 526,054 | 39,164 | 296 | 31 | 9,266 | 6,212,128 | 340 | 21,450 |
| AFRO | Comoros | 70% | 613,245 | 37,213 | 282 | 31 | 9,264 | 7,211,440 | 510 | dominated |
| AFRO | Congo | 0% | - | 1,171,342 | 12,739 | 4,794 | - | - | - | - |

| | | | | | | | | | | |
|---|---|---|---|---|---|---|---|---|---|---|
| AFRO | Congo | 20% | 1,103,290 | 507,465 | 3,956 | 877 | 56,950 | 14,013,180 | 20 | 250 |
| AFRO | Congo | 30% | 1,654,934 | 358,588 | 2,742 | 579 | 61,839 | 20,335,662 | 40 | 1,290 |
| AFRO | Congo | 40% | 2,188,191 | 309,421 | 2,394 | 392 | 64,988 | 26,658,144 | 130 | 2,010 |
| AFRO | Congo | 50% | 2,739,836 | 264,033 | 1,997 | 370 | 65,320 | 32,980,626 | 140 | 19,040 |
| AFRO | Congo | 60% | 3,291,481 | 237,805 | 1,964 | 392 | 64,922 | 39,303,108 | 240 | dominated |
| AFRO | Congo | 70% | 3,843,126 | 229,309 | 1,887 | 348 | 65,489 | 45,625,590 | 740 | 75,050 |
| AFRO | Cote d'Ivoire | 0% | - | 5,711,101 | 63,096 | 23,107 | - | - | - | - |
| AFRO | Cote d'Ivoire | 20% | 5,275,072 | 2,803,955 | 22,632 | 3,772 | 257,217 | 67,000,120 | 20 | 260 |
| AFRO | Cote d'Ivoire | 30% | 7,912,609 | 1,886,423 | 13,769 | 2,216 | 283,401 | 97,229,308 | 30 | 1,150 |
| AFRO | Cote d'Ivoire | 40% | 10,550,145 | 1,547,096 | 12,239 | 1,635 | 290,177 | 127,458,496 | 90 | 4,460 |
| AFRO | Cote d'Ivoire | 50% | 13,187,681 | 1,386,586 | 10,762 | 1,662 | 289,713 | 157,687,684 | dominated | dominated |
| AFRO | Cote d'Ivoire | 60% | 15,825,217 | 1,221,381 | 9,470 | 1,477 | 292,612 | 187,916,872 | 190 | 24,820 |
| AFRO | Cote d'Ivoire | 70% | 18,462,754 | 1,231,510 | 9,681 | 1,345 | 293,285 | 218,146,060 | dominated | 44,930 |
| AFRO | Democratic Repubilc of the C | 0% | - | 18,768,254 | 195,872 | 80,068 | - | - | - | - |
| AFRO | Democratic Repubilc of the C | 20% | 17,910,609 | 7,987,677 | 58,036 | 19,704 | 869,770 | 227,487,480 | 20 | 260 |
| AFRO | Democratic Repubilc of the C | 30% | 26,865,913 | 6,102,486 | 41,199 | 13,166 | 972,808 | 330,125,532 | 50 | 1,000 |
| AFRO | Democratic Repubilc of the C | 40% | 35,821,218 | 4,538,554 | 32,242 | 9,046 | 1,044,351 | 432,763,584 | 70 | 1,430 |
| AFRO | Democratic Repubilc of the C | 50% | 44,776,522 | 4,172,514 | 30,809 | 9,135 | 1,041,879 | 535,401,636 | dominated | dominated |
| AFRO | Democratic Repubilc of the C | 60% | 53,731,826 | 3,764,918 | 28,391 | 7,702 | 1,064,397 | 638,039,688 | 270 | 10,240 |
| AFRO | Democratic Repubilc of the C | 70% | 62,687,131 | 3,677,953 | 28,570 | 7,881 | 1,062,998 | 740,677,740 | 1,180 | dominated |
| AFRO | Eritrea | 0% | - | 760,823 | 9,095 | 3,702 | - | - | - | - |
| AFRO | Eritrea | 20% | 709,129 | 383,053 | 3,312 | 1,206 | 32,971 | 9,006,840 | 20 | 270 |
| AFRO | Eritrea | 30% | 1,063,694 | 257,819 | 1,933 | 610 | 41,964 | 13,070,556 | 30 | 450 |
| AFRO | Eritrea | 40% | 1,430,077 | 208,647 | 1,649 | 486 | 44,034 | 17,134,272 | 80 | 1,960 |
| AFRO | Eritrea | 50% | 1,784,642 | 184,052 | 1,411 | 362 | 45,946 | 21,197,988 | 170 | 2,120 |
| AFRO | Eritrea | 60% | 2,139,206 | 167,197 | 1,376 | 355 | 45,768 | 25,261,704 | 240 | dominated |
| AFRO | Eritrea | 70% | 2,493,771 | 160,485 | 1,333 | 344 | 45,765 | 29,325,420 | 610 | dominated |
| AFRO | Eswatini | 0% | - | 256,376 | 2,988 | 1,142 | - | - | - | - |
| AFRO | Eswatini | 20% | 232,377 | 108,921 | 945 | 217 | 11,902 | 2,951,480 | 20 | 250 |
| AFRO | Eswatini | 30% | 348,565 | 72,661 | 551 | 74 | 13,898 | 4,283,132 | 40 | 670 |
| AFRO | Eswatini | 40% | 464,754 | 62,498 | 451 | 58 | 14,140 | 5,614,784 | dominated | 5,500 |
| AFRO | Eswatini | 50% | 580,942 | 51,267 | 421 | 56 | 14,161 | 6,946,436 | 120 | dominated |
| AFRO | Eswatini | 60% | 697,130 | 47,205 | 360 | 49 | 14,234 | 8,278,088 | 330 | 28,380 |
| AFRO | Eswatini | 70% | 813,319 | 47,213 | 366 | 50 | 14,236 | 9,609,740 | dominated | 496,690 |
| AFRO | Ethiopia | 0% | - | 24,411,376 | 268,896 | 101,511 | - | - | - | - |
| AFRO | Ethiopia | 20% | 22,990,101 | 11,116,940 | 95,648 | 26,326 | 1,072,069 | 292,003,480 | 20 | 270 |
| AFRO | Ethiopia | 30% | 34,485,151 | 7,443,330 | 56,561 | 9,197 | 1,364,352 | 423,749,932 | 40 | 450 |
| AFRO | Ethiopia | 40% | 45,980,202 | 6,277,615 | 45,870 | 5,863 | 1,412,952 | 555,496,384 | 110 | 2,710 |
| AFRO | Ethiopia | 50% | 57,475,252 | 5,403,329 | 40,812 | 5,403 | 1,416,634 | 687,242,836 | 150 | 35,780 |
| AFRO | Ethiopia | 60% | 68,970,302 | 5,130,639 | 38,627 | 5,518 | 1,418,619 | 818,989,288 | 480 | 66,380 |
| AFRO | Ethiopia | 70% | 80,465,353 | 4,993,374 | 38,282 | 5,518 | 1,419,634 | 950,735,740 | 960 | 129,830 |
| AFRO | Gambia | 0% | - | 529,860 | 5,656 | 1,958 | - | - | - | - |
| AFRO | Gambia | 20% | 483,352 | 232,148 | 1,704 | 338 | 23,201 | 6,139,180 | 20 | 260 |
| AFRO | Gambia | 30% | 725,027 | 169,922 | 1,165 | 220 | 24,909 | 8,909,062 | 40 | 1,620 |
| AFRO | Gambia | 40% | 966,703 | 127,915 | 916 | 169 | 25,797 | 11,678,944 | 70 | 3,120 |
| AFRO | Gambia | 50% | 1,208,379 | 108,178 | 771 | 133 | 26,364 | 14,448,826 | 140 | 4,890 |
| AFRO | Gambia | 60% | 1,450,055 | 101,823 | 790 | 140 | 26,237 | 17,218,708 | 440 | dominated |
| AFRO | Gambia | 70% | 1,691,731 | 97,030 | 781 | 143 | 26,216 | 19,988,590 | 580 | dominated |
| AFRO | Ghana | 0% | - | 7,059,135 | 87,555 | 22,464 | - | - | - | - |
| AFRO | Ghana | 20% | 6,213,379 | 3,127,910 | 27,217 | 3,076 | 264,909 | 78,917,800 | 20 | 300 |
| AFRO | Ghana | 30% | 9,320,068 | 2,259,193 | 17,182 | 1,771 | 280,875 | 114,524,020 | 40 | 2,230 |
| AFRO | Ghana | 40% | 12,426,757 | 1,776,955 | 14,199 | 1,771 | 281,529 | 150,130,240 | 70 | dominated |
| AFRO | Ghana | 50% | 15,533,446 | 1,560,957 | 12,335 | 1,367 | 287,465 | 185,736,460 | 160 | 10,810 |

| | | | | | | | | | | |
|---|---|---|---|---|---|---|---|---|---|---|
| AFRO | Ghana | 60% | 18,640,136 | 1,408,838 | 11,247 | 1,522 | 285,353 | 221,342,680 | 230 | dominated |
| AFRO | Ghana | 70% | 21,746,825 | 1,374,723 | 11,154 | 1,491 | 286,890 | 256,948,900 | 1,040 | dominated |
| AFRO | Guinea | 0% | - | 3,007,504 | 31,976 | 13,198 | - | - | - | - |
| AFRO | Guinea | 20% | 2,626,137 | 1,410,127 | 10,847 | 2,876 | 145,109 | 33,355,280 | 20 | 230 |
| AFRO | Guinea | 30% | 3,939,206 | 958,439 | 6,500 | 1,550 | 168,183 | 48,404,552 | 30 | 650 |
| AFRO | Guinea | 40% | 5,296,044 | 821,853 | 5,896 | 1,287 | 171,331 | 63,453,824 | 110 | 4,780 |
| AFRO | Guinea | 50% | 6,609,112 | 709,601 | 5,227 | 1,116 | 173,875 | 78,503,096 | 130 | 5,920 |
| AFRO | Guinea | 60% | 7,922,181 | 647,119 | 4,780 | 1,116 | 173,731 | 93,552,368 | 240 | dominated |
| AFRO | Guinea | 70% | 9,235,250 | 625,871 | 4,754 | 1,195 | 173,019 | 108,601,640 | 710 | dominated |
| AFRO | Guinea Bissau | 0% | - | 443,550 | 4,900 | 921 | - | - | - | - |
| AFRO | Guinea Bissau | 20% | 393,561 | 211,458 | 1,685 | 173 | 8,709 | 4,998,720 | 20 | 570 |
| AFRO | Guinea Bissau | 30% | 590,341 | 157,828 | 1,106 | 122 | 9,532 | 7,254,048 | 40 | 2,740 |
| AFRO | Guinea Bissau | 40% | 787,121 | 125,556 | 895 | 102 | 9,716 | 9,509,376 | 70 | 12,250 |
| AFRO | Guinea Bissau | 50% | 983,902 | 108,580 | 799 | 94 | 9,816 | 11,764,704 | 130 | 22,520 |
| AFRO | Guinea Bissau | 60% | 1,180,682 | 100,248 | 756 | 104 | 9,702 | 14,020,032 | 270 | dominated |
| AFRO | Guinea Bissau | 70% | 1,377,462 | 97,253 | 740 | 98 | 9,765 | 16,275,360 | 750 | dominated |
| AFRO | Kenya | 0% | - | 11,430,906 | 128,515 | 42,211 | - | - | - | - |
| AFRO | Kenya | 20% | 10,753,325 | 5,642,404 | 43,663 | 5,915 | 552,936 | 136,580,880 | 20 | 250 |
| AFRO | Kenya | 30% | 16,129,987 | 4,320,580 | 31,618 | 3,388 | 597,067 | 198,203,592 | 50 | 1,400 |
| AFRO | Kenya | 40% | 21,685,871 | 3,236,107 | 23,230 | 2,366 | 615,665 | 259,826,304 | 60 | 3,310 |
| AFRO | Kenya | 50% | 27,062,533 | 2,865,295 | 21,186 | 1,882 | 621,348 | 321,449,016 | 170 | 10,840 |
| AFRO | Kenya | 60% | 32,439,196 | 2,607,942 | 19,035 | 1,506 | 626,716 | 383,071,728 | 240 | 11,480 |
| AFRO | Kenya | 70% | 37,815,858 | 2,540,458 | 19,250 | 1,828 | 622,364 | 444,694,440 | 910 | dominated |
| AFRO | Lesotho | 0% | - | 458,820 | 6,179 | 2,603 | - | - | - | - |
| AFRO | Lesotho | 20% | 428,757 | 208,783 | 2,000 | 495 | 24,065 | 5,445,760 | 20 | 230 |
| AFRO | Lesotho | 30% | 643,136 | 145,927 | 1,374 | 349 | 26,243 | 7,902,784 | 40 | 1,130 |
| AFRO | Lesotho | 40% | 857,514 | 121,483 | 1,128 | 238 | 27,897 | 10,359,808 | 100 | 1,490 |
| AFRO | Lesotho | 50% | 1,071,893 | 108,997 | 1,044 | 199 | 28,267 | 12,816,832 | 200 | 6,640 |
| AFRO | Lesotho | 60% | 1,286,271 | 101,366 | 973 | 202 | 28,317 | 15,273,856 | 320 | 48,920 |
| AFRO | Lesotho | 70% | 1,500,650 | 97,861 | 991 | 202 | 28,335 | 17,730,880 | 700 | 134,680 |
| AFRO | Liberia | 0% | - | 1,092,701 | 12,299 | 4,339 | - | - | - | - |
| AFRO | Liberia | 20% | 1,011,299 | 515,885 | 4,410 | 819 | 49,845 | 12,844,780 | 20 | 260 |
| AFRO | Liberia | 30% | 1,516,948 | 397,035 | 2,837 | 374 | 56,901 | 18,640,102 | 50 | 820 |
| AFRO | Liberia | 40% | 2,022,598 | 308,391 | 2,154 | 228 | 59,543 | 24,435,424 | 70 | 2,190 |
| AFRO | Liberia | 50% | 2,528,247 | 260,683 | 1,871 | 197 | 59,770 | 30,230,746 | 120 | dominated |
| AFRO | Liberia | 60% | 3,033,897 | 244,238 | 1,790 | 167 | 60,052 | 36,026,068 | 350 | 22,800 |
| AFRO | Liberia | 70% | 3,539,546 | 231,676 | 1,679 | 172 | 60,014 | 41,821,390 | 460 | dominated |
| AFRO | Madagascar | 0% | - | 5,824,758 | 67,121 | 27,247 | - | - | - | - |
| AFRO | Madagascar | 20% | 5,537,446 | 2,968,700 | 24,533 | 6,978 | 289,708 | 70,332,600 | 20 | 240 |
| AFRO | Madagascar | 30% | 8,306,169 | 2,278,749 | 17,666 | 4,237 | 337,554 | 102,065,340 | 50 | 660 |
| AFRO | Madagascar | 40% | 11,167,183 | 1,714,260 | 12,627 | 2,658 | 363,454 | 133,798,080 | 60 | 1,230 |
| AFRO | Madagascar | 50% | 13,935,906 | 1,456,743 | 11,298 | 1,966 | 373,198 | 165,530,820 | 120 | 3,260 |
| AFRO | Madagascar | 60% | 16,704,629 | 1,337,953 | 11,104 | 2,021 | 374,262 | 197,263,560 | 270 | 29,830 |
| AFRO | Madagascar | 70% | 19,473,352 | 1,256,212 | 11,131 | 2,049 | 373,412 | 228,996,300 | 390 | dominated |
| AFRO | Malawi | 0% | - | 4,096,674 | 43,026 | 18,021 | - | - | - | - |
| AFRO | Malawi | 20% | 3,825,817 | 2,002,231 | 15,630 | 5,280 | 176,427 | 48,592,740 | 20 | 280 |
| AFRO | Malawi | 30% | 5,738,726 | 1,480,778 | 10,044 | 3,424 | 204,934 | 70,516,866 | 40 | 770 |
| AFRO | Malawi | 40% | 7,651,635 | 1,134,526 | 7,576 | 2,564 | 220,404 | 92,440,992 | 60 | 1,420 |
| AFRO | Malawi | 50% | 9,564,543 | 1,010,710 | 6,983 | 2,315 | 224,345 | 114,365,118 | 180 | 5,560 |
| AFRO | Malawi | 60% | 11,477,452 | 956,473 | 7,117 | 2,487 | 222,843 | 136,289,244 | 400 | dominated |
| AFRO | Malawi | 70% | 13,390,361 | 922,937 | 6,658 | 2,334 | 225,833 | 158,213,370 | 650 | 29,460 |
| AFRO | Mali | 0% | - | 4,250,604 | 41,494 | 16,019 | - | - | - | - |
| AFRO | Mali | 20% | 4,049,795 | 1,994,116 | 13,467 | 3,443 | 171,252 | 51,437,540 | 20 | 300 |

| | | | | | | | | | | |
|---|---|---|---|---|---|---|---|---|---|---|
| AFRO | Mali | 30% | 6,074,692 | 1,347,036 | 8,505 | 1,539 | 204,680 | 74,645,186 | 40 | 690 |
| AFRO | Mali | 40% | 8,099,590 | 1,112,327 | 7,007 | 972 | 211,288 | 97,852,832 | 100 | 3,510 |
| AFRO | Mali | 50% | 10,124,487 | 943,049 | 6,156 | 1,073 | 209,825 | 121,060,478 | 140 | dominated |
| AFRO | Mali | 60% | 12,149,385 | 830,919 | 5,711 | 1,053 | 210,210 | 144,268,124 | 210 | dominated |
| AFRO | Mali | 70% | 14,174,282 | 833,167 | 5,589 | 1,094 | 209,050 | 167,475,770 | dominated | dominated |
| AFRO | Mauritania | 0% | - | 1,008,362 | 11,667 | 4,892 | - | - | - | - |
| AFRO | Mauritania | 20% | 929,907 | 452,533 | 3,757 | 1,288 | 50,527 | 11,811,000 | 20 | 230 |
| AFRO | Mauritania | 30% | 1,394,860 | 307,709 | 2,432 | 716 | 60,292 | 17,139,900 | 40 | 550 |
| AFRO | Mauritania | 40% | 1,875,312 | 247,092 | 1,823 | 460 | 64,471 | 22,468,800 | 90 | 1,280 |
| AFRO | Mauritania | 50% | 2,340,266 | 209,785 | 1,795 | 409 | 65,524 | 27,797,700 | 140 | 5,060 |
| AFRO | Mauritania | 60% | 2,805,219 | 194,756 | 1,669 | 442 | 64,671 | 33,126,600 | 350 | dominated |
| AFRO | Mauritania | 70% | 3,270,173 | 188,990 | 1,632 | 405 | 65,372 | 38,455,500 | 920 | dominated |
| AFRO | Mozambique | 0% | - | 7,028,518 | 76,051 | 30,289 | - | - | - | - |
| AFRO | Mozambique | 20% | 6,250,975 | 3,235,641 | 25,038 | 6,470 | 319,490 | 79,395,320 | 20 | 250 |
| AFRO | Mozambique | 30% | 9,376,462 | 2,300,745 | 15,629 | 2,532 | 382,245 | 115,216,988 | 40 | 570 |
| AFRO | Mozambique | 40% | 12,397,767 | 1,870,479 | 13,972 | 2,501 | 383,113 | 151,038,656 | 80 | dominated |
| AFRO | Mozambique | 50% | 15,523,254 | 1,618,383 | 11,878 | 2,063 | 389,862 | 186,860,324 | 140 | 9,400 |
| AFRO | Mozambique | 60% | 18,648,742 | 1,468,938 | 11,378 | 1,907 | 390,958 | 222,681,992 | 240 | 32,710 |
| AFRO | Mozambique | 70% | 21,774,229 | 1,415,237 | 10,721 | 1,969 | 389,860 | 258,503,660 | 670 | dominated |
| AFRO | Niger | 0% | - | 5,238,467 | 51,028 | 19,148 | - | - | - | - |
| AFRO | Niger | 20% | 4,840,916 | 2,576,811 | 16,533 | 4,333 | 209,926 | 61,485,780 | 20 | 290 |
| AFRO | Niger | 30% | 7,261,374 | 1,793,303 | 10,457 | 1,985 | 247,246 | 89,227,002 | 40 | 740 |
| AFRO | Niger | 40% | 9,681,832 | 1,503,618 | 9,295 | 1,525 | 252,796 | 116,968,224 | 100 | 5,000 |
| AFRO | Niger | 50% | 12,102,290 | 1,329,037 | 8,521 | 1,477 | 253,629 | 144,709,446 | 160 | 33,310 |
| AFRO | Niger | 60% | 14,522,748 | 1,249,251 | 8,255 | 1,404 | 254,177 | 172,450,668 | 350 | 50,660 |
| AFRO | Niger | 70% | 16,943,206 | 1,182,996 | 7,795 | 1,428 | 253,764 | 200,191,890 | 420 | dominated |
| AFRO | Nigeria | 0% | - | 44,250,442 | 453,100 | 187,795 | - | - | - | - |
| AFRO | Nigeria | 20% | 41,224,277 | 19,430,739 | 148,835 | 48,443 | 1,820,390 | 523,600,680 | 20 | 290 |
| AFRO | Nigeria | 30% | 61,836,416 | 15,136,389 | 108,018 | 25,562 | 2,182,714 | 759,839,412 | 60 | 650 |
| AFRO | Nigeria | 40% | 82,448,554 | 11,036,430 | 76,479 | 15,048 | 2,319,951 | 996,078,144 | 60 | 1,720 |
| AFRO | Nigeria | 50% | 103,060,693 | 9,535,923 | 63,698 | 14,018 | 2,345,935 | 1,232,316,876 | 160 | 9,090 |
| AFRO | Nigeria | 60% | 123,672,831 | 8,610,345 | 65,553 | 13,605 | 2,357,045 | 1,468,555,608 | 260 | 21,260 |
| AFRO | Nigeria | 70% | 144,284,970 | 8,480,476 | 64,729 | 12,781 | 2,358,652 | 1,704,794,340 | 1,820 | 146,990 |
| AFRO | Rwanda | 0% | - | 2,871,005 | 32,872 | 13,017 | - | - | - | - |
| AFRO | Rwanda | 20% | 2,590,141 | 1,271,835 | 10,944 | 3,044 | 143,811 | 32,898,080 | 20 | 230 |
| AFRO | Rwanda | 30% | 3,885,211 | 854,923 | 7,033 | 1,541 | 170,408 | 47,741,072 | 40 | 560 |
| AFRO | Rwanda | 40% | 5,180,282 | 685,549 | 5,336 | 1,088 | 177,300 | 62,584,064 | 90 | 2,150 |
| AFRO | Rwanda | 50% | 6,475,352 | 581,752 | 4,896 | 997 | 179,315 | 77,427,056 | 140 | 7,370 |
| AFRO | Rwanda | 60% | 7,770,423 | 566,857 | 4,792 | 958 | 179,229 | 92,270,048 | dominated | dominated |
| AFRO | Rwanda | 70% | 9,065,493 | 540,176 | 4,598 | 958 | 178,950 | 107,113,040 | 710 | dominated |
| AFRO | Senegal | 0% | - | 3,821,734 | 42,881 | 17,213 | - | - | - | - |
| AFRO | Senegal | 20% | 3,348,465 | 1,882,645 | 15,003 | 5,526 | 163,221 | 42,529,760 | 20 | 260 |
| AFRO | Senegal | 30% | 5,022,698 | 1,505,185 | 10,180 | 3,198 | 200,510 | 61,718,384 | 50 | 510 |
| AFRO | Senegal | 40% | 6,696,930 | 1,170,188 | 8,757 | 2,796 | 208,410 | 80,907,008 | 60 | dominated |
| AFRO | Senegal | 50% | 8,371,163 | 979,993 | 7,183 | 2,043 | 219,955 | 100,095,632 | 100 | 1,970 |
| AFRO | Senegal | 60% | 10,045,395 | 965,476 | 7,049 | 2,026 | 219,681 | 119,284,256 | dominated | dominated |
| AFRO | Senegal | 70% | 11,719,628 | 918,660 | 6,915 | 1,959 | 219,796 | 138,472,880 | 630 | dominated |
| AFRO | Sierra Leone | 0% | - | 1,715,327 | 19,315 | 7,512 | - | - | - | - |
| AFRO | Sierra Leone | 20% | 1,594,840 | 755,384 | 6,205 | 1,523 | 77,144 | 20,256,500 | 20 | 260 |
| AFRO | Sierra Leone | 30% | 2,392,261 | 555,778 | 4,091 | 726 | 89,125 | 29,395,850 | 50 | 760 |
| AFRO | Sierra Leone | 40% | 3,216,262 | 419,246 | 2,967 | 471 | 92,661 | 38,535,200 | 70 | 2,580 |
| AFRO | Sierra Leone | 50% | 4,013,682 | 376,053 | 2,719 | 423 | 93,233 | 47,674,550 | 210 | 15,960 |
| AFRO | Sierra Leone | 60% | 4,811,102 | 348,476 | 2,616 | 471 | 92,581 | 56,813,900 | 330 | dominated |

| | | | | | | | | | | |
|---|---|---|---|---|---|---|---|---|---|---|
| AFRO | Sierra Leone | 70% | 5,608,522 | 337,669 | 2,584 | 486 | 92,230 | 65,953,250 | 850 | dominated |
| AFRO | Sao Tome and Principe | 0% | - | 49,986 | 546 | 153 | - | - | - | - |
| AFRO | Sao Tome and Principe | 20% | 43,996 | 24,713 | 205 | 26 | 1,739 | 558,800 | 20 | 320 |
| AFRO | Sao Tome and Principe | 30% | 65,993 | 16,744 | 125 | 12 | 1,917 | 810,920 | 30 | 1,410 |
| AFRO | Sao Tome and Principe | 40% | 87,991 | 13,902 | 111 | 15 | 1,902 | 1,063,040 | 90 | dominated |
| AFRO | Sao Tome and Principe | 50% | 109,989 | 12,077 | 96 | 13 | 1,895 | 1,315,160 | 140 | dominated |
| AFRO | Sao Tome and Principe | 60% | 131,987 | 11,014 | 88 | 12 | 1,914 | 1,567,280 | 240 | dominated |
| AFRO | Sao Tome and Principe | 70% | 153,985 | 10,608 | 87 | 10 | 1,929 | 1,819,400 | 620 | 85,670 |
| AFRO | South Sudan | 0% | - | 2,538,645 | 28,593 | 9,837 | - | - | - | - |
| AFRO | South Sudan | 20% | 2,237,976 | 1,089,041 | 8,707 | 1,421 | 115,821 | 28,425,140 | 20 | 250 |
| AFRO | South Sudan | 30% | 3,356,964 | 798,601 | 5,886 | 772 | 126,222 | 41,250,026 | 40 | 1,230 |
| AFRO | South Sudan | 40% | 4,475,952 | 655,244 | 4,835 | 660 | 128,014 | 54,074,912 | 90 | 7,150 |
| AFRO | South Sudan | 50% | 5,594,940 | 577,512 | 4,275 | 582 | 128,963 | 66,899,798 | 160 | 13,520 |
| AFRO | South Sudan | 60% | 6,713,929 | 511,977 | 3,805 | 537 | 129,176 | 79,724,684 | 200 | 60,130 |
| AFRO | South Sudan | 70% | 7,832,917 | 498,111 | 4,006 | 526 | 128,936 | 92,549,570 | 920 | dominated |
| AFRO | Tanzania | 0% | - | 11,686,751 | 119,173 | 50,537 | - | - | - | - |
| AFRO | Tanzania | 20% | 11,946,005 | 5,746,424 | 42,174 | 15,233 | 518,901 | 151,729,440 | 30 | 290 |
| AFRO | Tanzania | 30% | 17,919,008 | 4,226,083 | 30,824 | 9,558 | 609,584 | 220,186,896 | 50 | 750 |
| AFRO | Tanzania | 40% | 23,892,011 | 3,193,964 | 23,178 | 7,467 | 644,002 | 288,644,352 | 70 | 1,990 |
| AFRO | Tanzania | 50% | 29,865,013 | 2,731,966 | 21,087 | 6,093 | 665,235 | 357,101,808 | 150 | 3,220 |
| AFRO | Tanzania | 60% | 35,838,016 | 2,515,125 | 19,952 | 6,153 | 664,771 | 425,559,264 | 320 | dominated |
| AFRO | Tanzania | 70% | 41,811,018 | 2,458,674 | 19,892 | 6,093 | 665,987 | 494,016,720 | 1,210 | 181,920 |
| AFRO | Togo | 0% | - | 1,791,047 | 20,129 | 7,551 | - | - | - | - |
| AFRO | Togo | 20% | 1,655,834 | 788,836 | 6,715 | 1,374 | 85,705 | 21,031,200 | 20 | 250 |
| AFRO | Togo | 30% | 2,483,752 | 544,716 | 3,983 | 530 | 99,202 | 30,520,080 | 40 | 700 |
| AFRO | Togo | 40% | 3,311,669 | 459,151 | 3,478 | 373 | 100,360 | 40,008,960 | 110 | 8,190 |
| AFRO | Togo | 50% | 4,139,586 | 379,530 | 3,055 | 397 | 100,755 | 49,497,840 | 120 | dominated |
| AFRO | Togo | 60% | 4,967,503 | 361,844 | 2,848 | 331 | 101,767 | 58,986,720 | dominated | 13,490 |
| AFRO | Togo | 70% | 5,795,420 | 334,827 | 2,807 | 306 | 101,947 | 68,475,600 | 420 | 52,660 |
| AFRO | Uganda | 0% | - | 8,807,064 | 81,556 | 33,437 | - | - | - | - |
| AFRO | Uganda | 20% | 9,147,285 | 3,806,246 | 27,490 | 8,371 | 382,063 | 116,182,140 | 20 | 300 |
| AFRO | Uganda | 30% | 13,720,928 | 2,728,405 | 17,793 | 4,437 | 448,638 | 168,601,326 | 50 | 790 |
| AFRO | Uganda | 40% | 18,294,570 | 2,370,848 | 15,277 | 4,208 | 455,745 | 221,020,512 | dominated | dominated |
| AFRO | Uganda | 50% | 22,868,213 | 1,893,815 | 14,225 | 3,659 | 461,345 | 273,439,698 | 130 | dominated |
| AFRO | Uganda | 60% | 27,441,856 | 1,764,276 | 12,990 | 2,836 | 474,960 | 325,858,884 | 400 | 5,970 |
| AFRO | Uganda | 70% | 32,015,498 | 1,717,712 | 12,945 | 2,790 | 475,276 | 378,278,070 | 1,130 | 165,570 |
| AFRO | Zambia | 0% | - | 4,065,420 | 40,392 | 14,120 | - | - | - | - |
| AFRO | Zambia | 20% | 3,676,632 | 1,644,060 | 12,575 | 2,041 | 184,212 | 46,697,900 | 20 | 250 |
| AFRO | Zambia | 30% | 5,514,948 | 1,175,132 | 8,181 | 974 | 201,310 | 67,767,110 | 40 | 1,230 |
| AFRO | Zambia | 40% | 7,353,265 | 972,254 | 6,839 | 588 | 207,413 | 88,836,320 | 100 | 3,450 |
| AFRO | Zambia | 50% | 9,191,581 | 842,768 | 6,398 | 791 | 204,926 | 109,905,530 | 160 | dominated |
| AFRO | Zambia | 60% | 11,029,897 | 766,875 | 5,828 | 699 | 205,638 | 130,974,740 | 280 | dominated |
| AFRO | Zambia | 70% | 12,868,213 | 746,596 | 5,699 | 588 | 207,419 | 152,043,950 | 1,040 | 11,605,160 |
| AFRO | Zimbabwe | 0% | - | 3,327,112 | 37,128 | 15,086 | - | - | - | - |
| AFRO | Zimbabwe | 20% | 2,972,303 | 1,584,767 | 12,871 | 4,147 | 145,624 | 37,752,020 | 20 | 260 |
| AFRO | Zimbabwe | 30% | 4,458,454 | 1,150,337 | 8,457 | 2,125 | 178,111 | 54,785,018 | 40 | 520 |
| AFRO | Zimbabwe | 40% | 5,944,605 | 967,002 | 6,837 | 1,858 | 181,720 | 71,818,016 | 90 | 4,720 |
| AFRO | Zimbabwe | 50% | 7,430,757 | 848,796 | 6,569 | 1,828 | 182,413 | 88,851,014 | 140 | 24,590 |
| AFRO | Zimbabwe | 60% | 8,916,908 | 771,196 | 5,841 | 1,784 | 181,648 | 105,884,012 | 220 | dominated |
| AFRO | Zimbabwe | 70% | 10,403,060 | 753,658 | 5,707 | 1,754 | 182,935 | 122,917,010 | 970 | 65,340 |
| EMRO | All countries | 0% | - | 113,700,934 | 1,456,853 | 407,648 | - | - | - | - |
| EMRO | All countries | 20% | 104,797,802 | 53,705,690 | 499,688 | 75,254 | 4,348,439 | 1,331,065,197 | 20 | 310 |
| EMRO | All countries | 30% | 157,196,703 | 37,249,365 | 327,614 | 48,439 | 4,785,496 | 1,931,616,659 | 40 | 1,370 |

| | | | | | | | | | | |
|---|---|---|---|---|---|---|---|---|---|---|
| EMRO | All countries | 40% | 209,741,754 | 30,515,817 | 251,677 | 40,810 | 4,891,803 | 2,532,168,122 | 90 | 5,650 |
| EMRO | All countries | 50% | 262,195,691 | 26,069,014 | 231,007 | 36,331 | 4,936,122 | 3,132,719,585 | 140 | 13,550 |
| EMRO | All countries | 60% | 314,594,593 | 24,173,943 | 222,273 | 35,956 | 4,946,319 | 3,733,271,048 | 320 | dominated |
| EMRO | All countries | 70% | 366,993,493 | 23,368,073 | 213,526 | 34,110 | 4,975,428 | 4,333,822,510 | 750 | 30,560 |
| EMRO | Afghanistan | 0% | - | 8,773,562 | 93,503 | 37,993 | - | - | - | - |
| EMRO | Afghanistan | 20% | 7,784,621 | 3,955,178 | 30,558 | 10,160 | 411,636 | 98,874,580 | 20 | 240 |
| EMRO | Afghanistan | 30% | 11,676,932 | 2,844,162 | 18,841 | 5,372 | 486,982 | 143,484,922 | 40 | 590 |
| EMRO | Afghanistan | 40% | 15,569,243 | 2,278,748 | 16,388 | 4,866 | 497,053 | 188,095,264 | 80 | dominated |
| EMRO | Afghanistan | 50% | 19,461,554 | 1,905,126 | 14,208 | 3,815 | 512,074 | 232,705,606 | 120 | 3,560 |
| EMRO | Afghanistan | 60% | 23,353,864 | 1,808,198 | 13,547 | 3,776 | 512,326 | 277,315,948 | dominated | 177,320 |
| EMRO | Afghanistan | 70% | 27,246,175 | 1,703,017 | 13,430 | 3,893 | 508,053 | 321,926,290 | 440 | dominated |
| EMRO | Djibouti | 0% | - | 212,095 | 2,934 | 1,379 | - | - | - | - |
| EMRO | Djibouti | 20% | 197,380 | 91,291 | 944 | 408 | 13,622 | 2,506,980 | 20 | 180 |
| EMRO | Djibouti | 30% | 296,070 | 66,264 | 693 | 310 | 15,298 | 3,638,082 | 50 | 670 |
| EMRO | Djibouti | 40% | 394,761 | 54,860 | 513 | 210 | 16,924 | 4,769,184 | 100 | 700 |
| EMRO | Djibouti | 50% | 490,161 | 49,850 | 518 | 211 | 16,980 | 5,900,286 | 230 | dominated |
| EMRO | Djibouti | 60% | 588,851 | 45,047 | 463 | 191 | 17,313 | 7,031,388 | 240 | 5,820 |
| EMRO | Djibouti | 70% | 687,541 | 44,379 | 467 | 201 | 17,190 | 8,162,490 | 1,690 | dominated |
| EMRO | Egypt | 0% | - | 21,806,352 | 302,295 | 44,515 | - | - | - | - |
| EMRO | Egypt | 20% | 20,464,753 | 9,669,540 | 100,390 | 13,201 | 325,925 | 259,928,360 | 20 | 800 |
| EMRO | Egypt | 30% | 30,697,130 | 6,780,549 | 65,801 | 8,187 | 396,335 | 377,203,124 | 40 | 1,670 |
| EMRO | Egypt | 40% | 40,929,507 | 5,517,133 | 49,325 | 6,038 | 416,738 | 494,477,888 | 90 | 5,750 |
| EMRO | Egypt | 50% | 51,161,883 | 4,812,256 | 50,655 | 6,959 | 405,044 | 611,752,652 | 170 | dominated |
| EMRO | Egypt | 60% | 61,394,260 | 4,299,768 | 44,515 | 6,242 | 415,029 | 729,027,416 | 230 | dominated |
| EMRO | Egypt | 70% | 71,626,637 | 4,195,489 | 43,799 | 5,526 | 420,642 | 846,302,180 | 1,120 | 90,110 |
| EMRO | Morocco | 0% | - | 7,767,440 | 124,021 | 29,640 | - | - | - | - |
| EMRO | Morocco | 20% | 7,381,462 | 3,615,838 | 42,854 | 5,906 | 281,762 | 93,753,940 | 20 | 330 |
| EMRO | Morocco | 30% | 11,072,193 | 2,614,406 | 28,643 | 4,503 | 302,653 | 136,053,946 | 40 | 2,020 |
| EMRO | Morocco | 40% | 14,762,924 | 2,080,230 | 21,630 | 3,507 | 316,986 | 178,353,952 | 80 | 2,950 |
| EMRO | Morocco | 50% | 18,453,654 | 1,683,695 | 18,898 | 2,805 | 325,089 | 220,653,958 | 110 | 5,220 |
| EMRO | Morocco | 60% | 22,144,385 | 1,620,799 | 17,053 | 2,399 | 330,229 | 262,953,964 | dominated | 8,230 |
| EMRO | Morocco | 70% | 25,835,116 | 1,552,920 | 16,241 | 2,252 | 333,561 | 305,253,970 | 650 | 12,690 |
| EMRO | Pakistan | 0% | - | 47,556,717 | 601,492 | 191,514 | - | - | - | - |
| EMRO | Pakistan | 20% | 44,174,182 | 23,798,570 | 212,499 | 22,089 | 2,241,306 | 561,068,220 | 20 | 250 |
| EMRO | Pakistan | 30% | 66,261,273 | 15,838,691 | 136,954 | 14,800 | 2,377,961 | 814,211,598 | 30 | 1,850 |
| EMRO | Pakistan | 40% | 88,348,364 | 13,352,540 | 103,820 | 14,579 | 2,377,596 | 1,067,354,976 | 100 | dominated |
| EMRO | Pakistan | 50% | 110,435,455 | 11,473,624 | 92,775 | 11,707 | 2,395,725 | 1,320,498,354 | 130 | 28,500 |
| EMRO | Pakistan | 60% | 132,522,546 | 10,710,218 | 96,088 | 13,254 | 2,380,777 | 1,573,641,732 | 330 | dominated |
| EMRO | Pakistan | 70% | 154,609,637 | 10,343,977 | 89,462 | 12,591 | 2,398,060 | 1,826,785,110 | 690 | 216,850 |
| EMRO | Somalia | 0% | - | 3,416,074 | 33,999 | 13,829 | - | - | - | - |
| EMRO | Somalia | 20% | 3,178,682 | 1,356,829 | 9,585 | 3,290 | 144,458 | 40,373,300 | 20 | 280 |
| EMRO | Somalia | 30% | 4,768,023 | 956,386 | 6,183 | 1,892 | 168,408 | 58,588,970 | 50 | 760 |
| EMRO | Somalia | 40% | 6,357,364 | 801,966 | 4,912 | 1,494 | 173,899 | 76,804,640 | 120 | 3,320 |
| EMRO | Somalia | 50% | 7,946,705 | 670,578 | 4,768 | 1,367 | 174,883 | 95,020,310 | 140 | 18,520 |
| EMRO | Somalia | 60% | 9,536,046 | 622,210 | 4,562 | 1,526 | 172,024 | 113,235,980 | 380 | dominated |
| EMRO | Somalia | 70% | 11,125,387 | 587,225 | 4,387 | 1,367 | 174,599 | 131,451,650 | 520 | dominated |
| EMRO | Sudan | 0% | - | 9,921,626 | 113,920 | 49,067 | - | - | - | - |
| EMRO | Sudan | 20% | 8,768,923 | 4,536,574 | 37,622 | 12,497 | 515,274 | 111,376,460 | 20 | 220 |
| EMRO | Sudan | 30% | 13,153,385 | 3,152,348 | 25,125 | 7,454 | 595,379 | 161,627,414 | 40 | 630 |
| EMRO | Sudan | 40% | 17,683,995 | 2,553,590 | 20,302 | 5,350 | 631,808 | 211,878,368 | 80 | 1,380 |
| EMRO | Sudan | 50% | 22,068,456 | 2,112,601 | 17,978 | 5,262 | 638,420 | 262,129,322 | 110 | dominated |
| EMRO | Sudan | 60% | 26,452,918 | 1,978,949 | 16,575 | 4,429 | 648,985 | 312,380,276 | 380 | 5,850 |
| EMRO | Sudan | 70% | 30,837,379 | 1,953,298 | 16,575 | 4,078 | 653,760 | 362,631,230 | 1,960 | 10,530 |

| | | | | | | | | | | |
|---|---|---|---|---|---|---|---|---|---|---|
| EMRO | Syrian Arab Republic | 0% | - | 3,798,025 | 51,520 | 15,522 | - | - | - | - |
| EMRO | Syrian Arab Republic | 20% | 3,499,650 | 1,908,550 | 19,390 | 2,468 | 187,884 | 44,450,000 | 20 | 240 |
| EMRO | Syrian Arab Republic | 30% | 5,249,475 | 1,389,885 | 14,070 | 1,855 | 196,160 | 64,505,000 | 40 | 2,420 |
| EMRO | Syrian Arab Republic | 40% | 6,999,300 | 1,131,025 | 11,130 | 1,488 | 201,795 | 84,560,000 | 80 | 3,560 |
| EMRO | Syrian Arab Republic | 50% | 8,807,452 | 971,478 | 9,502 | 1,120 | 207,001 | 104,615,000 | 130 | 3,850 |
| EMRO | Syrian Arab Republic | 60% | 10,557,278 | 865,165 | 8,960 | 1,120 | 208,009 | 124,670,000 | 190 | 19,900 |
| EMRO | Syrian Arab Republic | 70% | 12,307,102 | 839,510 | 8,872 | 1,190 | 206,835 | 144,725,000 | 780 | dominated |
| EMRO | Tunisia | 0% | - | 2,645,896 | 47,950 | 12,044 | - | - | - | - |
| EMRO | Tunisia | 20% | 2,363,564 | 1,259,681 | 16,145 | 2,565 | 110,742 | 30,020,260 | 20 | 270 |
| EMRO | Tunisia | 30% | 3,545,345 | 961,050 | 11,429 | 1,726 | 121,512 | 43,564,834 | 50 | 1,260 |
| EMRO | Tunisia | 40% | 4,727,127 | 736,028 | 8,900 | 1,560 | 125,034 | 57,109,408 | 60 | 3,850 |
| EMRO | Tunisia | 50% | 5,908,909 | 646,393 | 7,895 | 1,359 | 128,216 | 70,653,982 | 150 | 4,260 |
| EMRO | Tunisia | 60% | 7,090,691 | 587,310 | 7,481 | 1,253 | 129,231 | 84,198,556 | 230 | 13,350 |
| EMRO | Tunisia | 70% | 8,272,473 | 583,859 | 7,541 | 1,359 | 128,473 | 97,743,130 | 3,920 | dominated |
| EMRO | West Bank and Gaza | 0% | - | 1,148,921 | 12,565 | 1,617 | - | - | - | - |
| EMRO | West Bank and Gaza | 20% | 1,020,181 | 515,570 | 4,469 | 403 | 15,777 | 12,957,597 | 20 | 820 |
| EMRO | West Bank and Gaza | 30% | 1,530,272 | 399,772 | 3,173 | 342 | 17,080 | 18,803,819 | 50 | 4,490 |
| EMRO | West Bank and Gaza | 40% | 2,040,362 | 326,404 | 2,648 | 316 | 17,365 | 24,650,042 | 80 | dominated |
| EMRO | West Bank and Gaza | 50% | 2,550,453 | 281,302 | 2,357 | 235 | 18,150 | 30,496,265 | 130 | 10,930 |
| EMRO | West Bank and Gaza | 60% | 3,060,544 | 267,789 | 2,173 | 275 | 17,857 | 36,342,488 | 430 | dominated |
| EMRO | West Bank and Gaza | 70% | 3,570,634 | 257,229 | 2,224 | 281 | 17,816 | 42,188,710 | 550 | dominated |
| EMRO | Yemen | 0% | - | 6,654,226 | 72,654 | 10,528 | - | - | - | - |
| EMRO | Yemen | 20% | 5,964,404 | 2,998,069 | 25,232 | 2,267 | 100,054 | 75,755,500 | 20 | 760 |
| EMRO | Yemen | 30% | 8,946,605 | 2,245,852 | 16,702 | 1,998 | 107,729 | 109,934,950 | 50 | dominated |
| EMRO | Yemen | 40% | 11,928,807 | 1,683,293 | 12,109 | 1,402 | 116,605 | 144,114,400 | 60 | 4,130 |
| EMRO | Yemen | 50% | 14,911,009 | 1,462,111 | 11,453 | 1,491 | 114,539 | 178,293,850 | 150 | dominated |
| EMRO | Yemen | 60% | 17,893,210 | 1,368,490 | 10,856 | 1,491 | 114,539 | 212,473,300 | 370 | dominated |
| EMRO | Yemen | 70% | 20,875,412 | 1,307,170 | 10,528 | 1,372 | 116,441 | 246,652,750 | 560 | dominated |
| EURO | All countries | 0% | - | 21,559,264 | 401,351 | 102,741 | - | - | - | - |
| EURO | All countries | 20% | 19,816,468 | 10,258,273 | 137,413 | 23,231 | 887,886 | 251,694,310 | 20 | 280 |
| EURO | All countries | 30% | 29,724,702 | 7,323,175 | 92,528 | 15,017 | 998,742 | 365,254,026 | 40 | 1,020 |
| EURO | All countries | 40% | 39,632,935 | 5,868,565 | 74,347 | 12,791 | 1,026,646 | 478,813,742 | 80 | 4,070 |
| EURO | All countries | 50% | 49,429,620 | 5,054,107 | 66,660 | 11,496 | 1,042,002 | 592,373,459 | 140 | 7,400 |
| EURO | All countries | 60% | 59,337,854 | 4,712,484 | 63,100 | 10,202 | 1,056,815 | 705,933,175 | 330 | 7,670 |
| EURO | All countries | 70% | 69,246,088 | 4,579,267 | 62,104 | 10,234 | 1,057,413 | 819,492,891 | 850 | 189,830 |
| EURO | Kosovo | 0% | - | 376,327 | 8,135 | 2,110 | - | - | - | - |
| EURO | Kosovo | 20% | 358,814 | 200,674 | 3,068 | 488 | 17,406 | 4,557,390 | 30 | 260 |
| EURO | Kosovo | 30% | 538,221 | 142,501 | 2,096 | 402 | 18,710 | 6,613,598 | 40 | 1,580 |
| EURO | Kosovo | 40% | 717,627 | 114,118 | 1,744 | 321 | 19,818 | 8,669,806 | 70 | 1,860 |
| EURO | Kosovo | 50% | 897,034 | 98,515 | 1,518 | 289 | 20,124 | 10,726,015 | 130 | dominated |
| EURO | Kosovo | 60% | 1,076,441 | 90,366 | 1,373 | 251 | 20,574 | 12,782,223 | 250 | 5,440 |
| EURO | Kosovo | 70% | 1,255,848 | 87,109 | 1,371 | 257 | 20,523 | 14,838,431 | 630 | dominated |
| EURO | Kyrgyzstan | 0% | - | 1,380,918 | 19,549 | 7,014 | - | - | - | - |
| EURO | Kyrgyzstan | 20% | 1,304,870 | 677,641 | 7,158 | 1,103 | 76,267 | 16,573,500 | 20 | 220 |
| EURO | Kyrgyzstan | 30% | 1,957,304 | 490,745 | 4,639 | 620 | 82,745 | 24,051,150 | 40 | 1,150 |
| EURO | Kyrgyzstan | 40% | 2,609,739 | 415,329 | 3,687 | 444 | 85,167 | 31,528,800 | 100 | 3,090 |
| EURO | Kyrgyzstan | 50% | 3,262,174 | 348,481 | 3,367 | 365 | 86,462 | 39,006,450 | 110 | 5,780 |
| EURO | Kyrgyzstan | 60% | 3,914,608 | 334,452 | 3,302 | 418 | 85,651 | 46,484,100 | 530 | dominated |
| EURO | Kyrgyzstan | 70% | 4,567,043 | 321,369 | 3,223 | 372 | 86,335 | 53,961,750 | 570 | dominated |
| EURO | Moldova | 0% | - | 858,697 | 18,052 | 3,352 | - | - | - | - |
| EURO | Moldova | 20% | 806,719 | 381,048 | 5,716 | 1,017 | 20,987 | 10,246,360 | 20 | 490 |
| EURO | Moldova | 30% | 1,210,079 | 277,555 | 4,324 | 662 | 25,372 | 14,869,324 | 40 | 1,050 |
| EURO | Moldova | 40% | 1,613,439 | 215,658 | 3,243 | 597 | 25,856 | 19,492,288 | 70 | dominated |

| | | | | | | | | | | |
|---|---|---|---|---|---|---|---|---|---|---|
| EURO | Moldova | 50% | 2,016,798 | 188,331 | 2,872 | 532 | 26,948 | 24,115,252 | 170 | 5,870 |
| EURO | Moldova | 60% | 2,420,158 | 176,750 | 2,650 | 460 | 27,543 | 28,738,216 | 400 | 7,770 |
| EURO | Moldova | 70% | 2,823,518 | 169,251 | 2,622 | 416 | 27,876 | 33,361,180 | 620 | 13,900 |
| EURO | Tajikistan | 0% | - | 1,969,454 | 23,225 | 2,928 | - | - | - | - |
| EURO | Tajikistan | 20% | 1,907,409 | 958,340 | 8,508 | 1,030 | 21,721 | 24,226,520 | 20 | 1,120 |
| EURO | Tajikistan | 30% | 2,861,114 | 691,534 | 5,971 | 668 | 26,211 | 35,157,068 | 40 | 2,430 |
| EURO | Tajikistan | 40% | 3,814,818 | 546,356 | 4,407 | 496 | 28,653 | 46,087,616 | 80 | 4,470 |
| EURO | Tajikistan | 50% | 4,768,523 | 453,952 | 3,720 | 429 | 29,374 | 57,018,164 | 120 | 15,170 |
| EURO | Tajikistan | 60% | 5,722,228 | 412,080 | 3,539 | 391 | 29,454 | 67,948,712 | 260 | dominated |
| EURO | Tajikistan | 70% | 6,675,932 | 395,388 | 3,434 | 353 | 30,163 | 78,879,260 | 650 | 27,710 |
| EURO | Ukraine | 0% | - | 9,305,814 | 216,959 | 43,427 | - | - | - | - |
| EURO | Ukraine | 20% | 8,745,725 | 4,302,496 | 71,329 | 12,464 | 255,220 | 111,081,820 | 20 | 440 |
| EURO | Ukraine | 30% | 13,118,588 | 3,094,941 | 48,456 | 9,184 | 293,558 | 161,199,838 | 40 | 1,310 |
| EURO | Ukraine | 40% | 17,491,451 | 2,438,465 | 39,010 | 7,653 | 309,849 | 211,317,856 | 80 | 3,080 |
| EURO | Ukraine | 50% | 21,864,313 | 2,125,293 | 35,336 | 6,735 | 320,742 | 261,435,874 | 160 | 4,600 |
| EURO | Ukraine | 60% | 26,237,176 | 1,979,049 | 33,193 | 5,904 | 330,525 | 311,553,892 | 340 | 5,120 |
| EURO | Ukraine | 70% | 30,610,039 | 1,927,663 | 32,712 | 5,991 | 330,351 | 361,671,910 | 980 | dominated |
| EURO | Uzbekistan | 0% | - | 7,668,054 | 115,431 | 43,910 | - | - | - | - |
| EURO | Uzbekistan | 20% | 6,692,931 | 3,738,074 | 41,634 | 7,129 | 496,284 | 85,008,720 | 20 | 170 |
| EURO | Uzbekistan | 30% | 10,039,396 | 2,625,899 | 27,042 | 3,481 | 552,146 | 123,363,048 | 30 | 690 |
| EURO | Uzbekistan | 40% | 13,385,861 | 2,138,639 | 22,256 | 3,280 | 557,303 | 161,717,376 | 80 | 7,440 |
| EURO | Uzbekistan | 50% | 16,620,778 | 1,839,535 | 19,847 | 3,146 | 558,352 | 200,071,704 | 130 | dominated |
| EURO | Uzbekistan | 60% | 19,967,243 | 1,719,787 | 19,043 | 2,778 | 563,068 | 238,426,032 | 320 | 13,310 |
| EURO | Uzbekistan | 70% | 23,313,708 | 1,678,487 | 18,742 | 2,845 | 562,166 | 276,780,360 | 930 | dominated |
| PAHO | All countries | 0% | - | 10,084,834 | 146,861 | 49,666 | - | - | - | - |
| PAHO | All countries | 20% | 9,470,252 | 4,791,473 | 50,296 | 8,821 | 568,339 | 120,284,217 | 20 | 210 |
| PAHO | All countries | 30% | 14,205,377 | 3,501,739 | 35,289 | 5,055 | 630,825 | 174,554,183 | 40 | 870 |
| PAHO | All countries | 40% | 18,940,503 | 2,831,701 | 28,220 | 4,242 | 644,220 | 228,824,149 | 80 | 4,050 |
| PAHO | All countries | 50% | 23,603,858 | 2,430,728 | 24,713 | 3,692 | 651,291 | 283,094,114 | 140 | 7,670 |
| PAHO | All countries | 60% | 28,338,984 | 2,241,631 | 23,182 | 3,302 | 656,017 | 337,364,080 | 290 | 11,480 |
| PAHO | All countries | 70% | 33,074,109 | 2,158,338 | 22,843 | 3,288 | 655,949 | 391,634,046 | 650 | dominated |
| PAHO | Bolivia | 0% | - | 2,267,153 | 33,735 | 12,362 | - | - | - | - |
| PAHO | Bolivia | 20% | 2,334,367 | 1,072,235 | 11,661 | 2,008 | 147,011 | 29,649,420 | 20 | 200 |
| PAHO | Bolivia | 30% | 3,501,550 | 766,671 | 8,078 | 1,202 | 161,202 | 43,026,678 | 40 | 940 |
| PAHO | Bolivia | 40% | 4,668,733 | 623,863 | 6,432 | 1,039 | 164,567 | 56,403,936 | 90 | 3,980 |
| PAHO | Bolivia | 50% | 5,835,916 | 511,593 | 5,673 | 864 | 166,262 | 69,781,194 | 120 | 7,890 |
| PAHO | Bolivia | 60% | 7,003,100 | 474,414 | 5,346 | 852 | 166,239 | 83,158,452 | 360 | dominated |
| PAHO | Bolivia | 70% | 8,170,283 | 464,002 | 5,241 | 794 | 166,940 | 96,535,710 | 1,280 | 39,450 |
| PAHO | Dominica | 0% | - | 16,315 | 298 | 69 | - | - | - | - |
| PAHO | Dominica | 20% | 14,397 | 8,048 | 105 | 17 | 585 | 182,857 | 20 | 310 |
| PAHO | Dominica | 30% | 21,595 | 6,091 | 84 | 13 | 643 | 265,359 | 40 | 1,410 |
| PAHO | Dominica | 40% | 28,794 | 4,907 | 60 | 10 | 692 | 347,861 | 70 | 1,680 |
| PAHO | Dominica | 50% | 35,992 | 4,293 | 54 | 8 | 711 | 430,362 | 130 | 4,530 |
| PAHO | Dominica | 60% | 43,190 | 3,918 | 52 | 9 | 711 | 512,864 | 220 | dominated |
| PAHO | Dominica | 70% | 50,389 | 3,828 | 53 | 9 | 702 | 595,366 | 920 | dominated |
| PAHO | El Salvador | 0% | - | 1,413,125 | 23,474 | 10,141 | - | - | - | - |
| PAHO | El Salvador | 20% | 1,297,470 | 635,759 | 7,636 | 1,849 | 108,950 | 16,479,520 | 20 | 150 |
| PAHO | El Salvador | 30% | 1,946,205 | 474,013 | 5,314 | 1,032 | 121,697 | 23,914,768 | 50 | 580 |
| PAHO | El Salvador | 40% | 2,594,940 | 394,282 | 4,308 | 921 | 124,736 | 31,350,016 | 90 | 2,450 |
| PAHO | El Salvador | 50% | 3,243,676 | 340,854 | 3,808 | 694 | 127,291 | 38,785,264 | 140 | 2,910 |
| PAHO | El Salvador | 60% | 3,892,411 | 310,347 | 3,685 | 629 | 128,151 | 46,220,512 | 240 | 8,650 |
| PAHO | El Salvador | 70% | 4,541,146 | 304,060 | 3,672 | 688 | 127,432 | 53,655,760 | 1,180 | dominated |
| PAHO | Grenada | 0% | - | 24,339 | 453 | 169 | - | - | - | - |

| | | | | | | | | | | |
|---|---|---|---|---|---|---|---|---|---|---|
| PAHO | Grenada | 20% | 22,798 | 11,283 | 148 | 25 | 1,649 | 289,560 | 20 | 180 |
| PAHO | Grenada | 30% | 34,197 | 7,394 | 97 | 15 | 1,779 | 420,204 | 30 | 1,000 |
| PAHO | Grenada | 40% | 45,595 | 6,299 | 80 | 14 | 1,809 | 550,848 | 120 | dominated |
| PAHO | Grenada | 50% | 56,614 | 5,309 | 69 | 11 | 1,843 | 681,492 | 130 | 4,110 |
| PAHO | Grenada | 60% | 68,013 | 4,897 | 66 | 11 | 1,843 | 812,136 | 320 | dominated |
| PAHO | Grenada | 70% | 79,412 | 4,768 | 64 | 10 | 1,832 | 942,780 | 1,010 | dominated |
| PAHO | Guyana | 0% | - | 166,527 | 2,591 | 949 | - | - | - | - |
| PAHO | Guyana | 20% | 157,184 | 71,855 | 850 | 117 | 11,631 | 1,996,440 | 20 | 170 |
| PAHO | Guyana | 30% | 235,776 | 50,236 | 556 | 83 | 12,167 | 2,897,196 | 40 | 1,680 |
| PAHO | Guyana | 40% | 314,369 | 39,418 | 447 | 70 | 12,333 | 3,797,952 | 80 | 5,430 |
| PAHO | Guyana | 50% | 392,961 | 35,373 | 396 | 61 | 12,450 | 4,698,708 | 220 | 7,690 |
| PAHO | Guyana | 60% | 471,553 | 31,677 | 369 | 61 | 12,479 | 5,599,464 | 240 | 31,400 |
| PAHO | Guyana | 70% | 550,145 | 31,661 | 373 | 60 | 12,489 | 6,500,220 | 56,300 | 88,650 |
| PAHO | Haiti | 0% | - | 2,480,482 | 34,217 | 12,497 | - | - | - | - |
| PAHO | Haiti | 20% | 2,280,172 | 1,246,911 | 12,417 | 2,337 | 141,726 | 28,961,080 | 20 | 200 |
| PAHO | Haiti | 30% | 3,420,258 | 890,850 | 8,198 | 1,174 | 160,222 | 42,027,772 | 40 | 710 |
| PAHO | Haiti | 40% | 4,560,344 | 759,818 | 6,773 | 844 | 164,103 | 55,094,464 | 100 | 3,370 |
| PAHO | Haiti | 50% | 5,662,427 | 651,202 | 5,975 | 867 | 164,160 | 68,161,156 | 120 | dominated |
| PAHO | Haiti | 60% | 6,802,513 | 605,298 | 5,621 | 730 | 165,799 | 81,227,848 | 280 | 15,400 |
| PAHO | Haiti | 70% | 7,942,599 | 572,848 | 5,405 | 764 | 165,027 | 94,294,540 | 400 | dominated |
| PAHO | Honduras | 0% | - | 2,212,416 | 29,906 | 9,639 | - | - | - | - |
| PAHO | Honduras | 20% | 1,981,002 | 1,074,444 | 10,401 | 1,407 | 125,266 | 25,161,240 | 20 | 200 |
| PAHO | Honduras | 30% | 2,971,503 | 842,416 | 8,202 | 971 | 134,320 | 36,513,516 | 50 | 1,250 |
| PAHO | Honduras | 40% | 3,962,004 | 623,295 | 6,092 | 812 | 136,284 | 47,865,792 | 50 | 5,780 |
| PAHO | Honduras | 50% | 4,919,488 | 553,419 | 5,528 | 684 | 138,132 | 59,218,068 | 160 | 6,140 |
| PAHO | Honduras | 60% | 5,909,989 | 504,503 | 4,943 | 575 | 139,508 | 70,570,344 | 230 | 8,250 |
| PAHO | Honduras | 70% | 6,900,490 | 479,341 | 4,923 | 545 | 140,103 | 81,922,620 | 450 | 19,080 |
| PAHO | Nicaragua | 0% | - | 1,438,696 | 20,959 | 3,483 | - | - | - | - |
| PAHO | Nicaragua | 20% | 1,324,268 | 638,891 | 6,648 | 993 | 28,006 | 16,819,880 | 20 | 600 |
| PAHO | Nicaragua | 30% | 1,986,401 | 441,303 | 4,463 | 523 | 34,943 | 24,408,692 | 40 | 1,090 |
| PAHO | Nicaragua | 40% | 2,648,535 | 360,806 | 3,768 | 490 | 35,795 | 31,997,504 | 90 | 8,910 |
| PAHO | Nicaragua | 50% | 3,310,669 | 311,863 | 2,987 | 470 | 36,428 | 39,586,316 | 160 | dominated |
| PAHO | Nicaragua | 60% | 3,972,803 | 291,567 | 2,894 | 404 | 37,256 | 47,175,128 | 370 | 10,390 |
| PAHO | Nicaragua | 70% | 4,634,936 | 283,382 | 2,914 | 391 | 37,374 | 54,763,940 | 930 | 64,110 |
| PAHO | Saint Lucia | 0% | - | 42,065 | 791 | 190 | - | - | - | - |
| PAHO | Saint Lucia | 20% | 36,396 | 20,645 | 276 | 40 | 1,883 | 462,280 | 20 | 250 |
| PAHO | Saint Lucia | 30% | 54,595 | 14,039 | 189 | 27 | 2,058 | 670,852 | 30 | 1,190 |
| PAHO | Saint Lucia | 40% | 72,793 | 12,260 | 172 | 28 | 2,071 | 879,424 | 120 | dominated |
| PAHO | Saint Lucia | 50% | 90,991 | 10,693 | 149 | 22 | 2,158 | 1,087,996 | 130 | 4,180 |
| PAHO | Saint Lucia | 60% | 109,189 | 9,546 | 135 | 20 | 2,167 | 1,296,568 | 180 | dominated |
| PAHO | Saint Lucia | 70% | 127,387 | 9,177 | 130 | 18 | 2,185 | 1,505,140 | 570 | 15,110 |
| PAHO | Saint Vincent and the Grenad | 0% | - | 23,716 | 437 | 167 | - | - | - | - |
| PAHO | Saint Vincent and the Grenad | 20% | 22,198 | 11,402 | 154 | 28 | 1,632 | 281,940 | 20 | 170 |
| PAHO | Saint Vincent and the Grenad | 30% | 33,297 | 8,726 | 108 | 15 | 1,792 | 409,146 | 50 | 800 |
| PAHO | Saint Vincent and the Grenad | 40% | 44,396 | 6,753 | 88 | 14 | 1,829 | 536,352 | 60 | 3,380 |
| PAHO | Saint Vincent and the Grenad | 50% | 55,124 | 6,129 | 74 | 11 | 1,856 | 663,558 | dominated | 4,780 |
| PAHO | Saint Vincent and the Grenad | 60% | 66,223 | 5,464 | 71 | 11 | 1,864 | 790,764 | 200 | 16,250 |
| PAHO | Saint Vincent and the Grenad | 70% | 77,322 | 5,271 | 68 | 9 | 1,864 | 917,970 | 660 | dominated |
| SEARO | All countries | 0% | - | 125,648,706 | 1,924,766 | 651,884 | - | - | - | - |
| SEARO | All countries | 20% | 114,304,732 | 56,725,991 | 645,652 | 118,389 | 6,944,554 | 1,451,815,270 | 20 | 210 |
| SEARO | All countries | 30% | 171,457,097 | 39,302,339 | 453,488 | 79,003 | 7,585,453 | 2,106,846,884 | 40 | 1,020 |
| SEARO | All countries | 40% | 228,609,462 | 32,143,940 | 363,069 | 57,777 | 7,879,863 | 2,761,878,498 | 90 | 2,220 |
| SEARO | All countries | 50% | 284,753,064 | 27,828,013 | 311,764 | 49,056 | 7,982,214 | 3,416,910,112 | 150 | 6,400 |

| | | | | | | | | | | |
|---|---|---|---|---|---|---|---|---|---|---|
| SEARO | All countries | 60% | 341,905,428 | 25,509,387 | 296,198 | 49,981 | 7,984,484 | 4,071,941,726 | 280 | 288,610 |
| SEARO | All countries | 70% | 399,057,794 | 24,434,597 | 289,888 | 49,156 | 7,983,492 | 4,726,973,340 | 610 | dominated |
| SEARO | Bangladesh | 0% | - | 37,951,856 | 569,491 | 225,128 | - | - | - | - |
| SEARO | Bangladesh | 20% | 32,934,306 | 18,566,266 | 206,025 | 47,430 | 2,496,838 | 418,307,520 | 20 | 170 |
| SEARO | Bangladesh | 30% | 49,401,459 | 13,091,543 | 144,431 | 29,809 | 2,813,536 | 607,039,968 | 30 | 600 |
| SEARO | Bangladesh | 40% | 65,868,612 | 10,585,321 | 114,293 | 18,116 | 2,984,818 | 795,772,416 | 80 | 1,100 |
| SEARO | Bangladesh | 50% | 82,335,766 | 9,622,720 | 101,777 | 17,622 | 2,994,178 | 984,504,864 | 200 | 20,160 |
| SEARO | Bangladesh | 60% | 98,802,919 | 8,788,904 | 97,331 | 18,280 | 2,985,273 | 1,173,237,312 | 230 | dominated |
| SEARO | Bangladesh | 70% | 115,270,072 | 8,409,957 | 94,531 | 19,598 | 2,970,218 | 1,361,969,760 | 500 | dominated |
| SEARO | Bhutan | 0% | - | 160,054 | 2,414 | 685 | - | - | - | - |
| SEARO | Bhutan | 20% | 154,185 | 71,250 | 823 | 120 | 7,927 | 1,958,340 | 20 | 250 |
| SEARO | Bhutan | 30% | 231,277 | 53,042 | 611 | 96 | 8,270 | 2,841,906 | 50 | dominated |
| SEARO | Bhutan | 40% | 308,369 | 40,909 | 460 | 65 | 8,805 | 3,725,472 | 70 | 2,010 |
| SEARO | Bhutan | 50% | 385,461 | 33,191 | 378 | 56 | 8,888 | 4,609,038 | 110 | 10,710 |
| SEARO | Bhutan | 60% | 462,554 | 30,913 | 374 | 66 | 8,741 | 5,492,604 | 390 | dominated |
| SEARO | Bhutan | 70% | 539,646 | 30,359 | 372 | 59 | 8,858 | 6,376,170 | 1,590 | dominated |
| SEARO | Indonesia | 0% | - | 58,959,797 | 882,112 | 247,265 | - | - | - | - |
| SEARO | Indonesia | 20% | 54,699,130 | 25,606,676 | 289,387 | 39,114 | 2,661,952 | 694,748,420 | 20 | 260 |
| SEARO | Indonesia | 30% | 82,048,694 | 17,011,489 | 199,125 | 26,258 | 2,845,198 | 1,008,205,778 | 40 | 1,710 |
| SEARO | Indonesia | 40% | 109,398,259 | 14,339,170 | 163,840 | 23,249 | 2,887,391 | 1,321,663,136 | 120 | dominated |
| SEARO | Indonesia | 50% | 135,836,172 | 12,037,747 | 135,941 | 17,232 | 2,948,094 | 1,635,120,494 | 140 | 6,090 |
| SEARO | Indonesia | 60% | 163,185,736 | 10,919,038 | 125,821 | 16,958 | 2,961,913 | 1,948,577,852 | 280 | dominated |
| SEARO | Indonesia | 70% | 190,535,301 | 10,484,410 | 125,274 | 14,770 | 2,976,953 | 2,262,035,210 | 720 | 21,720 |
| SEARO | Maldives | 0% | - | 116,745 | 1,728 | 376 | - | - | - | - |
| SEARO | Maldives | 20% | 108,189 | 51,658 | 627 | 69 | 5,006 | 1,374,140 | 20 | 270 |
| SEARO | Maldives | 30% | 162,284 | 41,306 | 469 | 52 | 5,191 | 1,994,126 | 60 | dominated |
| SEARO | Maldives | 40% | 216,378 | 31,933 | 371 | 40 | 5,465 | 2,614,112 | 70 | 2,700 |
| SEARO | Maldives | 50% | 270,473 | 27,322 | 312 | 37 | 5,509 | 3,234,098 | 130 | dominated |
| SEARO | Maldives | 60% | 324,568 | 25,383 | 294 | 32 | 5,587 | 3,854,084 | 320 | 10,210 |
| SEARO | Maldives | 70% | 378,662 | 24,534 | 285 | 33 | 5,591 | 4,474,070 | 730 | 138,630 |
| SEARO | Myanmar | 0% | - | 11,829,985 | 182,491 | 69,427 | - | - | - | - |
| SEARO | Myanmar | 20% | 10,880,912 | 4,734,160 | 53,757 | 8,161 | 773,867 | 138,201,400 | 20 | 180 |
| SEARO | Myanmar | 30% | 16,321,368 | 3,509,880 | 40,155 | 5,985 | 804,659 | 200,555,260 | 50 | 2,030 |
| SEARO | Myanmar | 40% | 21,761,824 | 2,652,868 | 28,892 | 3,700 | 832,380 | 262,909,120 | 70 | 2,250 |
| SEARO | Myanmar | 50% | 27,202,280 | 2,265,034 | 26,171 | 3,537 | 836,411 | 325,262,980 | 160 | 15,470 |
| SEARO | Myanmar | 60% | 32,642,735 | 2,109,421 | 26,443 | 3,754 | 835,287 | 387,616,840 | 400 | dominated |
| SEARO | Myanmar | 70% | 38,083,191 | 2,023,943 | 25,246 | 3,809 | 835,437 | 449,970,700 | 730 | dominated |
| SEARO | Nepal | 0% | - | 6,060,613 | 86,071 | 21,882 | - | - | - | - |
| SEARO | Nepal | 20% | 5,826,817 | 2,859,039 | 29,778 | 4,021 | 223,495 | 74,007,980 | 20 | 330 |
| SEARO | Nepal | 30% | 8,740,226 | 1,935,309 | 19,493 | 2,710 | 242,048 | 107,398,982 | 40 | 1,800 |
| SEARO | Nepal | 40% | 11,653,635 | 1,592,221 | 16,929 | 2,214 | 248,725 | 140,789,984 | 100 | 5,000 |
| SEARO | Nepal | 50% | 14,469,930 | 1,344,265 | 14,306 | 1,894 | 253,120 | 174,180,986 | 130 | 7,600 |
| SEARO | Nepal | 60% | 17,383,338 | 1,251,114 | 12,704 | 1,661 | 255,575 | 207,571,988 | 360 | 13,600 |
| SEARO | Nepal | 70% | 20,296,747 | 1,191,645 | 12,500 | 1,777 | 252,728 | 240,962,990 | 560 | dominated |
| SEARO | Democratic People's Republic | 0% | - | 5,711,786 | 109,844 | 54,368 | - | - | - | - |
| SEARO | Democratic People's Republic | 20% | 5,155,247 | 2,654,058 | 37,379 | 15,570 | 425,441 | 65,478,190 | 20 | 150 |
| SEARO | Democratic People's Republic | 30% | 7,732,871 | 2,094,941 | 28,614 | 11,033 | 502,303 | 95,020,712 | 50 | 380 |
| SEARO | Democratic People's Republic | 40% | 10,310,495 | 1,632,315 | 22,402 | 8,120 | 540,129 | 124,563,234 | 60 | 780 |
| SEARO | Democratic People's Republic | 50% | 12,888,119 | 1,423,016 | 19,257 | 6,728 | 559,109 | 154,105,756 | 140 | 1,560 |
| SEARO | Democratic People's Republic | 60% | 15,465,742 | 1,345,963 | 19,747 | 7,167 | 555,768 | 183,648,278 | 380 | dominated |
| SEARO | Democratic People's Republic | 70% | 18,043,366 | 1,276,825 | 18,458 | 6,857 | 559,752 | 213,190,800 | 430 | 91,980 |
| SEARO | Sri Lanka | 0% | - | 4,573,645 | 87,155 | 31,457 | - | - | - | - |
| SEARO | Sri Lanka | 20% | 4,282,372 | 2,060,948 | 26,789 | 3,726 | 334,926 | 54,391,560 | 20 | 160 |

| | | | | | | | | | | |
|---|---|---|---|---|---|---|---|---|---|---|
| SEARO | Sri Lanka | 30% | 6,423,558 | 1,478,251 | 19,851 | 2,977 | 347,590 | 78,932,004 | 40 | 1,940 |
| SEARO | Sri Lanka | 40% | 8,564,743 | 1,197,878 | 15,290 | 2,206 | 355,187 | 103,472,448 | 90 | 3,230 |
| SEARO | Sri Lanka | 50% | 10,705,929 | 1,014,467 | 13,105 | 1,884 | 359,922 | 128,012,892 | 130 | 5,180 |
| SEARO | Sri Lanka | 60% | 12,847,115 | 983,866 | 12,977 | 1,992 | 359,407 | 152,553,336 | dominated | dominated |
| SEARO | Sri Lanka | 70% | 14,988,301 | 937,869 | 12,720 | 2,184 | 356,998 | 177,093,780 | 640 | dominated |
| SEARO | Timor-Leste | 0% | - | 284,225 | 3,460 | 1,296 | - | - | - | - |
| SEARO | Timor-Leste | 20% | 263,574 | 121,936 | 1,087 | 178 | 15,103 | 3,347,720 | 20 | 220 |
| SEARO | Timor-Leste | 30% | 395,360 | 86,578 | 739 | 83 | 16,658 | 4,858,148 | 40 | 970 |
| SEARO | Timor-Leste | 40% | 527,147 | 71,325 | 592 | 67 | 16,964 | 6,368,576 | 100 | 4,930 |
| SEARO | Timor-Leste | 50% | 658,934 | 60,251 | 517 | 66 | 16,983 | 7,879,004 | 140 | 80,770 |
| SEARO | Timor-Leste | 60% | 790,721 | 54,785 | 507 | 71 | 16,934 | 9,389,432 | 280 | dominated |
| SEARO | Timor-Leste | 70% | 922,508 | 55,055 | 502 | 69 | 16,957 | 10,899,860 | dominated | dominated |
| WPRO | All countries | 0% | - | 52,678,660 | 822,791 | 307,679 | - | - | - | - |
| WPRO | All countries | 20% | 49,123,084 | 25,317,288 | 281,860 | 72,604 | 3,104,645 | 623,925,564 | 20 | 200 |
| WPRO | All countries | 30% | 73,684,627 | 17,326,496 | 189,433 | 43,948 | 3,569,580 | 905,428,988 | 40 | 610 |
| WPRO | All countries | 40% | 98,247,073 | 14,514,880 | 158,901 | 38,038 | 3,676,346 | 1,186,932,412 | 100 | 2,640 |
| WPRO | All countries | 50% | 122,797,686 | 12,531,101 | 136,423 | 31,309 | 3,772,646 | 1,468,435,836 | 140 | 2,920 |
| WPRO | All countries | 60% | 147,359,228 | 11,598,079 | 129,090 | 30,596 | 3,777,246 | 1,749,939,260 | 300 | dominated |
| WPRO | All countries | 70% | 171,920,771 | 11,130,296 | 124,198 | 28,245 | 3,813,745 | 2,031,442,684 | 600 | 13,700 |
| WPRO | Cambodia | 0% | - | 3,800,563 | 51,411 | 17,287 | - | - | - | - |
| WPRO | Cambodia | 20% | 3,343,466 | 1,757,719 | 17,722 | 2,842 | 196,690 | 42,466,260 | 20 | 220 |
| WPRO | Cambodia | 30% | 5,015,198 | 1,344,074 | 13,509 | 1,622 | 218,371 | 61,626,234 | 50 | 880 |
| WPRO | Cambodia | 40% | 6,686,931 | 969,752 | 8,978 | 1,154 | 224,658 | 80,786,208 | 50 | 3,050 |
| WPRO | Cambodia | 50% | 8,358,664 | 885,355 | 8,159 | 1,087 | 225,362 | 99,946,182 | 230 | dominated |
| WPRO | Cambodia | 60% | 10,030,397 | 805,137 | 7,758 | 1,070 | 226,938 | 119,106,156 | 240 | 16,810 |
| WPRO | Cambodia | 70% | 11,702,130 | 766,834 | 7,457 | 1,020 | 227,159 | 138,266,130 | 500 | 86,780 |
| WPRO | Federated State of Micronesi | 0% | - | 26,133 | 365 | 127 | - | - | - | - |
| WPRO | Federated State of Micronesi | 20% | 22,998 | 12,976 | 135 | 22 | 1,282 | 292,100 | 20 | 230 |
| WPRO | Federated State of Micronesi | 30% | 34,497 | 9,319 | 90 | 13 | 1,427 | 423,890 | 40 | 910 |
| WPRO | Federated State of Micronesi | 40% | 45,995 | 7,825 | 73 | 10 | 1,459 | 555,680 | 90 | 4,090 |
| WPRO | Federated State of Micronesi | 50% | 57,494 | 6,519 | 64 | 8 | 1,484 | 687,470 | 100 | 5,310 |
| WPRO | Federated State of Micronesi | 60% | 68,993 | 6,111 | 62 | 7 | 1,501 | 819,260 | 320 | 7,570 |
| WPRO | Federated State of Micronesi | 70% | 80,492 | 6,040 | 62 | 7 | 1,484 | 951,050 | 1,860 | dominated |
| WPRO | Fiji | 0% | - | 194,324 | 2,845 | 493 | - | - | - | - |
| WPRO | Fiji | 20% | 178,782 | 91,338 | 1,005 | 116 | 3,859 | 2,270,760 | 20 | 590 |
| WPRO | Fiji | 30% | 268,173 | 62,315 | 667 | 98 | 4,145 | 3,295,284 | 40 | dominated |
| WPRO | Fiji | 40% | 357,564 | 48,297 | 505 | 59 | 4,612 | 4,319,808 | 70 | 2,720 |
| WPRO | Fiji | 50% | 446,955 | 42,598 | 460 | 59 | 4,661 | 5,344,332 | 180 | 20,760 |
| WPRO | Fiji | 60% | 536,346 | 37,655 | 434 | 63 | 4,601 | 6,368,856 | 210 | dominated |
| WPRO | Fiji | 70% | 625,737 | 37,054 | 417 | 57 | 4,647 | 7,393,380 | 1,700 | dominated |
| WPRO | Kiribati | 0% | - | 26,023 | 319 | 101 | - | - | - | - |
| WPRO | Kiribati | 20% | 23,598 | 13,147 | 122 | 20 | 1,158 | 299,720 | 20 | 260 |
| WPRO | Kiribati | 30% | 35,396 | 8,985 | 76 | 11 | 1,315 | 434,948 | 30 | 860 |
| WPRO | Kiribati | 40% | 47,195 | 7,305 | 56 | 7 | 1,370 | 570,176 | 80 | 2,480 |
| WPRO | Kiribati | 50% | 58,994 | 6,151 | 49 | 7 | 1,370 | 705,404 | 120 | dominated |
| WPRO | Kiribati | 60% | 70,793 | 5,576 | 47 | 6 | 1,370 | 840,632 | 240 | dominated |
| WPRO | Kiribati | 70% | 82,592 | 5,401 | 45 | 6 | 1,370 | 975,860 | 770 | dominated |
| WPRO | Lao People's Democratic Rep | 0% | - | 1,719,428 | 23,421 | 6,441 | - | - | - | - |
| WPRO | Lao People's Democratic Rep | 20% | 1,455,454 | 826,919 | 8,057 | 779 | 76,124 | 18,486,120 | 20 | 240 |
| WPRO | Lao People's Democratic Rep | 30% | 2,183,182 | 640,100 | 6,157 | 604 | 78,891 | 26,826,708 | 40 | 3,010 |
| WPRO | Lao People's Democratic Rep | 40% | 2,910,909 | 502,087 | 4,425 | 546 | 79,276 | 35,167,296 | 60 | dominated |
| WPRO | Lao People's Democratic Rep | 50% | 3,638,636 | 417,845 | 3,712 | 422 | 81,402 | 43,507,884 | 100 | 6,640 |
| WPRO | Lao People's Democratic Rep | 60% | 4,366,363 | 392,226 | 3,493 | 378 | 81,912 | 51,848,472 | 330 | 16,350 |

| | | | | | | | | | | |
|---|---|---|---|---|---|---|---|---|---|---|
| WPRO | Lao People's Democratic Rep | 70% | 5,094,091 | 386,011 | 3,559 | 408 | 81,757 | 60,189,060 | 1,340 | dominated |
| WPRO | Marshall Islands | 0% | - | 13,147 | 169 | 28 | - | - | - | - |
| WPRO | Marshall Islands | 20% | 11,838 | 6,120 | 58 | 7 | 241 | 150,353 | 20 | 620 |
| WPRO | Marshall Islands | 30% | 17,756 | 4,297 | 35 | 4 | 284 | 218,189 | 40 | 1,550 |
| WPRO | Marshall Islands | 40% | 23,873 | 3,605 | 32 | 4 | 295 | 286,025 | 100 | 6,520 |
| WPRO | Marshall Islands | 50% | 29,791 | 3,104 | 26 | 3 | 302 | 353,862 | 140 | 8,900 |
| WPRO | Marshall Islands | 60% | 35,710 | 2,940 | 27 | 3 | 310 | 421,698 | dominated | 8,900 |
| WPRO | Marshall Islands | 70% | 41,629 | 2,767 | 26 | 3 | 310 | 489,534 | 400 | dominated |
| WPRO | Mongolia | 0% | - | 662,984 | 9,070 | 1,367 | - | - | - | - |
| WPRO | Mongolia | 20% | 655,734 | 284,935 | 3,072 | 364 | 11,034 | 8,328,660 | 20 | 750 |
| WPRO | Mongolia | 30% | 983,602 | 230,891 | 2,263 | 233 | 12,961 | 12,086,394 | 70 | 1,950 |
| WPRO | Mongolia | 40% | 1,311,469 | 177,259 | 1,833 | 226 | 13,228 | 15,844,128 | 70 | dominated |
| WPRO | Mongolia | 50% | 1,628,407 | 152,192 | 1,469 | 157 | 13,950 | 19,601,862 | 150 | 7,600 |
| WPRO | Mongolia | 60% | 1,956,274 | 140,331 | 1,505 | 180 | 13,784 | 23,359,596 | 320 | dominated |
| WPRO | Mongolia | 70% | 2,284,142 | 132,514 | 1,384 | 131 | 14,318 | 27,117,330 | 480 | 20,450 |
| WPRO | Papua New Guinea | 0% | - | 1,941,025 | 23,513 | 8,097 | - | - | - | - |
| WPRO | Papua New Guinea | 20% | 1,789,221 | 715,178 | 6,379 | 680 | 95,939 | 22,725,380 | 20 | 240 |
| WPRO | Papua New Guinea | 30% | 2,683,832 | 531,819 | 4,348 | 492 | 99,713 | 32,978,642 | 60 | 2,720 |
| WPRO | Papua New Guinea | 40% | 3,578,442 | 431,120 | 3,964 | 456 | 99,317 | 43,231,904 | 100 | dominated |
| WPRO | Papua New Guinea | 50% | 4,473,053 | 393,775 | 3,838 | 358 | 101,153 | 53,485,166 | dominated | 14,240 |
| WPRO | Papua New Guinea | 60% | 5,367,663 | 343,153 | 3,382 | 322 | 101,136 | 63,738,428 | 230 | dominated |
| WPRO | Papua New Guinea | 70% | 6,262,274 | 330,296 | 3,310 | 349 | 101,033 | 73,991,690 | 800 | dominated |
| WPRO | Philippines | 0% | - | 22,438,025 | 328,086 | 97,746 | - | - | - | - |
| WPRO | Philippines | 20% | 21,914,008 | 11,047,518 | 110,677 | 15,122 | 1,106,879 | 278,335,740 | 20 | 250 |
| WPRO | Philippines | 30% | 32,871,013 | 7,253,386 | 73,091 | 11,177 | 1,167,035 | 403,915,566 | 30 | 2,090 |
| WPRO | Philippines | 40% | 43,828,017 | 5,768,892 | 55,338 | 8,657 | 1,197,964 | 529,495,392 | 80 | 4,060 |
| WPRO | Philippines | 50% | 54,785,021 | 5,099,790 | 50,407 | 7,561 | 1,208,745 | 655,075,218 | 190 | 11,650 |
| WPRO | Philippines | 60% | 65,742,025 | 4,721,626 | 47,668 | 7,123 | 1,213,720 | 780,655,044 | 330 | dominated |
| WPRO | Philippines | 70% | 76,699,029 | 4,580,267 | 47,120 | 6,575 | 1,222,983 | 906,234,870 | 890 | 17,640 |
| WPRO | Samoa | 0% | - | 42,735 | 560 | 162 | - | - | - | - |
| WPRO | Samoa | 20% | 39,996 | 18,207 | 171 | 26 | 1,693 | 508,000 | 20 | 300 |
| WPRO | Samoa | 30% | 59,994 | 13,315 | 114 | 16 | 1,829 | 737,200 | 50 | 1,690 |
| WPRO | Samoa | 40% | 80,659 | 10,763 | 92 | 14 | 1,874 | 966,400 | 90 | 5,070 |
| WPRO | Samoa | 50% | 100,657 | 9,007 | 83 | 11 | 1,890 | 1,195,600 | 130 | 14,930 |
| WPRO | Samoa | 60% | 120,655 | 8,177 | 76 | 11 | 1,879 | 1,424,800 | 280 | dominated |
| WPRO | Samoa | 70% | 140,653 | 7,885 | 74 | 10 | 1,897 | 1,654,000 | 780 | 59,710 |
| WPRO | Solomon Islands | 0% | - | 151,062 | 1,759 | 820 | - | - | - | - |
| WPRO | Solomon Islands | 20% | 137,386 | 69,853 | 563 | 214 | 8,764 | 1,744,980 | 20 | 200 |
| WPRO | Solomon Islands | 30% | 206,079 | 46,300 | 350 | 150 | 9,938 | 2,532,282 | 30 | 670 |
| WPRO | Solomon Islands | 40% | 274,773 | 35,676 | 284 | 113 | 10,592 | 3,319,584 | 70 | 1,200 |
| WPRO | Solomon Islands | 50% | 343,466 | 30,007 | 231 | 100 | 10,719 | 4,106,886 | 140 | dominated |
| WPRO | Solomon Islands | 60% | 412,159 | 27,554 | 233 | 95 | 10,854 | 4,894,188 | 320 | 6,020 |
| WPRO | Solomon Islands | 70% | 480,852 | 26,264 | 227 | 93 | 10,929 | 5,681,490 | 610 | 10,450 |
| WPRO | Tonga | 0% | - | 22,205 | 296 | 55 | - | - | - | - |
| WPRO | Tonga | 20% | 20,798 | 10,133 | 96 | 12 | 435 | 264,160 | 20 | 610 |
| WPRO | Tonga | 30% | 31,197 | 7,255 | 69 | 10 | 461 | 383,344 | 40 | dominated |
| WPRO | Tonga | 40% | 41,596 | 5,676 | 53 | 7 | 505 | 502,528 | 80 | 3,440 |
| WPRO | Tonga | 50% | 51,995 | 4,839 | 46 | 6 | 512 | 621,712 | 140 | dominated |
| WPRO | Tonga | 60% | 62,394 | 4,600 | 44 | 5 | 537 | 740,896 | 500 | 7,290 |
| WPRO | Tonga | 70% | 72,793 | 4,488 | 45 | 5 | 519 | 860,080 | 1,060 | dominated |
| WPRO | Tuvalu | 0% | - | 2,628 | 33 | 5 | - | - | - | - |
| WPRO | Tuvalu | 20% | 2,358 | 1,293 | 13 | 2 | 33 | 29,952 | 20 | dominated |
| WPRO | Tuvalu | 30% | 3,537 | 944 | 9 | 1 | 51 | 43,465 | 40 | 850 |

| | | | | | | | | | | |
|---|---|---|---|---|---|---|---|---|---|---|
| WPRO | Tuvalu | 40% | 4,756 | 725 | 7 | 1 | 51 | 56,979 | 60 | dominated |
| WPRO | Tuvalu | 50% | 5,935 | 641 | 6 | 1 | 51 | 70,493 | 160 | dominated |
| WPRO | Tuvalu | 60% | 7,114 | 600 | 6 | 1 | 51 | 84,006 | 330 | dominated |
| WPRO | Tuvalu | 70% | 8,293 | 572 | 6 | 1 | 51 | 97,520 | 480 | dominated |
| WPRO | Vanuatu | 0% | - | 67,569 | 835 | 226 | - | - | - | - |
| WPRO | Vanuatu | 20% | 61,594 | 28,866 | 241 | 30 | 2,576 | 782,320 | 20 | 300 |
| WPRO | Vanuatu | 30% | 92,391 | 22,584 | 174 | 23 | 2,642 | 1,135,288 | 60 | dominated |
| WPRO | Vanuatu | 40% | 123,188 | 17,371 | 133 | 16 | 2,743 | 1,488,256 | 70 | 4,210 |
| WPRO | Vanuatu | 50% | 153,985 | 15,056 | 125 | 17 | 2,765 | 1,841,224 | 150 | 16,290 |
| WPRO | Vanuatu | 60% | 184,782 | 13,899 | 111 | 15 | 2,760 | 2,194,192 | 310 | dominated |
| WPRO | Vanuatu | 70% | 215,578 | 13,358 | 116 | 15 | 2,772 | 2,547,160 | 650 | 102,080 |
| WPRO | Vietnam | 0% | - | 21,570,809 | 380,109 | 174,724 | - | - | - | - |
| WPRO | Vietnam | 20% | 19,465,853 | 10,433,086 | 133,549 | 52,368 | 1,597,937 | 247,241,060 | 20 | 150 |
| WPRO | Vietnam | 30% | 29,198,780 | 7,150,912 | 88,481 | 29,494 | 1,970,515 | 358,791,554 | 30 | 300 |
| WPRO | Vietnam | 40% | 38,931,706 | 6,528,527 | 83,128 | 26,768 | 2,038,400 | 470,342,048 | dominated | dominated |
| WPRO | Vietnam | 50% | 48,664,633 | 5,464,222 | 67,748 | 21,512 | 2,118,279 | 581,892,542 | 130 | 1,510 |
| WPRO | Vietnam | 60% | 58,397,560 | 5,088,494 | 64,244 | 21,317 | 2,115,893 | 693,443,036 | 300 | dominated |
| WPRO | Vietnam | 70% | 68,130,486 | 4,830,545 | 60,350 | 19,565 | 2,142,516 | 804,993,530 | 430 | 9,200 |
